# Positions of the International Society for Clinical Densitometry and their Etiology: A Scoping Review

**DOI:** 10.1101/2023.01.29.23285144

**Authors:** John Shepherd

## Abstract

The International Society for Clinical Densitometry convenes a Position Development Conference (PDC) every 2 to 3 years to make recommendations for guidelines and standards in the field of musculoskeletal measurement and assessment. The recommendations pertain to clinically relevant issues regarding the acquisition, quality control, interpretation, and reporting of measures of various aspects of musculoskeletal health. These PDCs have been meeting since 2002 and have generated 214 Adult, 26 FRAX, 41 pediatric, and 9 general nomenclature consideration positions, for a total of 290 positions. All positions are justified by detailed documents that present the background and rationale for each position. However, the linkage to these publications is not maintained by the ISCD or any other publication such that physicians cannot easily understand the etiology of the positions. Further, the wording of many positions has changed over the years after being reviewed by subsequent PDCs. This Scoping review captures the references, changes, and timeline associated with each position through the 2019 PDC. It is meant to serve as a guide to clinicians and researchers for intelligent use and application of the positions.

## INTRODUCTION

The International Society for Clinical Densitometry (ISCD) is a non-profit professional organization dedicated to the advancement of assessment of musculoskeletal health, particularly (but not limited to) bone densitometry. A major focus of the ISCD is the development of guidelines and establishment of standards for bone densitometry, assessment of fracture risk, and other aspects of musculoskeletal measurement. The Society conducts Position Development Conferences (PCD) approximately every two to three years to develop guidelines and standards (expressed as Position Statements) for new technologies used to assess musculoskeletal health and fracture risk, and to update older guidelines and standards as new data become available. The ISCD Official Positions are widely used by clinicians and densitometry technologists as a reference regarding the indications for, acquisition of, and interpretation and reporting of measures of musculoskeletal health, as well as the incorporation of those measures into fracture risk assessment. The curricula of the densitometry educational courses provided by ISCD are largely based on these Positions.

The ISCD PDC process is designed to summarize and use the best scientific evidence available to develop and update Position Statements regarding musculoskeletal assessment. Since musculoskeletal assessment technologies are evolving, some clinically important issues are addressed in the absence of robust evidence and are largely based on expert opinion. However, the PDC process grades and highlights the limitations of the available evidence pertinent to each statement and indicates where additional research is needed to improve the scientific evidence on which Positions are based and to resolve areas of ambiguity and controversy.

The primary way for clinicians to learn about the positions and refer to them is through the official PDC documents from the ISCD for Adults, FRAX, and Pediatrics. The linkages back to the primary peer-reviewed publications, that serve as the rationale for each position, are not presented in these summary documents. Thus, it is very difficult to find the details on the positions, to know if the position is still valid, or is valid for a particular patient population. In this scoping review, each current position is linked to the ISCD summaries of its associated PDC where it was developed and to the associated publications. We also track the changes in wording, if any, that occurred in subsequent PDC.

### Scoping Question

How and why have the ISCD positions evolved over time regarding the diagnosis of osteoporosis and the use of the densitometry system?

### Rationale

Positions are broadly used in clinical practice and presented without reference to the core publications. Further, there are no means for practicing physicians and researchers to easily understand how the wording of a position has changed over time, the quality of the evidence, the regional applicability, and outstanding controversies associated with the creation of the position.

### Objectives

The objective of the review is to capture each position statement and offer its etiology and pedigree.

## METHODOLOGY

A Scoping review was performed following the methodological guidelines of Peters et al.^2^ A Scoping review was chosen versus a systematic review since all of the literature on the topic was from definitive resources but the clarity of the concepts and definitions of the positions had been lost over time. The protocol for this review is self-contained in this manuscript.

### Eligibility criteria

We considered only the peer-reviewed accepted publications in the English language from the years 2001 through 2022 that were directly attributed to the deliberations of the task forces and executive committees of the PDCs.

### Information sources

It was known *a priori* that all position papers were published in the Journal for Clinical Densitometry. PubMed, Google Scholar Search and Publish or Perish ^3^ were used to find all relevant documents and their associated citations. In addition to peer review publications, we used the most recent form of the Positions distributed by the ISCD. These consist of three documents found on their website ^4^: The 2019 ISCD Official Positions Pediatric, the 2019 ISCD Official Positions Adult, and the 2019 Official Positions in FRAX. The most recent search was performed in January, 2023.

### Search

We used the following search parameters: Journal name: “Journal of Clinical Densitometry”; Keywords: “Position Development Conference”, “Positions”, “Executive Summary”; Years: 2003-2004, 2005-2006, 2007-2008, etc.

### Selection of sources of evidence

Publications were selected that were a direct result of the PDC it was associated with and published by the participants. The relevant publications were generally in the form of an Executive summary that summarized new and previous positions were followed in the same issue by publications authored by each task force providing background and discussion on each question examined.

### Data charting process

We took the approach of starting with the current ISCD position documents and their wording. We then traced back through all the identified Executive Summaries to find when each position was developed. We then noted exact wording changes since the conception of the position and reviewed the publications to determine the rationale for any wording changes. We organized the positions using the same headings and order one finds in the ISCD position documents. We gave each position a number prefixed by a letter signifying which document it is found with A=Adult, F=FRAX, P=Pediactric. We created a table that catalogs each position by the conference it was discussed and developed and the associated publications. Wording changes to each position are noted in the footnotes.

### Synthesis of results

Besides tracking the wording changes for each position, we also note the relative citations related to the positions as a whole by years.

## RESULTS

In total, there have been 10 conferences that resulted in 72 publications generated from 233 authors. See Table 1. From these publications, 5978 citations have been generated. Figure 1 is a summary of the number of citations per year for all PDC publications. In total 219 Adults, 28 FRAX-specific, and 41 Pediatric positions were found. In addition, there were 9 Nomenclature Positions discovered that are not specific to adults or children. To keep track of the positions and which ISCD document they are contained, a prefix was created where A=adult, F=FRAX, P=Pediatric and Nomenclature positions were given the prefix N. Executive summaries of the position statements from prior PDC’s held in 2001, 2003, 2005, 2007, 2010, 2013, 2015, 2019 have been published. ^5 6 7 8 9 10 11 12 13 14 15^

**Table 1.** Summary of Publications.

|  |  |
| --- | --- |
| Total Publications | 72 |
| Publication with the most citations | Engleke et al. <sup>1</sup><br>404 citations |
| Total Times Cited from All Publications | 6775 |
| Publications >= 100 citations | 22 |
| Publications with >= 200 citations | 9 |
| Publications with >= 300 citations | 4 |
| Total authors | 233 |
| Top 5 authors of the most PDC publications | Shepherd JA 17<br>Lewiecki EM 15<br>Schousboe JT 12<br>Binkley NC 12<br>Hans DB 11 |

**Figure 1.**
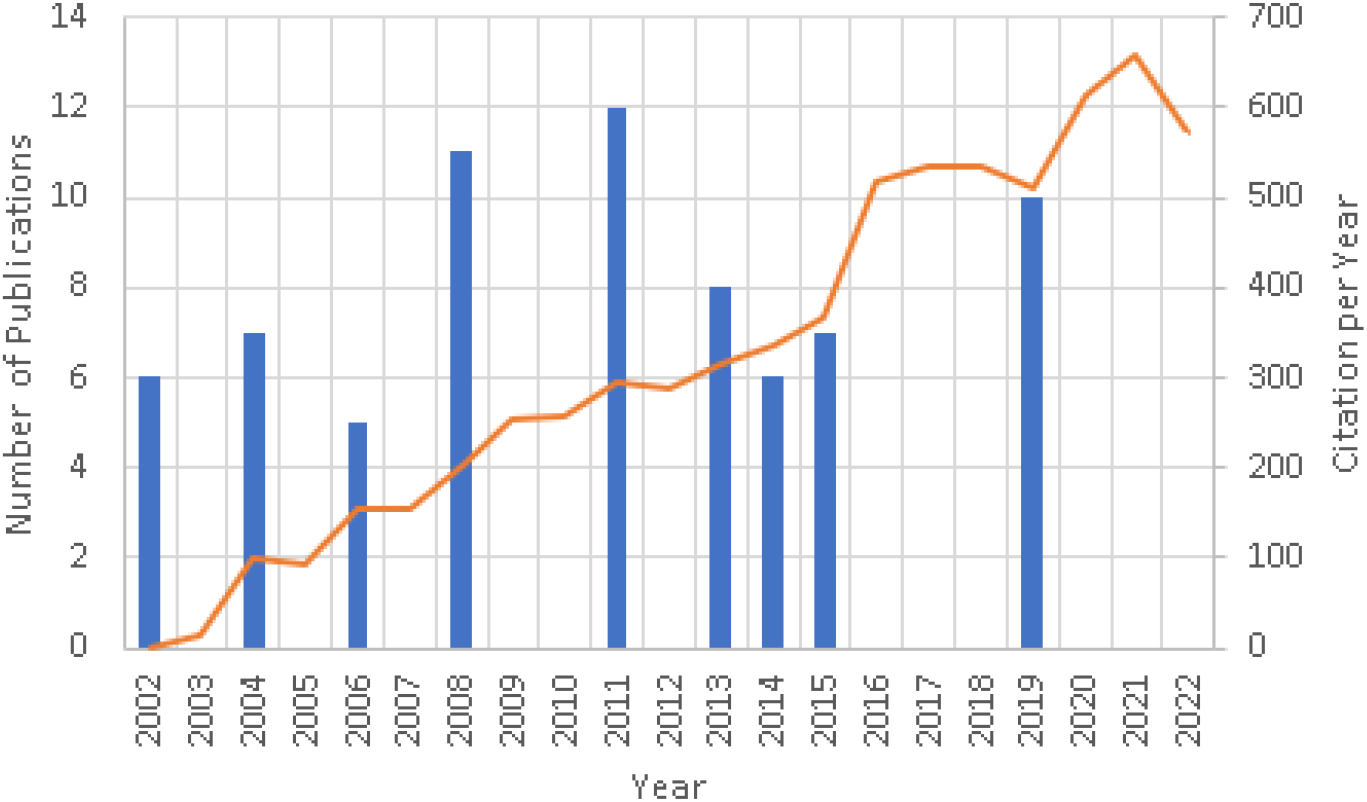
Publication year of all PDC papers with number of citations per year.

### *2002* PDC: Denver Colorado (July 20-22, 2002) ^5 16 17 18 19 20^

This was the first ISCD PDC and generated six papers. The methods for developing positions were not yet standardized. The RAND method of Quality, Strength and Applicability, as was used at future conferences, had yet to be considered. The positions were presented in a narrative form in the executive summary and not summarized as position statements. This PDC addressed the most pressing questions of the time: Diagnosis of Osteoporosis by Central DXA Skeletal Sites ^17^, Osteoporosis Diagnosis in Males and Non-Caucasians ^18^, Serial BMD Measurements in Patient Management ^19^, and Peripheral Skeletal Site Measurements in Osteoporosis Diagnosis ^20^. These topics were considered pressing areas that needed clarification. Topics without controversy in the field were not addressed, but would be in future conferences.

### 2003 PDC: Cincinnati, Ohio (July 2003) ^6 21 22 23 24 25 26^

Like the first PDC, the methods for researching and forming positions had yet to be standardized. However, formal literature searches were performed with task forces. An expert panel reviewed the proposed positions, but no RAND criteria were followed especially for the grading of the position. This was the first PDC that summarized all ISCD positions in a comprehensive Executive Summary ^21^ organized into 15 categories. The 2003 PDC considered five categories for new positions including: the diagnosis of osteoporosis in men, premenopausal women, and children ^23^, technical standardization for dual-energy X-ray absorptiometry (DXA) ^24^, indications for bone densitometry ^25^, Reporting of bone density results, and nomenclature and decimal places for bone densitometry ^26^. There were now 98 bulletized positions from the society. Many of these were consolidated at later PDCs. Also at this PDC, the Canadian Panel of the ISCD issued an update to their standards of guidelines for performing DXA scans ^27^ that had been developed in coordination with the first PDC ^28^. These and subsequent works were regional interpretations of the official positions. Thus, the Canadian-specific and others similar works by regional societies were not included in the scope of the review.

### 2005 PDC: Vancouver, British Columbia, Canada (July 15–17, 2005) ^7 29 30 31 32^

Topics for consideration at 2005 PDC were developed by the ISCD Scientific Advisory Committee and the PDC Steering Committee. Specifically, these topics were the following: technical standardization, vertebral fracture assessment, application of the 1994 World Health Organization (WHO) classification to various skeletal sites, and application of the 1994 WHO classification to populations. It was the first time the positions addressed populations other than Caucasian women. The expert panel included representatives of the American Society for Bone and Mineral Research (ASBMR) and the International Osteoporosis Foundation (IOF), and the positions from this PDC were endorsed by both organizations. Endorsements like these were not always the case for previous and future PDCs. There was no substantive change in the format of the PDC or in its preparation from previous efforts.

### 2007 PDC: Lansdowne, Virginia (July 20–22, 2007) ^8 33 34 1 35 36^

For the 2007 PDC as the previous, the Expert Panel included representatives from other stakeholder societies including the ASBMR, the International Society for Bone and Mineral Research (IBMS) and the National Osteoporosis Foundation (NOF). Topics considered included vertebral fracture assessment, technical and clinical issues relevant to dual-energy X-ray absorptiometry (DXA), and bone densitometry technologies other than central DXA. This is the first PDC that began using a more formal approach to formulating the questions, refinement of the statements and appropriateness ratings using the RAND/UCLA Appropriate Method (RAM) ^37^. From this and subsequent PDCs, positions were rated for appropriateness, necessity, and quality of evidence as defined below.

#### Appropriateness

Statements that the Expert Panel rated as ‘‘appropriate without disagreement’’ according to predefined criteria derived from the RAM were referred to the ISCD BOD with a recommendation to become ISCD Official Positions (see below). A statement was defined as ‘‘appropriate’’ when the expected health benefit exceeded the expected negative consequences by a significant margin such that it was worth performing.

#### Necessity

Recommended Official Positions that were rated by the Expert Panel were then rated according to necessity to perform in all circumstances (see below), i.e., whether the health benefits outweighed the risks to such an extent that it must be offered to all patients. Necessity rating was conducted in a similar fashion as the appropriateness rating, in that each Official Position had to be rated as necessary without disagreement using similar predefined RAM criteria.

#### Quality of evidence

Good: Evidence includes consistent results from well-designed well-conducted studies in representative populations.

Fair: Evidence is sufficient to determine effects on outcomes, but the strength of the evidence is limited by the number, quality, or consistency of the individual studies.

Poor: Evidence is insufficient to assess the effects on outcomes because of limited number or power of studies, important flaws in their design or conduct, gaps in the chain of evidence, or lack of information.

#### Strength of recommendations

A: Strong recommendation supported by the evidence

B: Recommendation supported by the evidence

C: Recommendation supported primarily by expert opinion

#### Application of recommendations

W: Worldwide recommendation

L: Application of recommendation may vary according to local requirements

### 2007 Pediatric PDC: Montreal, Quebec, Canada (2007) ^9 38 39 40 41^

In 2007, ISCD convened its first Pediatric PDC to address issues specific to the assessment of skeletal health in children as young as 5 years, adolescents and adults below 20 years old since they were not explicitly included in previous positions. For this PDC, the Expert Panel included representatives of the ASBMR and IBMS. Before this pediatric conference, the positions developed were only directly applicable for adults without consideration of the appropriateness for children. Topics considered were included DXA prediction of fracture and definition of pediatric osteoporosis; DXA assessment in diseases that may affect the pediatric skeleton; DXA interpretation and reporting; and peripheral quantitative computed tomography measurement.

### 2010 PDC: Bucharest, Romania (November 14, 2010) ^10 42 43 44 45 46 47 48 49 50 51 52^

This PDC was jointly sponsored by the ISCD and the IOF to clarify a number of important issues pertaining to the interpretation and implementation of FRAX® in clinical practice. FRAX® (http://www.shef.ac.uk/FRAX) is a simple computer-based tool that integrates clinical information and femoral neck BMD as an option to predict the 10-year probability of major osteoporotic fracture and hip fracture ^53^. It has become the defacto fracture risk predictor used word-wide since it was developed from studying population-based cohorts from Europe, North America, Asia and Australia. The FRAX® algorithms give the 10-year probability of fracture. The output is a 10-year probability of hip fracture and the 10-year probability of a major osteoporotic fracture (clinical spine, forearm, hip or shoulder fracture). Yet, at the time of this PDC, the ISCD and IOF had no positions on how it should be incorporated into clinical practice.

### 2013 PDC: March 21–March 23, 2013 in Tampa, FL, USA ^11 12 54 55 56 57 58 59^

There have been many scientific advances in measurement of fat and lean body mass as determined by dual-energy X-ray absorptiometry (DXA). Previously, no guidelines to the use of DXA for body composition existed. The recommendations pertain to clinically relevant issues regarding DXA indications of use, acquisition, analysis, quality control, interpretation, and reporting were addressed. In addition, indications for DXA and vertebral fracture assessment and use of reference data to calculate bone mineral density T-scores were also updated. The Expert Panel included representatives of the IOF, the ASBMR, the NOF, Osteoporosis Canada, and the North American Menopause Society. The Task Forces included participants from 6 countries and a variety of interests including academic institutions, private clinics, and industry.

### 2013 Pediatric PDC: Baltimore, MD USA (October 2-3, 2013) ^13 60 61 62 63 64^

It had been 6 years since the last Pediatric PDC. The conference was co-sponsored by the ASBMR with the aim to focus on advances in the field since that initial conference that would lead to revisions of the original positions. The previous topics were revisited with the addition of recommendations for infants and young children.

### 2015 PDC: Chicago, IL, USA (February 26–28, 2015) ^14 65 66 67 68 69 70^

There have been many scientific advances in fracture risk prediction beyond bone density. This PDC was convened to address the use of beyond the measurement of bone mineral density for fracture risk assessment, including Trabecular Bone Score (TBS) and hip geometry measures. Previously, no guidelines for nonbone mineral density DXA measures existed. Furthermore, there had been advances in the analysis of quantitative computed tomography (QCT) and ISCD had no comprehensive positions on QCT. Task forces were asked to address these advances that included finite element analysis, QCT of the hip, DXA-equivalent hip measurements, and opportunistic screening.

### 2019 PDC: Kuala Lumpur, Malaysia (March 20-23, 2019) ^15 71 72 73 74 75 76 77 78 79^

This PDC was convened to both revisit previous positions including monitoring bone density change with DXA, DXA machine cross-calibration, and pediatric bone health, but also novel questions in smaller subspecialty areas where the ISCD had to date been silent including spinal cord injury, periprosthetic and orthopedic bone health, and transgender medicine.

## ADULT POSITIONS BY CATEGORY

The adult positions are organized in a similar order and format as presented in the 2019 version of the Official Positions for Adults, Pediatrics and FRAX on the ISCD website and in the associated PDF version. In total, there are 43 categories.

### 1. Indications for Bone Mineral Density (BMD) Testing

Indications for BMD testing were first established at the 2003 PDC. Men under 70 years of age and women during the menopausal transition was added to the 2003 positions and both were then modified again at the 2013 PDC.

Position A1 regarding women 65 years and older wasn’t created in its current form until the 2003 ^25^. It was created *defacto* at the 2001 PDC when the statement was made in Binkley et al. ^18^ that Caucasian and Non-Caucasian women should have the same indications, *“The ISCD position is to apply existing indications for bone mass measurement in postmenopausal Caucasian women to non-Caucasian women. Thus, bone mass measurement is indicated for the following postmenopausal non-Caucasian women:*

1. *Those over age 65*.
2. *Those under age 65 with one or more risk factors*.
3. *Those with a prior fragility fracture*
4. *Those for whom knowledge of BMD would affect osteoporosis prevention/treatment decisions*.*”*

This was resolved to make official positions for all race women at the 2005 PDC.

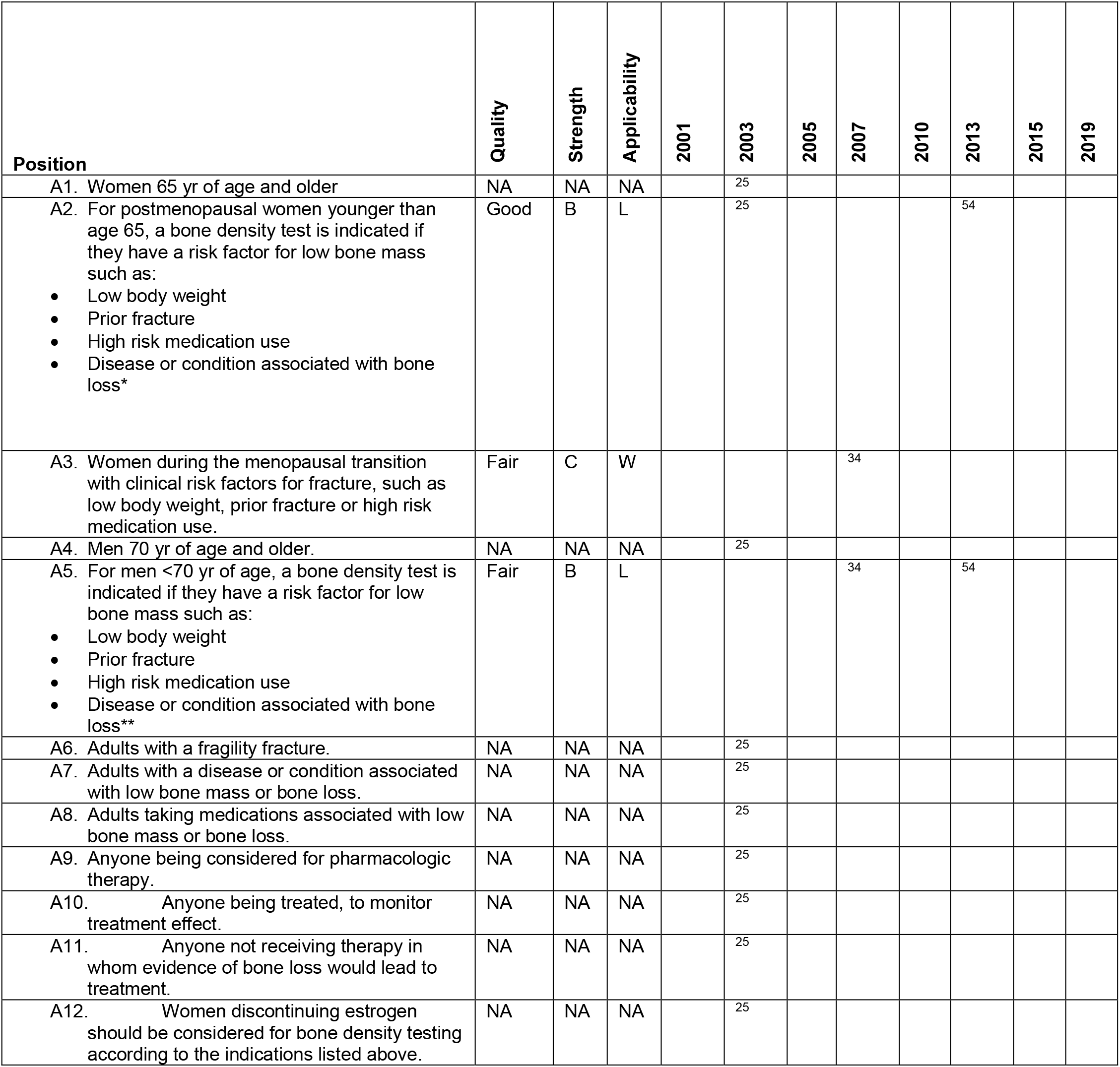
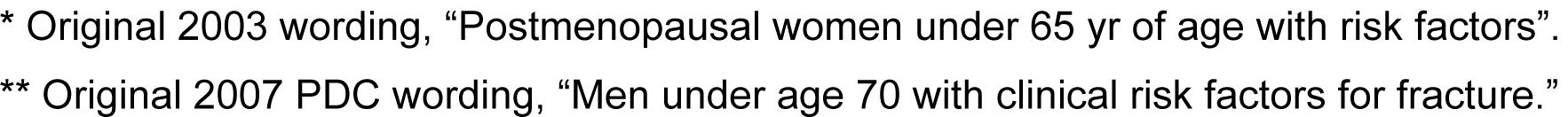

| Position | Quality | Strength | Applicability | 2001 | 2003 | 2005 | 2007 | 2010 | 2013 | 2015 | 2019 |
| --- | --- | --- | --- | --- | --- | --- | --- | --- | --- | --- | --- |
| A1. Women 65 yr of age and older | NA | NA | NA |  | <sup>25</sup> |  |  |  |  |  |  |
| A2. For postmenopausal women younger than age 65, a bone density test is indicated if they have a risk factor for low bone mass such as:<br>• Low body weight<br>• Prior fracture<br>• High risk medication use<br>• Disease or condition associated with bone loss* | Good | B | L |  | <sup>25</sup> |  |  |  | <sup>54</sup> |  |  |
| A3. Women during the menopausal transition with clinical risk factors for fracture, such as low body weight, prior fracture or high risk medication use. | Fair | C | W |  |  |  | <sup>34</sup> |  |  |  |  |
| A4. Men 70 yr of age and older. | NA | NA | NA |  | <sup>25</sup> |  |  |  |  |  |  |
| A5. For men <70 yr of age, a bone density test is indicated if they have a risk factor for low bone mass such as:<br>• Low body weight<br>• Prior fracture<br>• High risk medication use<br>• Disease or condition associated with bone loss** | Fair | B | L |  |  |  | <sup>34</sup> |  | <sup>54</sup> |  |  |
| A6. Adults with a fragility fracture. | NA | NA | NA |  | <sup>25</sup> |  |  |  |  |  |  |
| A7. Adults with a disease or condition associated with low bone mass or bone loss. | NA | NA | NA |  | <sup>25</sup> |  |  |  |  |  |  |
| A8. Adults taking medications associated with low bone mass or bone loss. | NA | NA | NA |  | <sup>25</sup> |  |  |  |  |  |  |
| A9. Anyone being considered for pharmacologic therapy. | NA | NA | NA |  | <sup>25</sup> |  |  |  |  |  |  |
| A10. Anyone being treated, to monitor treatment effect. | NA | NA | NA |  | <sup>25</sup> |  |  |  |  |  |  |
| A11. Anyone not receiving therapy in whom evidence of bone loss would lead to treatment. | NA | NA | NA |  | <sup>25</sup> |  |  |  |  |  |  |
| A12. Women discontinuing estrogen should be considered for bone density testing according to the indications listed above. | NA | NA | NA |  | <sup>25</sup> |  |  |  |  |  |  |

\* Original 2003 wording, “Postmenopausal women under 65 yr of age with risk factors”.
\*\* Original 2007 PDC wording, “Men under age 70 with clinical risk factors for fracture.”

### 2. Reference Database for T-Scores

The use and calculation of T-scores has been addressed since 2001 at four separate PDCs. At the 2001 PDC, “there was insufficient data to definitively conclude that men and women fracture at the same DXA-measured BMD” ^5^. The question of what reference data to use for men and non-Caucasian populations has been examine multiple times since the first PDC. However, standardization to Caucasian females from the NHANES III for Femur Neck would not occur until 2005. Before that time, manufacturer specific reference data was common. In 2013, the conference examined if NHANES III should be used for spine as well. Ultimately, this proposal was turned down. See position A16.

| Position | Quality | Strength | Applicability | 2001 | 2003 | 2005 | 2007 | 2010 | 2013 | 2015 | 2019 |
| --- | --- | --- | --- | --- | --- | --- | --- | --- | --- | --- | --- |
| A13. Use a uniform Caucasian (nonrace adjusted) female normative database for **women of all ethnic groups**.<br>*Note: Application of recommendation may vary according to local requirements.* | NA | NA | NA | 18 | 23 |  |  |  |  |  |  |
| A14. Use a uniform Caucasian (nonrace adjusted) female reference for **men** of all ethnic groups. **<br>*\*Note: Application of recommendation may vary according to local requirements.* | Fair | B | L | 18 | 23 |  |  |  | 55 |  |  |
| A15. Manufacturers should continue to use NHANES III data as the reference standard for femoral neck and total hip T-scores. | Poor | C | W |  |  | 29 |  |  | 55 |  |  |
| A16. Manufacturers should continue to use their own databases for the lumbar spine as the reference standard for T-scores. | Poor | C | W |  |  |  |  |  | 55 |  |  |
| A17. If local reference data are available they should be used to calculate only Z-scores but not T-scores. | Poor | C | W |  |  |  |  |  | 55 |  |  |

\*\* In 2001, the position stated, “a male normative database should continue to be used for central DXA diagnosis of osteoporosis in men”<sup>5</sup> due to insufficient evidence to the contrary. In 2003, this was updated to, “Use a uniform white (nonrace adjusted) male normative database for men of all ethnic groups. This applies to the US population.” In 2005. The wording of the position was informally changed from “white” to “Caucasian”<sup>7</sup>.

### 3. Central DXA for Diagnosis

All of the positions in this section were finalized by the end of the 2005 PDC. However, there were critical updates to what sites were deemed valid for diagnosing osteoporosis during that time. Originally, trochanter was thought of as a valid diagnostic site. But trochanter was dropped at the 2005 PDC and 1/3^rd^ distal femur was added if the spine and hip were both not valid. The current positions do not make mention which central DXA site is preferred for diagnosis and monitoring. However, the 2001 positions explicitly state that, “PA spine is the preferred site for monitoring. The total hip should be used when the PA spine is technically invalid.” These specific position statements were dropped in 2003.

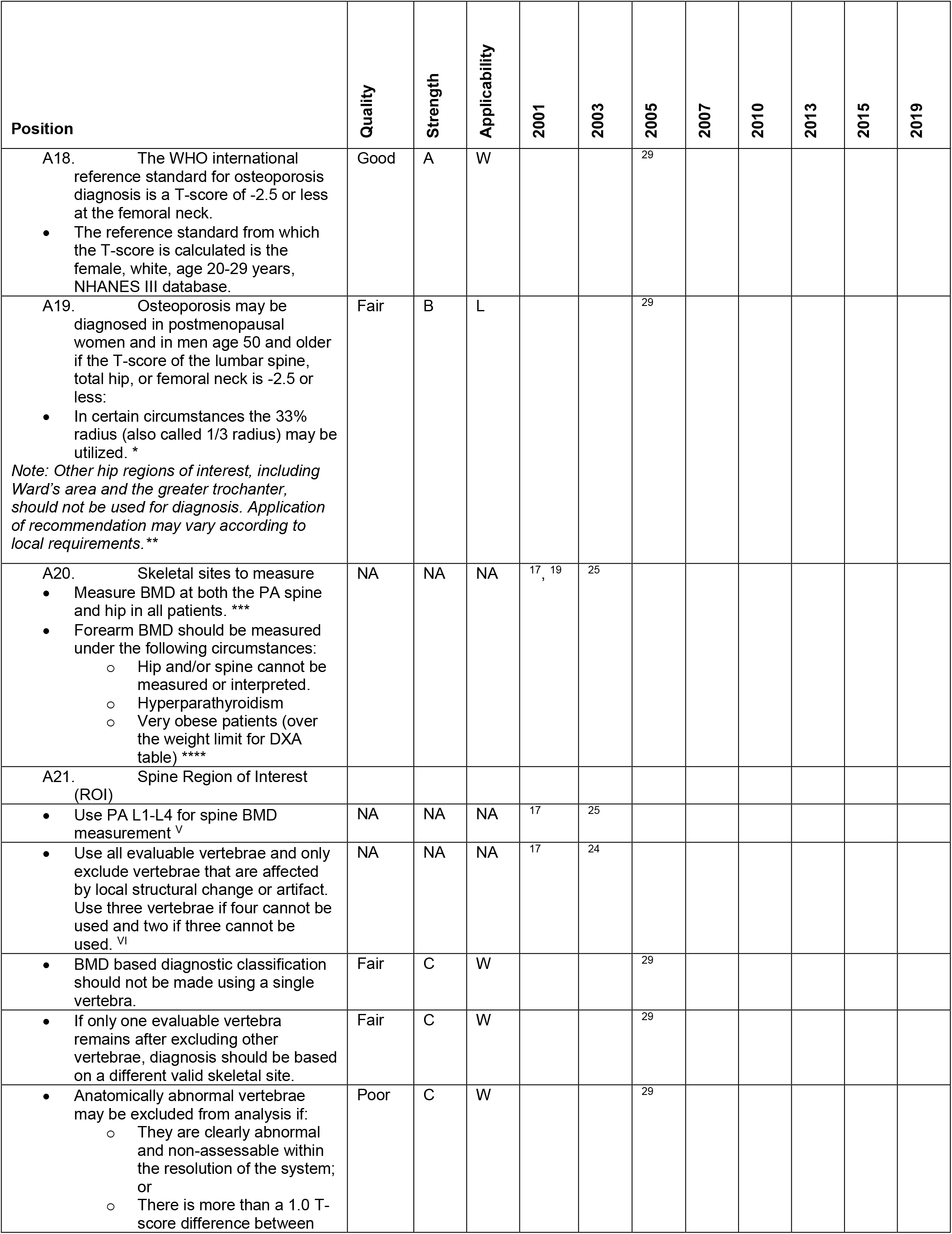
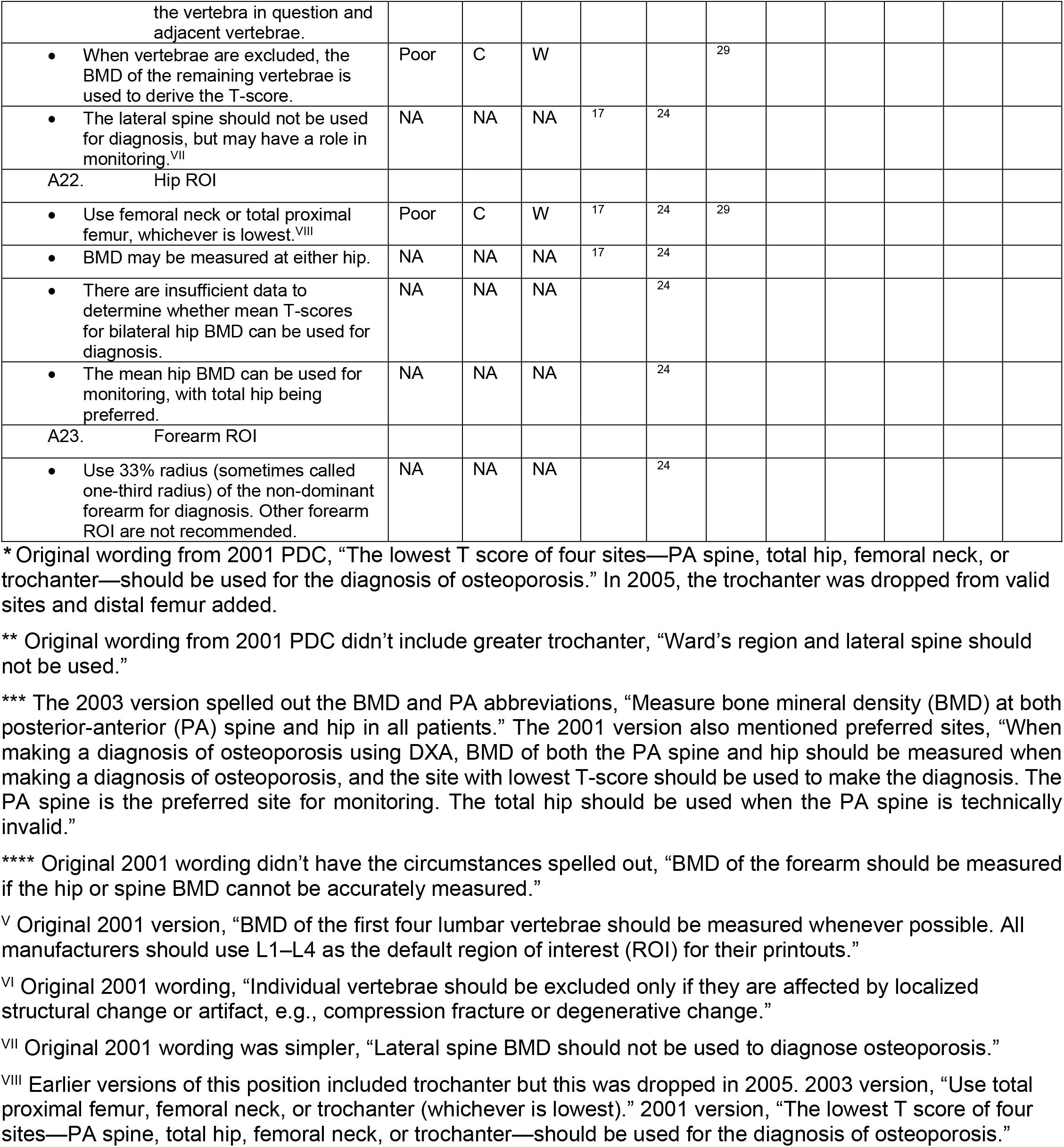

| Position | Quality | Strength | Applicability | 2001 | 2003 | 2005 | 2007 | 2010 | 2013 | 2015 | 2019 |
| --- | --- | --- | --- | --- | --- | --- | --- | --- | --- | --- | --- |
| A18. The WHO international reference standard for osteoporosis diagnosis is a T-score of -2.5 or less at the femoral neck.<br>The reference standard from which the T-score is calculated is the female, white, age 20-29 years, NHANES III database. | Good | A | W |  |  | 29 |  |  |  |  |  |
| A19. Osteoporosis may be diagnosed in postmenopausal women and in men age 50 and older if the T-score of the lumbar spine, total hip, or femoral neck is -2.5 or less:<br>In certain circumstances the 33% radius (also called 1/3 radius) may be utilized. *<br>*Note: Other hip regions of interest, including Ward's area and the greater trochanter, should not be used for diagnosis. Application of recommendation may vary according to local requirements.\*\** | Fair | B | L |  |  | 29 |  |  |  |  |  |
| A20. Skeletal sites to measure<br>Measure BMD at both the PA spine and hip in all patients. ***<br>Forearm BMD should be measured under the following circumstances:<br>Hip and/or spine cannot be measured or interpreted.<br>Hyperparathyroidism<br>Very obese patients (over the weight limit for DXA table) **** | NA | NA | NA | 17, 19 | 25 |  |  |  |  |  |  |
| A21. Spine Region of Interest (ROI) |  |  |  |  |  |  |  |  |  |  |  |
| Use PA L1-L4 for spine BMD measurement <sup>v</sup> | NA | NA | NA | 17 | 25 |  |  |  |  |  |  |
| Use all evaluable vertebrae and only exclude vertebrae that are affected by local structural change or artifact. Use three vertebrae if four cannot be used and two if three cannot be used. <sup>vi</sup> | NA | NA | NA | 17 | 24 |  |  |  |  |  |  |
| BMD based diagnostic classification should not be made using a single vertebra. | Fair | C | W |  |  | 29 |  |  |  |  |  |
| If only one evaluable vertebra remains after excluding other vertebrae, diagnosis should be based on a different valid skeletal site. | Fair | C | W |  |  | 29 |  |  |  |  |  |
| Anatomically abnormal vertebrae may be excluded from analysis if:<br>They are clearly abnormal and non-assessable within the resolution of the system; or<br>There is more than a 1.0 T-score difference between | Poor | C | W |  |  | 29 |  |  |  |  |  |

|  |  |  |  |  |  |  |
| --- | --- | --- | --- | --- | --- | --- |
| the vertebra in question and adjacent vertebrae. |  |  |  |  |  |  |
| When vertebrae are excluded, the BMD of the remaining vertebrae is used to derive the T-score. | Poor | C | W |  |  | 29 |
| The lateral spine should not be used for diagnosis, but may have a role in monitoring.<sup>VII</sup> | NA | NA | NA | 17 | 24 |  |
| A22. Hip ROI |  |  |  |  |  |  |
| Use femoral neck or total proximal femur, whichever is lowest.<sup>VIII</sup> | Poor | C | W | 17 | 24 | 29 |
| BMD may be measured at either hip. | NA | NA | NA | 17 | 24 |  |
| There are insufficient data to determine whether mean T-scores for bilateral hip BMD can be used for diagnosis. | NA | NA | NA |  | 24 |  |
| The mean hip BMD can be used for monitoring, with total hip being preferred. | NA | NA | NA |  | 24 |  |
| A23. Forearm ROI |  |  |  |  |  |  |
| Use 33% radius (sometimes called one-third radius) of the non-dominant forearm for diagnosis. Other forearm ROI are not recommended. | NA | NA | NA |  | 24 |  |

\* Original wording from 2001 PDC, “The lowest T score of four sites—PA spine, total hip, femoral neck, or trochanter—should be used for the diagnosis of osteoporosis.” In 2005, the trochanter was dropped from valid sites and distal femur added.
\*\* Original wording from 2001 PDC didn’t include greater trochanter, “Ward’s region and lateral spine should not be used.”
\*\*\* The 2003 version spelled out the BMD and PA abbreviations, “Measure bone mineral density (BMD) at both posterior-anterior (PA) spine and hip in all patients.” The 2001 version also mentioned preferred sites, “When making a diagnosis of osteoporosis using DXA, BMD of both the PA spine and hip should be measured when making a diagnosis of osteoporosis, and the site with lowest T-score should be used to make the diagnosis. The PA spine is the preferred site for monitoring. The total hip should be used when the PA spine is technically invalid.”
\*\*\*\* Original 2001 wording didn’t have the circumstances spelled out, “BMD of the forearm should be measured if the hip or spine BMD cannot be accurately measured.”
<sup>V</sup> Original 2001 version, “BMD of the first four lumbar vertebrae should be measured whenever possible. All manufacturers should use L1–L4 as the default region of interest (ROI) for their printouts.”
<sup>VI</sup> Original 2001 wording, “Individual vertebrae should be excluded only if they are affected by localized structural change or artifact, e.g., compression fracture or degenerative change.”
<sup>VII</sup> Original 2001 wording was simpler, “Lateral spine BMD should not be used to diagnose osteoporosis.”
<sup>VIII</sup> Earlier versions of this position included trochanter but this was dropped in 2005. 2003 version, “Use total proximal femur, femoral neck, or trochanter (whichever is lowest).” 2001 version, “The lowest T score of four sites—PA spine, total hip, femoral neck, or trochanter—should be used for the diagnosis of osteoporosis.”

### 4. Fracture Assessment

The presentation of these two positions in abstract of the controversies going on in the field obscures their meaning. A24 is stating that BMD can be used for both diagnostic classification of osteoporosis and quantifying fracture risk in fracture models. From the discussion in Hans ^29^, this was an important distinction at the time since diagnosis was being consolidated around one DXA measurement site, the femur neck, one reference database, NHANES III, and one T-score values of – 2.5. Simultaneously, it was being recognized that other technologies and regions of interests were still clinically important for other purposes, like monitoring change over time and quantifying fracture risk. These positions were trying to convey that it is OK to use one specific measurement for diagnosis and others for fracture risk. In the subsequent years, alternative technologies did not fair well in the marketplace, such as peripheral DXA, heel ultrasound, etc. and these distinctions became less important. But there is renewed interests in alternative technologies to DXA where these positions may again prove to be useful.

| Position | Quality | Strength | Applicability | 2001 | 2003 | 2005 | 2007 | 2010 | 2013 | 2015 | 2019 |
| --- | --- | --- | --- | --- | --- | --- | --- | --- | --- | --- | --- |
| A24. A distinction is made between diagnostic classification and the use of BMD for fracture risk assessment. | Good | A | W |  |  | <sup>29</sup> |  |  |  |  |  |
| A25. For fracture risk assessment, any well-validated technique can be used, including measurements of more than 1 site where this has been shown to improve the assessment of risk. | Good | A | W |  |  | <sup>29</sup> |  |  |  |  |  |

### 5. Use of the Term “Osteopenia”

These positions were created to encourage the use of “low bone mass” over osteopenia for interpreting T-score values. Osteopenia was often being thought of as a disease and ISCD wanted to discourage this. A full discussion can be found in Leslie et al. ^30^.

| Position | Quality | Strength | Applicability | 2001 | 2003 | 2005 | 2007 | 2010 | 2013 | 2015 | 2019 |
| --- | --- | --- | --- | --- | --- | --- | --- | --- | --- | --- | --- |
| A26. The term “osteopenia” is retained, but “low bone mass” or “low bone density” is preferred. | Poor | C | W |  |  | <sup>30</sup> |  |  |  |  |  |
| A27. People with low bone mass or density are not necessarily at high fracture risk. | Poor | C | W |  |  | <sup>30</sup> |  |  |  |  |  |

### 6. BMD Reporting in Postmenopausal Women and Men Age 50 and Older

These positions were to clarify the use of T- and Z-scores based on age and menopausal status, and the use of the WHO definitions of normal, osteopenia, osteoporosis, and established osteoporosis. Guidance on the specifics of how the T- and Z-score was to be calculated with respect to reference populations and race were covered in later positions.

| Position | Quality | Strength | Applicability | 2001 | 2003 | 2005 | 2007 | 2010 | 2013 | 2015 | 2019 |
| --- | --- | --- | --- | --- | --- | --- | --- | --- | --- | --- | --- |
| A28. T-scores are preferred. | Fair | B | W |  |  | <sup>30</sup> |  |  |  |  |  |
| A29. The WHO densitometric classification is applicable. | Fair | B | W |  |  | <sup>30</sup> |  |  |  |  |  |

### 7. BMD Reporting in Females Prior to Menopause and in Males Younger than Age 50

| Position | Quality | Strength | Applicability | 2001 | 2003 | 2005 | 2007 | 2010 | 2013 | 2015 | 2019 |
| --- | --- | --- | --- | --- | --- | --- | --- | --- | --- | --- | --- |
| A30. Z-scores, not T-scores, are preferred. This is particularly important in children. | Poor | C | W |  |  | 30 |  |  |  |  |  |
| A31. A Z-score of -2.0 or lower is defined as “below the expected range for age”, and a Z-score above -2.0 is “within the expected range for age.” | Poor | C | W |  | 23 | 30 |  |  |  |  |  |
| A32. Osteoporosis cannot be diagnosed in men under age 50 on the basis of BMD alone. | Fair | B | W |  | 23 |  | 34 |  |  |  |  |
| A33. The WHO diagnostic criteria may be applied to women in the menopausal transition. | Fair | B | W |  | 23 |  | 34 |  |  |  |  |

### 8. Z-Score Reference Database

| Position | Quality | Strength | Applicability | 2001 | 2003 | 2005 | 2007 | 2010 | 2013 | 2015 | 2019 |
| --- | --- | --- | --- | --- | --- | --- | --- | --- | --- | --- | --- |
| A34. Z-scores should be population specific where adequate reference data exist. For the purpose of Z-score calculation, the patient's self-reported ethnicity should be used. | Poor | C | W |  |  | 30 |  |  |  |  |  |

### 9. Serial BMD Measurements

| Position | Quality | Strength | Applicability | 2001 | 2003 | 2005 | 2007 | 2010 | 2013 | 2015 | 2019 |
| --- | --- | --- | --- | --- | --- | --- | --- | --- | --- | --- | --- |
| A35. Serial BMD testing, in combination with clinical assessment of fracture risk, bone turnover markers, and other factors including height loss and TBS, can be used to determine whether treatment should be initiated in untreated patients, according to locally applicable guidelines.* | Good | A | W | 19 | 25 |  |  |  |  |  | 73 |
| A36. Serial BMD testing can monitor response to therapy by finding an increase or stability of bone density. | NA | NA | NA | 19 | 25 |  |  |  |  |  |  |
| A37. Serial BMD testing should be used to monitor individuals following cessation of osteoporosis pharmacologic therapy. | Good | B | W |  |  |  |  |  |  |  | 73 |
| A38. Serial BMD testing can detect loss of bone density, indicating the need for assessment of treatment adherence, evaluation of secondary causes of osteoporosis, and re-evaluation of treatment options. | Good | B | W | 19 | 25 |  |  |  |  |  | 73 |
| A39. Follow-up BMD testing should be done when the results are likely to influence patient management. | Good | A | W |  | 25 |  |  |  |  |  | 73 |
| A40. Intervals between BMD testing should be determined according to each patient's clinical status: typically 1 yr after initiation or change of therapy is appropriate, with longer intervals once therapeutic effect is established. | NA | NA | NA | 19 | 25 |  |  |  |  |  |  |
| A41. In conditions associated with rapid bone loss, such as glucocorticoid therapy, testing more frequently is appropriate. | NA | NA | NA |  | 25 |  |  |  |  |  |  |

\* From 2003, "Serial BMD testing can be used to determine whether treatment should be started on untreated patients, because significant loss may be an indication for treatment." From 2001, "Subjects not on treatment can be monitored to determine if and when significant bone loss occurs."

### 10. Phantom Scanning and Calibration

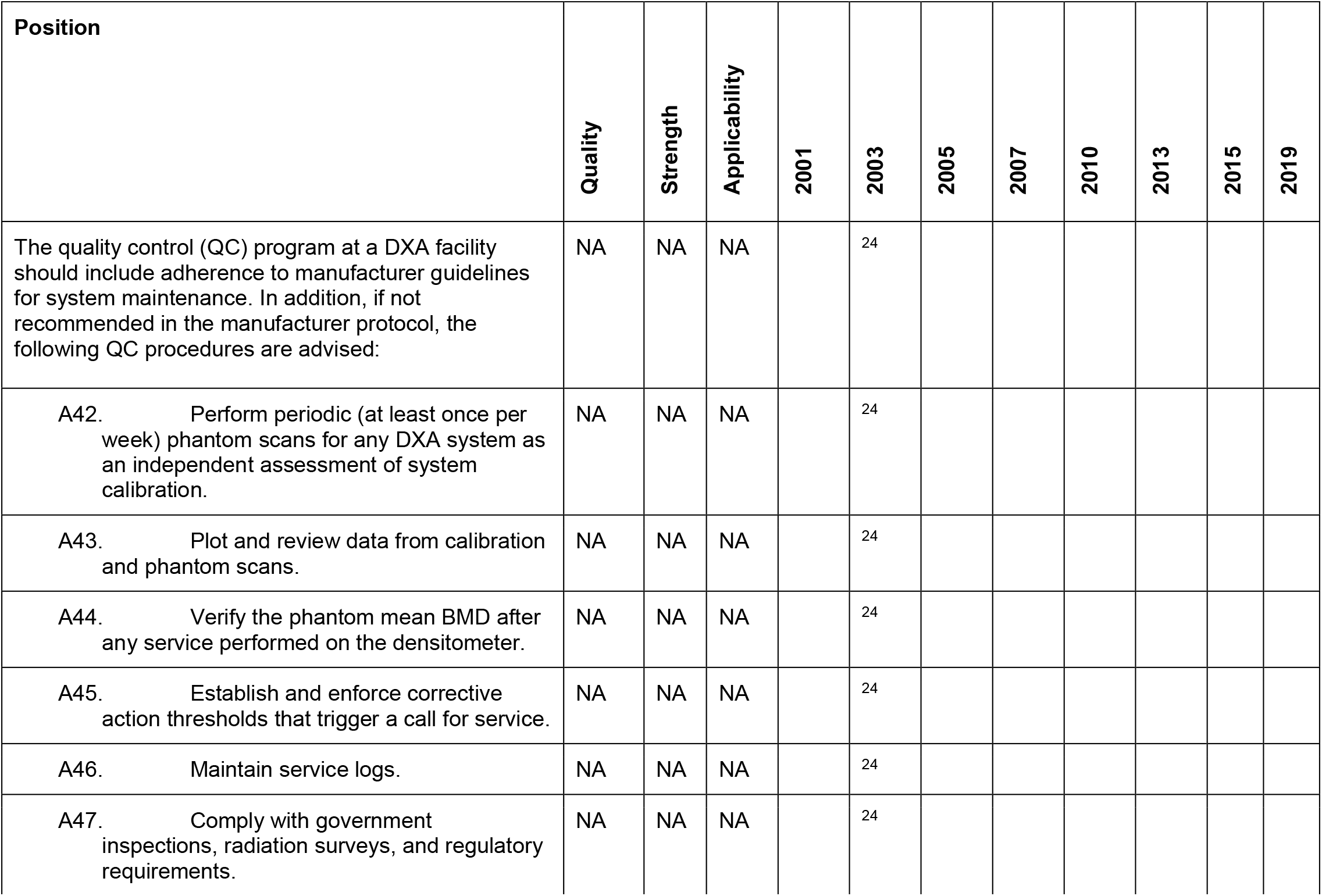

### 11. Precision Assessment

| Position | Quality | Strength | Applicability | 2001 | 2003 | 2005 | 2007 | 2010 | 2013 | 2015 | 2019 |
| --- | --- | --- | --- | --- | --- | --- | --- | --- | --- | --- | --- |
| A48. Each DXA facility should determine its precision error and calculate the least significant change (LSC). | NA | NA | NA |  | 24 |  |  |  |  |  |  |
| A49. The precision error supplied by the manufacturer should not be used. | NA | NA | NA |  |  |  |  |  |  |  |  |
| A50. If a DXA facility has more than 1 technologist, an average precision error combining data from all technologists should be used to establish precision error and LSC for the facility, provided the precision error for each technologist is within a pre-established range of acceptable performance. | NA | NA | NA |  | 24 |  |  |  |  |  |  |
| A51. Every technologist should perform an in vivo precision assessment using patients representative of the clinic's patient population. | NA | NA | NA |  | 24 |  |  |  |  |  |  |
| A52. Each technologist should do 1 complete precision assessment after basic scanning skills have been learned (eg, manufacturer training) and after having performed approximately 100 patient-scans. | NA | NA | NA |  | 24 |  |  |  |  |  |  |
| A53. A repeat precision assessment should be done if a new DXA system is installed. | NA | NA | NA |  | 24 |  |  |  |  |  |  |
| A54. A repeat precision assessment should be done if a technologist's skill level has changed. | NA | NA | NA | 19 | 24 |  |  |  |  |  |  |
| A55. To perform a precision analysis:<br>• Measure 15 patients 3 times, or 30 patients 2 times, repositioning the patient after each scan.<br>• Calculate the root mean square standard deviation (RMS-SD) for the group.<br>• Calculate LSC for the group at 95% confidence interval. | NA | NA | NA |  | 24 |  |  |  |  |  |  |
| A56. The minimum acceptable precision for an individual technologist is:<br>• Lumbar Spine: 1.9% (LSC=5.3%)<br>• Total Hip: 1.8% (LSC=5.0%)<br>• Femoral Neck: 2.5% (LSC=6.9%)<br>• Retraining is required if a technologist's precision is worse than these values. | Good | B | W |  |  | 31 |  |  |  |  |  |
| A57. Precision assessment should be standard clinical practice. Precision assessment is not research and may potentially benefit patients. It should not require approval of an institutional review | NA | NA | NA |  | 24 |  |  |  |  |  |  |
| board. Adherence to local radiologic safety regulations is necessary. Performance of a precision assessment requires the consent of participating patients. |  |  |  |  |  |  |  |  |  |  |  |

### 12. Cross Calibration of DXA: Changing Hardware or Systems

| Position | Quality | Strength | Applicability | 2001 | 2003 | 2005 | 2007 | 2010 | 2013 | 2015 | 2019 |
| --- | --- | --- | --- | --- | --- | --- | --- | --- | --- | --- | --- |
| A58. When changing hardware, but not the entire system, or when replacing a system with the same technology (manufacturer and model), cross-calibration should be performed by having one technologist do 10 phantom scans, with repositioning, before and after a hardware change.<br>If a greater than 1% difference in mean BMD is observed, contact the manufacturer for service/correction. | Good | A | W |  | 24 | 31 |  |  |  |  |  |
| A59. When changing an entire system to one made by the same manufacturer using a different technology, or when changing to a system made by a different manufacturer, one approach to cross-calibration is:<br>Scan 30 patients representative of the facility's patient population once on the initial system and then twice on the new system within 60 days.<br>Measure those anatomic sites commonly measured in clinical practice, typically spine and proximal femur.<br>Facilities must comply with locally applicable regulations regarding DXA.<br>Calculate the average BMD relationship and LSC between the initial and new machine using the ISCD DXA Machine Cross-Calibration Tool (www.ISCD.org).<br>Use this LSC for comparison between the previous and new system. Inter-system quantitative comparisons can only be made if cross-calibration is performed on each skeletal site commonly measured.<br>Once a new precision assessment has been performed on the new system, all future scans should be compared to scans performed on the new system using the newly established intra-system LSC. | Good | A | W |  |  | 31 |  |  |  |  |  |

### 13. Cross Calibration of DXA: Adding Hardware or Systems

With lower reimbursement for DXA scanning at independent clinics in the US versus DXA scanning that takes place in medical centers ^80^, there has been a consolidation of DXA systems away from single system sites, to sites with multiple systems. The 2005 position was written in a language that implies that the need for cross-calibration was most likely due to replacing an older system with a new one. The 2019 positions are worded to address the needs of multi-system sites.

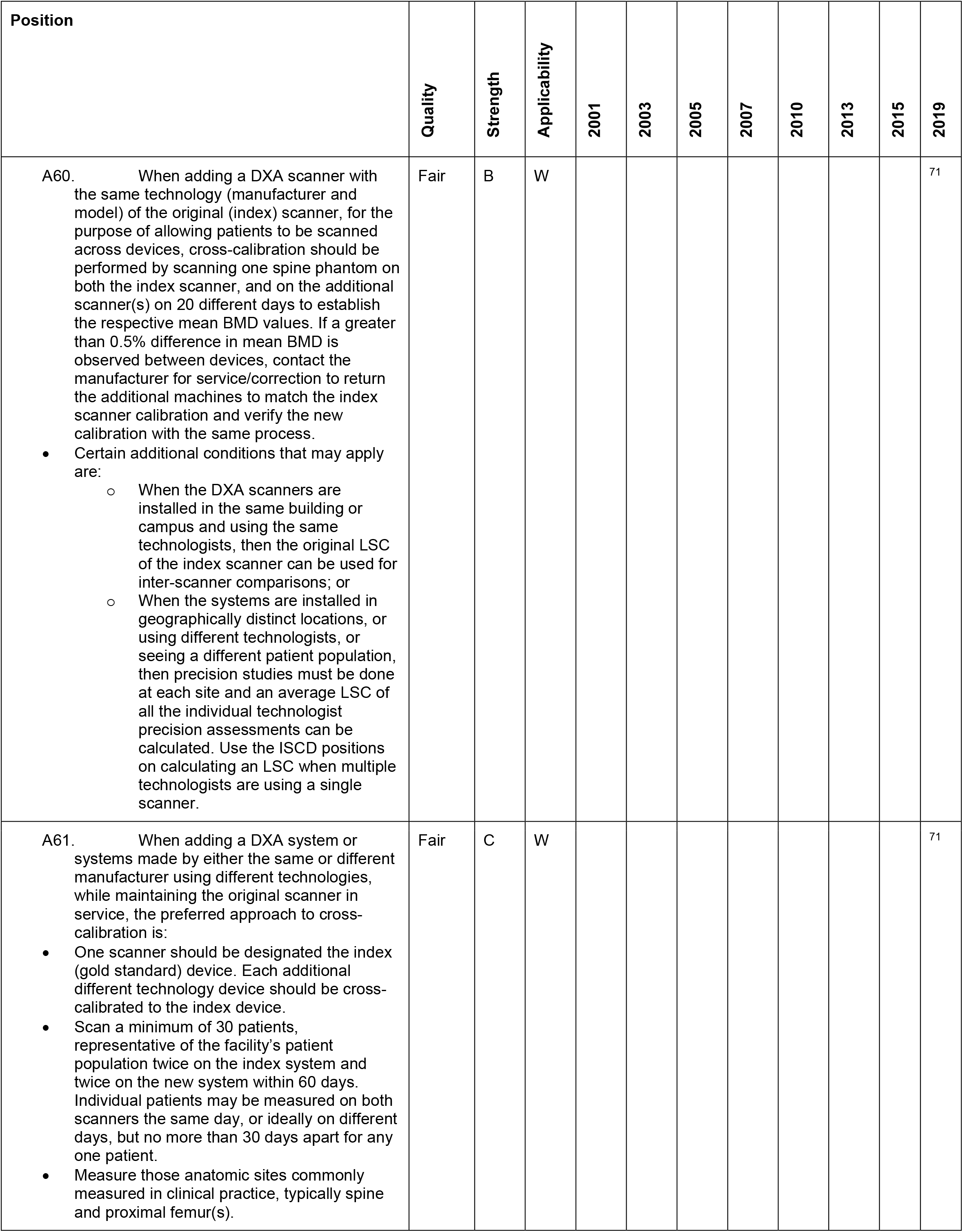
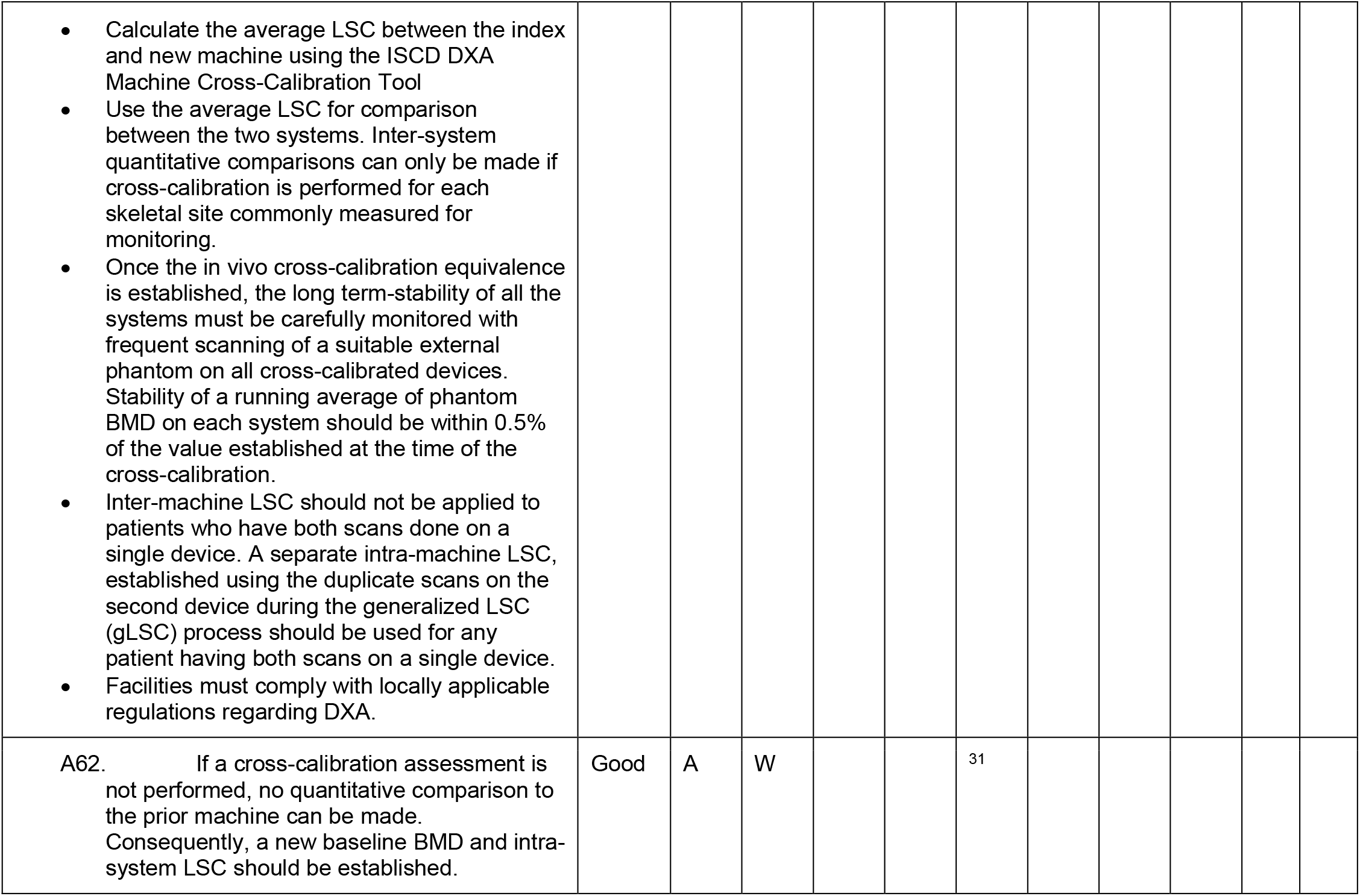

| Position | Quality | Strength | Applicability | 2001 | 2003 | 2005 | 2007 | 2010 | 2013 | 2015 | 2019 |
| --- | --- | --- | --- | --- | --- | --- | --- | --- | --- | --- | --- |
| A60. When adding a DXA scanner with the same technology (manufacturer and model) of the original (index) scanner, for the purpose of allowing patients to be scanned across devices, cross-calibration should be performed by scanning one spine phantom on both the index scanner, and on the additional scanner(s) on 20 different days to establish the respective mean BMD values. If a greater than 0.5% difference in mean BMD is observed between devices, contact the manufacturer for service/correction to return the additional machines to match the index scanner calibration and verify the new calibration with the same process.<br>Certain additional conditions that may apply are:<br>When the DXA scanners are installed in the same building or campus and using the same technologists, then the original LSC of the index scanner can be used for inter-scanner comparisons; or<br>When the systems are installed in geographically distinct locations, or using different technologists, or seeing a different patient population, then precision studies must be done at each site and an average LSC of all the individual technologist precision assessments can be calculated. Use the ISCD positions on calculating an LSC when multiple technologists are using a single scanner. | Fair | B | W |  |  |  |  |  |  |  | 71 |
| A61. When adding a DXA system or systems made by either the same or different manufacturer using different technologies, while maintaining the original scanner in service, the preferred approach to cross-calibration is:<br>One scanner should be designated the index (gold standard) device. Each additional different technology device should be cross-calibrated to the index device.<br>Scan a minimum of 30 patients, representative of the facility's patient population twice on the index system and twice on the new system within 60 days. Individual patients may be measured on both scanners the same day, or ideally on different days, but no more than 30 days apart for any one patient.<br>Measure those anatomic sites commonly measured in clinical practice, typically spine and proximal femur(s). | Fair | C | W |  |  |  |  |  |  |  | 71 |

|  |  |  |  |  |  |  |
| --- | --- | --- | --- | --- | --- | --- |
| Calculate the average LSC between the index and new machine using the ISCD DXA Machine Cross-Calibration Tool<br>Use the average LSC for comparison between the two systems. Inter-system quantitative comparisons can only be made if cross-calibration is performed for each skeletal site commonly measured for monitoring.<br>Once the in vivo cross-calibration equivalence is established, the long term-stability of all the systems must be carefully monitored with frequent scanning of a suitable external phantom on all cross-calibrated devices. Stability of a running average of phantom BMD on each system should be within 0.5% of the value established at the time of the cross-calibration.<br>Inter-machine LSC should not be applied to patients who have both scans done on a single device. A separate intra-machine LSC, established using the duplicate scans on the second device during the generalized LSC (gLSC) process should be used for any patient having both scans on a single device.<br>Facilities must comply with locally applicable regulations regarding DXA. |  |  |  |  |  |  |
| A62. If a cross-calibration assessment is not performed, no quantitative comparison to the prior machine can be made. Consequently, a new baseline BMD and intra-system LSC should be established. | Good | A | W |  |  | 31 |

### 14. BMD Comparison Between Facilities

Although DXA quality control is a foundational principle for the ISCD, the comparison between facilities has only generated two positions. Since it is common for patients to move or to change health providers, this category of positions is of high interest. Yet, the positions themselves effectively state that DXA scans cannot be quantitatively and temporally followed if the patient is seen at more than one clinic. The author sees this as a failing in scope for the ISCD since this situation is so common.

| Position | Quality | Strength | Applicability | 2001 | 2003 | 2005 | 2007 | 2010 | 2013 | 2015 | 2019 |
| --- | --- | --- | --- | --- | --- | --- | --- | --- | --- | --- | --- |
| A63. It is not possible to quantitatively compare BMD or to calculate a LSC between facilities without cross-calibration. | Poor | C | W |  |  | 31 |  |  |  |  |  |
| A64. Patients should return to the same DXA device that was used to perform their most recent prior study, provided that the facility in vivo precision and LSC values are known and do not exceed established maximum values. | Fair | C | W |  |  |  |  |  |  |  | 71 |

### 15. Vertebral Fracture Assessment Nomenclature

Several other terms have previously been used for describing vertebral fractures from lateral DXA scans. Including morphometric X-ray absorptiometry (MXA), Instant Vertebral Assessment (IVA), Radiographic Vertebral Assessment (RVA), Lateral Vertebral Assessment (LVA), Dual-energy Vertebral Assessment (DVA) and others. Some of these terms were coined by specific DXA manufacturers. VFA was viewed by the expert panel of the 2005 PDC as a neutral term that the field could standardize around. See Vokes et al. ^32^ for the complete discussion.

| Position | Quality | Strength | Applicability | 2001 | 2003 | 2005 | 2007 | 2010 | 2013 | 2015 | 2019 |
| --- | --- | --- | --- | --- | --- | --- | --- | --- | --- | --- | --- |
| A65. Vertebral Fracture Assessment (VFA) is the correct term to denote densitometric spine imaging performed for the purpose of detecting vertebral fractures. | Poor | C | W |  |  | 32 |  |  |  |  |  |

### 16. Indications for VFA

The 2013 PDC greatly simplified the previous recommendations from 2005 and 2007 for greater ease of use and utilization by practitioners.

| Position | Quality | Strength | Applicability | 2001 | 2003 | 2005 | 2007 | 2010 | 2013 | 2015 | 2019 |
| --- | --- | --- | --- | --- | --- | --- | --- | --- | --- | --- | --- |
| A66. Lateral Spine imaging with Standard Radiography or Densitometric VFA is indicated when T-score is < -1.0 and of one or more of the following is present:<br>• Women age ≥ 70 years or men ≥ age 80 years<br>• Historical height loss > 4 cm (>1.5 inches)<br>• Self-reported but undocumented prior vertebral fracture<br>• Glucocorticoid therapy equivalent to ≥ 5 mg of prednisone or equivalent per day for ≥ 3 months* | Fair | B | W |  |  | 32 | 33 |  | 56 |  |  |

\* Original wording from 2005, "Consider VFA when the results may influence clinical management. When BMD measurement is indicated, performance of VFA should be considered in clinical situations that may be associated with vertebral fractures. Examples include: Documented height loss of greater than 2 cm (0.75 in) or historical height loss greater than 4 cm (1.5 in) since young adult; History of fracture after age 50; Commitment to long-term oral or parenteral glucocorticoid therapy; History and/or findings suggestive of vertebral fracture not documented by prior radiologic study."

Original wording from 2007, “Postmenopausal women with low bone mass (osteopenia) by BMD criteria, PLUS any one of the following: Age greater than or equal to 70 yr, Historical height loss greater than 4 cm (1.6 in), Prospective height loss greater than 2 cm (0.8 in), Self-reported vertebral fracture (not previously documented), Two or more of the following: - Age 60 to 69 yr, - Self-reported prior non-vertebral fracture - Historical height loss of 2 to 4 cm, - Chronic systemic diseases associated with increased risk of vertebral fractures (for example, moderate to severe chronic obstructive pulmonary disorder (COPD) or chronic obstructive airways disease (COAD), seropositive rheumatoid arthritis, Crohn’s disease) (Grade: Fair-B-W-Necessary). Men with low bone mass (osteopenia) by BMD criteria, PLUS any one of the following: - Age 80 yr or older, - Historical height loss greater than 6 cm (2.4 in), - Prospective height loss greater than 3 cm (1.2 in) - Self-reported vertebral fracture (not previously documented), - Two or more of the following; - Age 70 to 79 yr, - Self-reported prior non-vertebral fracture, - Historical height loss of 3 to 6 cm, - On pharmacologic androgen deprivation therapy or following orchietomy, - Chronic systemic diseases associated with increased risk of vertebral fractures (for example, moderate to severe COPD or COAD, seropositive rheumatoid arthritis, Crohn’s disease) (Grade: Fair-C-W). Women or men on chronic glucocorticoid therapy (equivalent to 5 mg or more of prednisone daily for 3 mo or longer) (Grade: Fair-B-W-Necessary). Postmenopausal women or men with osteoporosis by BMD criteria, if documentation of one or more vertebral fractures will alter clinical management. (Grade: Good-C-W-Necessary)”

### 17. Methods for Defining and Reporting Fractures on VFA

One position was modified in 2007 from the 2005 positions on defining and reporting VFA. The changes drew upon an extensive literature search found in ^33^ such that the recommendation to use the Genant semiquantitative method were strengthened.

| Position | Quality | Strength | Applicability | 2001 | 2003 | 2005 | 2007 | 2010 | 2013 | 2015 | 2019 |
| --- | --- | --- | --- | --- | --- | --- | --- | --- | --- | --- | --- |
| A67. The methodology utilized for vertebral fracture identification should be similar to standard radiological approaches and be provided in the report. | Fair | B | W |  |  | <sup>32</sup> |  |  |  |  |  |
| A68. Fracture diagnosis should be based on visual evaluation and include assessment of grade/severity. Morphometry alone is not recommended because it is unreliable for diagnosis. | Fair | B | W |  |  | <sup>32</sup> |  |  |  |  |  |
| A69. The Genant visual semi-quantitative method is the current clinical technique of choice for diagnosing vertebral fracture with VFA.* | Good | B | W |  |  | <sup>32</sup> | <sup>33</sup> |  |  |  |  |
| A70. Severity of deformity may be confirmed by morphometric measurement if desired. | Fair | B | W |  |  | <sup>32</sup> |  |  |  |  |  |

\* Original 2005 PDC wording, "The severity of vertebral fractures may be determined using the semiquantitative (SQ) assessment criteria developed by Genant."

### 18. Indications for Following VFA with Another Imaging Modality

| Position | Quality | Strength | Applicability | 2001 | 2003 | 2005 | 2007 | 2010 | 2013 | 2015 | 2019 |
| --- | --- | --- | --- | --- | --- | --- | --- | --- | --- | --- | --- |
| A71. The decision to perform additional imaging must be based on each patient's overall clinical picture, including the VFA result. | Poor | C | W |  |  | <sup>32</sup> |  |  |  |  |  |
| A72. Indications for follow-up imaging studies include:<br>• Lesions in vertebrae that cannot be attributed to benign causes<br>• Vertebral deformities in a patient with a known history of a relevant malignancy<br>• Equivocal fractures<br>• Unidentifiable vertebrae between T7-L4 | Fair | C | W |  |  | <sup>32</sup> | <sup>33</sup> |  |  |  |  |
| Sclerotic or lytic changes, or findings suggestive of conditions other than osteoporosis* |  |  |  |  |  |  |  |  |  |  |  |
| Note: VFA is designed to detect vertebral fractures and not other abnormalities. | NA | NA | NA |  |  | 32 |  |  |  |  |  |

\*Original wording from 2005 PDC, “Consider additional imaging when there are: Equivocal fractures, Unidentifiable vertebrae between T7-L4, Sclerotic or lytic changes, or findings suggestive of conditions other than osteoporosis. Note: VFA is designed to detect vertebral fractures and not other abnormalities.”

### 19. Serial Lateral Imaging

| Position | Quality | Strength | Applicability | 2001 | 2003 | 2005 | 2007 | 2010 | 2013 | 2015 | 2019 |
| --- | --- | --- | --- | --- | --- | --- | --- | --- | --- | --- | --- |
| A73.Repeat VFA or radiographic lateral spine imaging in patients with continued high risk (e.g., historical height loss > 4 cm (>1.5 inches), self-reported but undocumented vertebral fracture, or glucocorticoid therapy equivalent to ≥ 5 mg of prednisone or equivalent per day for greater than or equal to three months). | Poor | C | W |  |  |  |  |  |  |  | 72 |

### 20. DXA to Detect Abnormalities in the Spectrum of AFF

Atypical femur fractures (AFF) are a type of fracture that occurs at the femur and is characterized by transverse or short oblique fractures with minimal or no trauma. Atypical femur fractures have been associated with the use of bisphosphonates, a class of drugs used to treat osteoporosis. ^81^

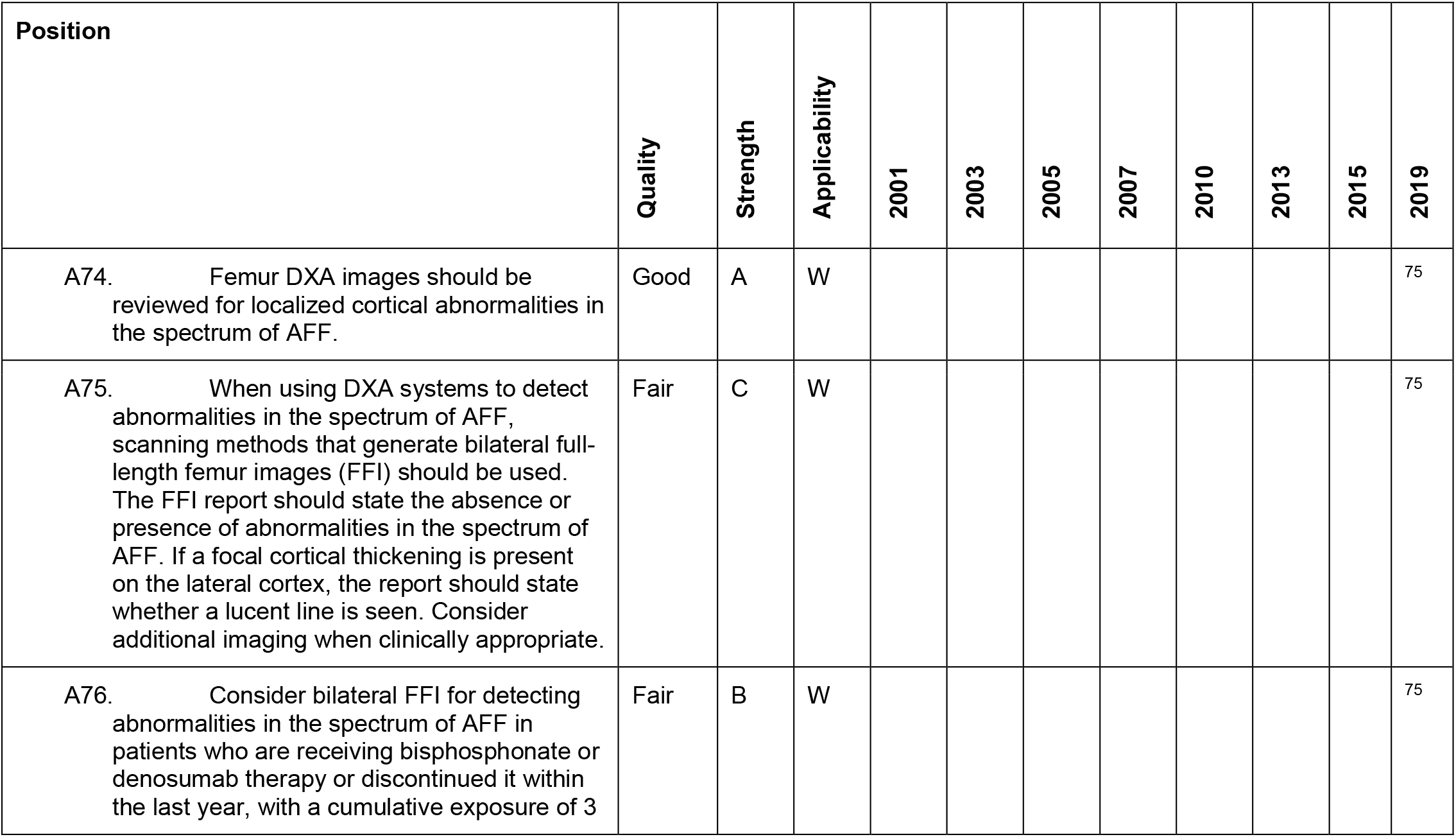
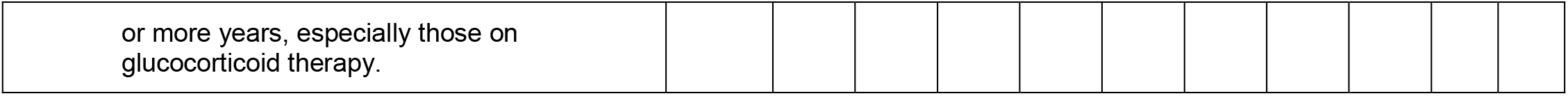

| Position | Quality | Strength | Applicability | 2001 | 2003 | 2005 | 2007 | 2010 | 2013 | 2015 | 2019 |
| --- | --- | --- | --- | --- | --- | --- | --- | --- | --- | --- | --- |
| A74. Femur DXA images should be reviewed for localized cortical abnormalities in the spectrum of AFF. | Good | A | W |  |  |  |  |  |  |  | 75 |
| A75. When using DXA systems to detect abnormalities in the spectrum of AFF, scanning methods that generate bilateral full-length femur images (FFI) should be used. The FFI report should state the absence or presence of abnormalities in the spectrum of AFF. If a focal cortical thickening is present on the lateral cortex, the report should state whether a lucent line is seen. Consider additional imaging when clinically appropriate. | Fair | C | W |  |  |  |  |  |  |  | 75 |
| A76. Consider bilateral FFI for detecting abnormalities in the spectrum of AFF in patients who are receiving bisphosphonate or denosumab therapy or discontinued it within the last year, with a cumulative exposure of 3 | Fair | B | W |  |  |  |  |  |  |  | 75 |
| or more years, especially those on glucocorticoid therapy. |  |  |  |  |  |  |  |  |  |  |  |

### 21. Baseline DXA Report: Minimum Requirements

These positions have not been updated in 20 years and most likely need revision to reflect the current needs of clinicians and patients.

| Position | Quality | Strength | Applicability | 2001 | 2003 | 2005 | 2007 | 2010 | 2013 | 2015 | 2019 |
| --- | --- | --- | --- | --- | --- | --- | --- | --- | --- | --- | --- |
| A77. Demographics (name, medical record identifying number, date of birth, sex). | NA | NA | NA |  | 25 |  |  |  |  |  |  |
| A78. Requesting provider. | NA | NA | NA |  | 25 |  |  |  |  |  |  |
| A79. Indications for the test. | NA | NA | NA |  | 25 |  |  |  |  |  |  |
| A80. Manufacturer and model of instrument used. | NA | NA | NA |  | 25 |  |  |  |  |  |  |
| A81. Technical quality and limitations of the study, stating why a specific site or ROI is invalid or not included. | NA | NA | NA |  | 25 |  |  |  |  |  |  |
| A82. BMD in g/cm<sup>2</sup> for each site. | NA | NA | NA |  | 25 |  |  |  |  |  |  |
| A83. The skeletal sites, ROI, and, if appropriate, the side, that were scanned. | NA | NA | NA |  | 25 |  |  |  |  |  |  |
| A84. The T-score and/or Z-score where appropriate. | NA | NA | NA |  | 25 |  |  |  |  |  |  |
| A85. WHO criteria for diagnosis in postmenopausal females and in men age 50 and over. | NA | NA | NA |  | 25 |  |  |  |  |  |  |
| A86. Risk factors including information regarding previous non-traumatic fractures. | NA | NA | NA |  | 25 |  |  |  |  |  |  |
| A87. A statement about fracture risk. Any use of relative fracture risk must specify the population of comparison (e.g., young-adult or age-matched). The ISCD favors the use of absolute fracture risk prediction when such methodologies are established. | NA | NA | NA |  | 25 |  |  |  |  |  |  |
| A88. A general statement that a medical evaluation for secondary causes of low BMD may be appropriate. | NA | NA | NA |  | 25 |  |  |  |  |  |  |
| A89. Recommendations for the necessity and timing of the next BMD study. | NA | NA | NA |  | 25 |  |  |  |  |  |  |

### 22. Follow-Up DXA Report

These positions have not been updated in 20 years and most likely need revision to reflect the current needs of clinicians and patients.

| Position | Quality | Strength | Applicability | 2001 | 2003 | 2005 | 2007 | 2010 | 2013 | 2015 | 2019 |
| --- | --- | --- | --- | --- | --- | --- | --- | --- | --- | --- | --- |
| A90. Statement regarding which previous or baseline study and ROI is being used for comparison. | NA | NA | NA |  | 25 |  |  |  |  |  |  |
| A91. Statement about the LSC at your facility and the statistical significance of the comparison. | NA | NA | NA |  | 25 |  |  |  |  |  |  |
| A92. Report significant change, if any, between the current and previous study or studies in g/cm2 and percentage. | NA | NA | NA |  | 25 |  |  |  |  |  |  |
| A93. Comments on any outside study including manufacturer and model on which previous studies were performed and the appropriateness of the comparison. | NA | NA | NA |  | 25 |  |  |  |  |  |  |
| A94. Recommendations for the necessity and timing of the next BMD study. | NA | NA | NA |  | 25 |  |  |  |  |  |  |

### 23. DXA Report: Optional Items

This topic has not been addressed since 2013. There have been many updates in DXA systems since then. These positions are most likely in need of update to reflect the current technological capabilities.

| Position | Quality | Strength | Applicability | 2001 | 2003 | 2005 | 2007 | 2010 | 2013 | 2015 | 2019 |
| --- | --- | --- | --- | --- | --- | --- | --- | --- | --- | --- | --- |
| A95. Recommendation for further non-BMD testing, such as X-ray, magnetic resonance imaging, computed tomography, etc. | NA | NA | NA |  | 25 |  |  |  |  |  |  |
| A96. Recommendations for pharmacological and nonpharmacological interventions. | NA | NA | NA |  | 25 |  |  |  |  |  |  |
| A97. Addition of the percentage compared to a reference population. | NA | NA | NA |  | 25 |  |  |  |  |  |  |
| A98. Specific recommendations for evaluation of secondary osteoporosis. | NA | NA | NA |  | 25 |  |  |  |  |  |  |

### 24. DXA Report: Items that Should not be Included

| Position | Quality | Strength | Applicability | 2001 | 2003 | 2005 | 2007 | 2010 | 2013 | 2015 | 2019 |
| --- | --- | --- | --- | --- | --- | --- | --- | --- | --- | --- | --- |
| A99. A statement that there is bone loss without knowledge of previous bone density. | NA | NA | NA |  | 25 |  |  |  |  |  |  |
| A100. Mention of “mild,” “moderate,” or “marked” osteopenia or osteoporosis. | NA | NA | NA |  | 25 |  |  |  |  |  |  |
| A101. Separate diagnoses for different ROI (eg, osteopenia at the hip and osteoporosis at the spine). | NA | NA | NA |  | 25 |  |  |  |  |  |  |
| A102. Expressions such as “She has the bones of an 80-year-old,” if the patient is not 80 yr old. | NA | NA | NA |  | 25 |  |  |  |  |  |  |
| A103. Results from skeletal sites that are not technically valid. | NA | NA | NA |  | 25 |  |  |  |  |  |  |
| A104. The change in BMD if it is not a significant change based on the precision error and LSC.* | NA | NA | NA | 19 | 25 |  |  |  |  |  |  |

\* Original wording from 2001, “The BMD difference must exceed LSC as determined by an in vivo precision study to be reported as significant.”

### 25. Components of a VFA report

Vertebral Fracture Assessment (VFA) is a low radiation method for imaging the lateral spine for fracture assessment. The 2005 and 2007 PDCs addressed pressing issues regarding indications of use, methodology, and recommendations for additional imaging. One additional position on the use of serial VFA was also addressed in 2019, A95.

| Position | Quality | Strength | Applicability | 2001 | 2003 | 2005 | 2007 | 2010 | 2013 | 2015 | 2019 |
| --- | --- | --- | --- | --- | --- | --- | --- | --- | --- | --- | --- |
| A105. Patient identification, referring physician, indication (s) for study, technical quality, and interpretation. | Poor | C | W |  |  | 32 |  |  |  |  |  |
| A106. A follow-up VFA report should also include comparability of studies and clinical significance of changes, if any. | Poor | C | W |  |  | 32 |  |  |  |  |  |
| A107. VFA reports should comment on the following:<br>• Unevaluable vertebrae<br>• Deformed vertebrae, and whether or not the deformities are consistent with vertebral fracture<br>• Unexplained vertebral and extra-vertebral pathology | Good | C | W |  |  |  | 33 |  |  |  |  |
| A108. Optional components include fracture risk and recommendations for additional studies. | Poor | C | W |  |  | 32 |  |  |  |  |  |

### 26. Trabecular Bone Score (TBS)

TBS is a proprietary measure of trabecular complexity in the spine. The 2015 and 2019 PDCs addressed the use of TBS but none of the existing positions were updated.

| Position | Quality | Strength | Applicability | 2001 | 2003 | 2005 | 2007 | 2010 | 2013 | 2015 | 2019 |
| --- | --- | --- | --- | --- | --- | --- | --- | --- | --- | --- | --- |
| A109. TBS is associated with vertebral, hip and major osteoporotic fracture risk in postmenopausal women. | Good | B | W |  |  |  |  |  |  | 66 |  |
| A110. TBS is associated with hip fracture risk in men over the age of 50 yr. | Fair | B | W |  |  |  |  |  |  | 66 |  |
| A111. TBS is associated with major osteoporotic fracture risk in men over the age of 50 yr. | Fair | B | W |  |  |  |  |  |  | 66 |  |
| A112. TBS should not be used alone to determine treatment recommendations in clinical practice. | Good | A | W |  |  |  |  |  |  | 66 |  |
| A113. TBS can be used in association with FRAX and BMD to adjust FRAX-probability of fracture in postmenopausal women and older men. | Good | B | W |  |  |  |  |  |  | 66 |  |
| In patients receiving anti-fracture therapy: |  |  |  |  |  |  |  |  |  | 66 |  |
| A114. The role of TBS in monitoring anti-resorptive therapy is unclear. | Good | A | W |  |  |  |  |  |  |  | 74 |
| A115. TBS is potentially useful for monitoring anabolic therapy. | Fair | B | W |  |  |  |  |  |  |  | 74 |
| A116. TBS is associated with major osteoporotic fracture risk in postmenopausal women with type II diabetes. | Fair | B | L |  |  |  |  |  |  | 66 |  |

### 27. Hip Geometry

These positions are for hip geometry measures that may be useful for fracture risk assessment including hip axis length (HAL), cross-sectional area (CSA), Outer shaft diameter (OD), section modulus (SM), buckling ratio (BR), cross-sectional moment of inertia (CSMI), Neck Shaft Angle (NSA). Positions on their use were addressed at the 2015 PDC and not since.

| Position | Quality | Strength | Applicability | 2001 | 2003 | 2005 | 2007 | 2010 | 2013 | 2015 | 2019 |
| --- | --- | --- | --- | --- | --- | --- | --- | --- | --- | --- | --- |
| A117. Hip axis length (HAL) derived from DXA is associated with hip fracture risk in postmenopausal women. | Fair | B | L |  |  |  |  |  |  | 65 |  |
| A118. The following hip geometry parameters derived from DXA (CSA, OD, SM, BR, CSMI, NSA) should not be used to assess hip fracture risk. | Fair | B | W |  |  |  |  |  |  | 65 |  |
| A119. Hip geometry parameters derived from DXA (CSA, OD, SM, BR, CSMI, HAL, NSA) should not be used to initiate treatment. | Fair | B | W |  |  |  |  |  |  | 65 |  |
| A120. Hip geometry parameters derived from DXA (CSA, OD, SM, BR, CSMI, HAL, NSA) should not be used for monitoring. | Good | A | W |  |  |  |  |  |  | 65 |  |

### 28. General Recommendations for Non-Central DXA Devices: QCT, pQCT, QUS and pDXA

The following general recommendations for QCT, pQCT, QUS, and pDXA are analogous to those defined for central DXA technologies. Examples of technical differences among devices, fracture prediction ability for current manufacturers, and equivalence study requirements are provided in the full text documents printed in the Journal of Clinical Densitometry.

| Position | Quality | Strength | Applicability | 2001 | 2003 | 2005 | 2007 | 2010 | 2013 | 2015 | 2019 |
| --- | --- | --- | --- | --- | --- | --- | --- | --- | --- | --- | --- |
| A121. Bone density measurements from different devices cannot be directly compared. | Good | A | W |  |  |  | 1, 35, 36 |  |  |  |  |
| A122. Different devices should be independently validated for fracture risk prediction by prospective trials, or by demonstration of equivalence to a clinically validated device. | Good | B | W |  |  |  | 1, 35, 36 |  |  |  |  |
| A123. T-scores from measurements other than DXA at the femur neck, total femur, lumbar spine, or one-third (33%) radius cannot be used according to the WHO diagnostic classification because those T-scores are not equivalent to T-scores derived by DXA. | Good | A | W |  |  |  | 1, 35, 36 |  |  |  |  |
| A124. Device-specific education and training should be provided to the operators and interpreters prior to clinical use. | Good | A | W |  |  |  | 1, 35, 36 |  |  |  |  |
| A125. Quality control procedures should be performed regularly. | Good | A | W |  |  |  | 1, 35, 36 |  |  |  |  |

### 29. Baseline Non Central Devices (QCT, pQCT, QUS, pDXA) Report: Minimum Requirements

Three papers are listed for each of these reporting requirements since they are in common for each of the listed technologies and they have not been revised since their inception.

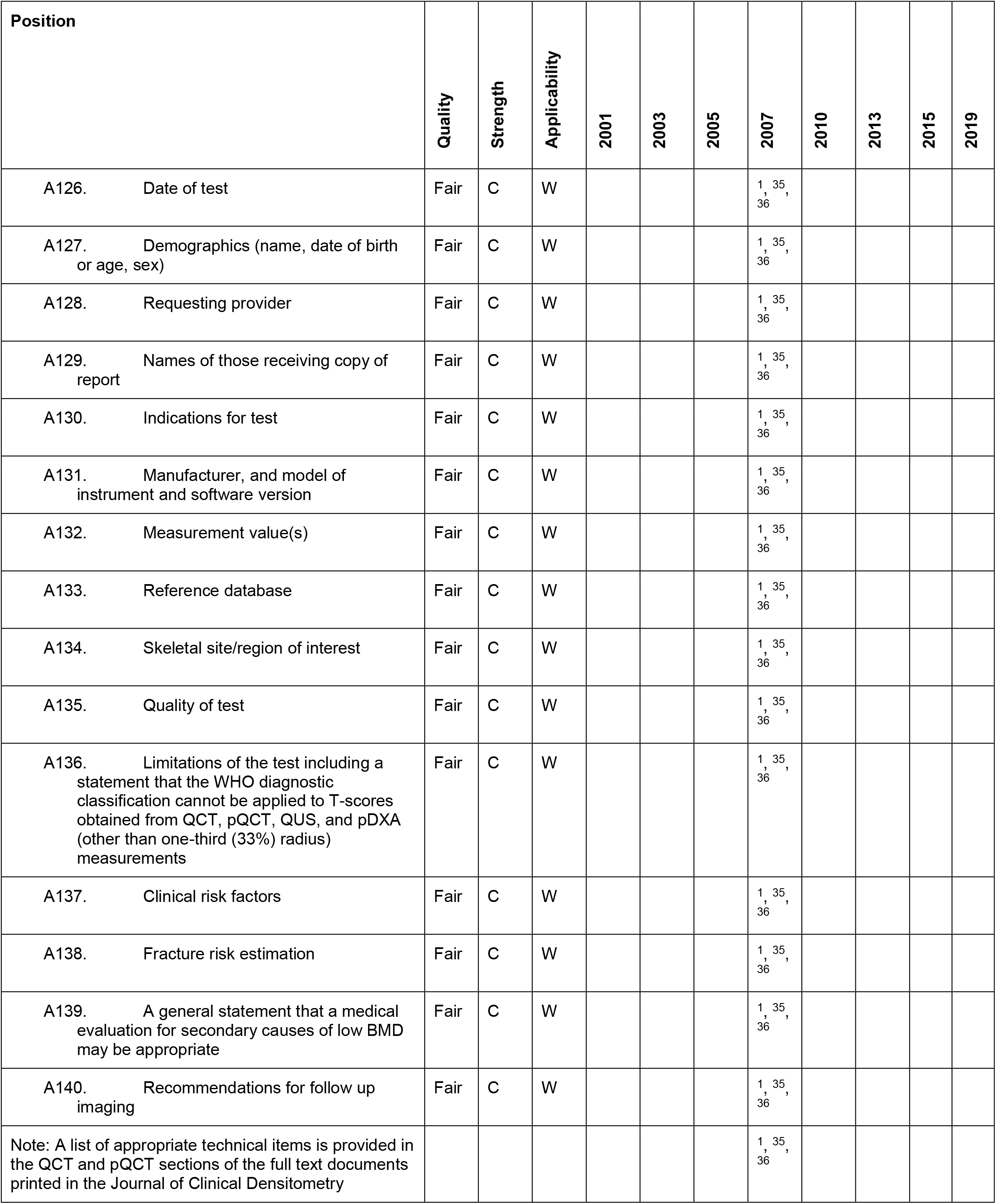

| Position | Quality | Strength | Applicability | 2001 | 2003 | 2005 | 2007 | 2010 | 2013 | 2015 | 2019 |
| --- | --- | --- | --- | --- | --- | --- | --- | --- | --- | --- | --- |
| A126. Date of test | Fair | C | W |  |  |  | 1, 35, 36 |  |  |  |  |
| A127. Demographics (name, date of birth or age, sex) | Fair | C | W |  |  |  | 1, 35, 36 |  |  |  |  |
| A128. Requesting provider | Fair | C | W |  |  |  | 1, 35, 36 |  |  |  |  |
| A129. Names of those receiving copy of report | Fair | C | W |  |  |  | 1, 35, 36 |  |  |  |  |
| A130. Indications for test | Fair | C | W |  |  |  | 1, 35, 36 |  |  |  |  |
| A131. Manufacturer, and model of instrument and software version | Fair | C | W |  |  |  | 1, 35, 36 |  |  |  |  |
| A132. Measurement value(s) | Fair | C | W |  |  |  | 1, 35, 36 |  |  |  |  |
| A133. Reference database | Fair | C | W |  |  |  | 1, 35, 36 |  |  |  |  |
| A134. Skeletal site/region of interest | Fair | C | W |  |  |  | 1, 35, 36 |  |  |  |  |
| A135. Quality of test | Fair | C | W |  |  |  | 1, 35, 36 |  |  |  |  |
| A136. Limitations of the test including a statement that the WHO diagnostic classification cannot be applied to T-scores obtained from QCT, pQCT, QUS, and pDXA (other than one-third (33%) radius) measurements | Fair | C | W |  |  |  | 1, 35, 36 |  |  |  |  |
| A137. Clinical risk factors | Fair | C | W |  |  |  | 1, 35, 36 |  |  |  |  |
| A138. Fracture risk estimation | Fair | C | W |  |  |  | 1, 35, 36 |  |  |  |  |
| A139. A general statement that a medical evaluation for secondary causes of low BMD may be appropriate | Fair | C | W |  |  |  | 1, 35, 36 |  |  |  |  |
| A140. Recommendations for follow up imaging | Fair | C | W |  |  |  | 1, 35, 36 |  |  |  |  |
| Note: A list of appropriate technical items is provided in the QCT and pQCT sections of the full text documents printed in the Journal of Clinical Densitometry |  |  |  |  |  |  | 1, 35, 36 |  |  |  |  |

### 30. Non Central DXA Devices (QCT, pQCT, QUS, pDXA) Report: Optional Items

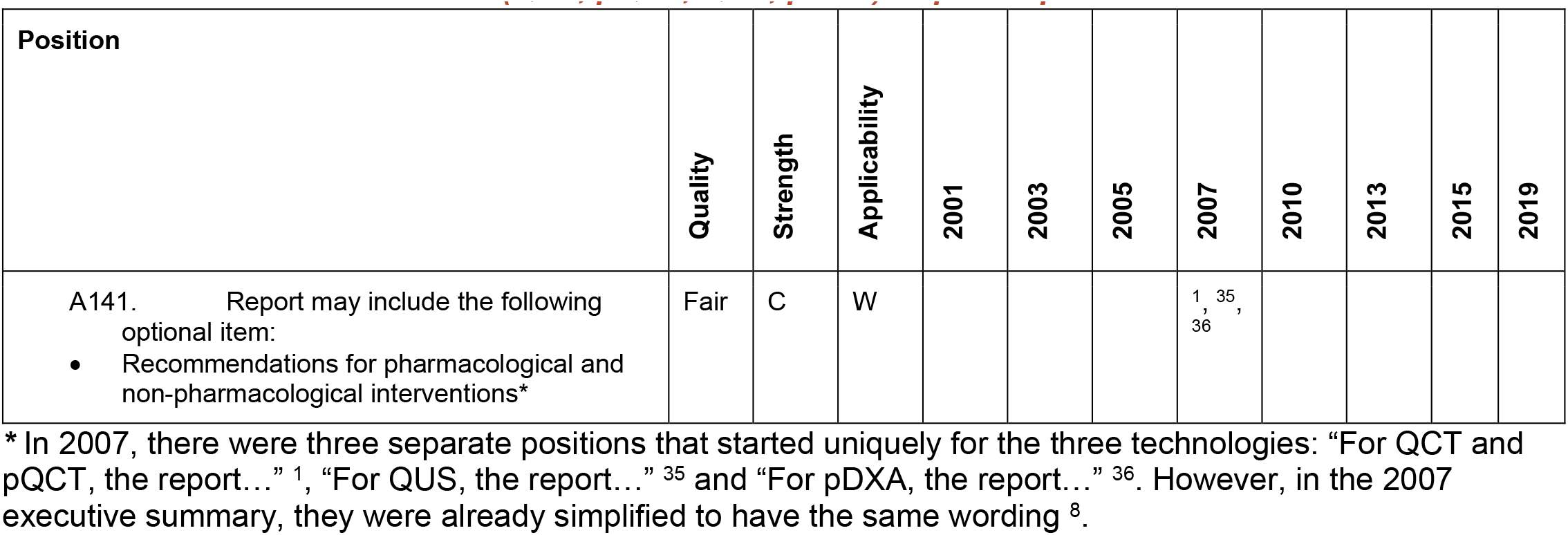

### 31. QCT and pQCT

Central quantitative computed tomography (QCT) of the spine and hip, and peripheral QCT (pQCT) have been addressed at the 2007 and 2015 PDCs. Further mention of CT is made in for orthopedic use in another section, see A241. Only one position, A173, has been updated. It addressed the situation when both QCT and DXA are available. The position was updated to state that DXA is preferred in this situation to limit radiation exposure.

| Position | Quality | Strength | Applicability | 2001 | 2003 | 2005 | 2007 | 2010 | 2013 | 2015 | 2019 |
| --- | --- | --- | --- | --- | --- | --- | --- | --- | --- | --- | --- |
| Acquisition – |  |  |  |  |  |  |  |  |  |  |  |
| A142. With single-slice QCT, L1-L3 should be scanned; with 3D QCT, L1-L2 should be scanned. | Fair | B | W |  |  |  | 1 |  |  |  |  |
| A143. QCT acquisition of the proximal femur should extend from the femoral head to the proximal shaft. | Good | A | W |  |  |  |  |  |  | 68 |  |
| A144. For density-based QCT measurements the in-scan calibration phantom can be replaced by asynchronous calibration if scanner stability is maintained. | Fair | B | W |  |  |  |  |  |  | 70 |  |
| A145. Opportunistic CT to screen for patients with low BMD or low bone strength of the spine or proximal femur is possible only if validated machine-specific cutoff values and scanner stability have been established. | Fair | C | W |  |  |  |  |  |  | 70 |  |
| Diagnosis – |  |  |  |  |  |  |  |  |  |  |  |
| A146. Femoral neck and total hip T-scores calculated from 2D projections of QCT data are equivalent to the corresponding DXA T-scores for diagnosis of osteoporosis in accordance with the WHO criteria. | Fair | B | W |  |  |  |  |  |  | 68 |  |
| Fracture Prediction – |  |  |  |  |  |  |  |  |  |  |  |
| A147. Spinal trabecular BMD as measured by QCT has at least the same ability to predict vertebral fractures as AP spinal BMD measured by central DXA in postmenopausal women. There is lack of sufficient evidence to support this position for men. | Fair | B | W |  |  |  | 1 |  |  |  |  |
| A148. There is lack of sufficient evidence to recommend spine QCT for hip fracture prediction in either women or men. | Good | A | W |  |  |  | 1 |  |  |  |  |
| A149. Total femur trabecular BMD measured by QCT predicts hip fractures as well as hip BMD measured by DXA in postmenopausal women and older men. | Fair | B | W |  |  |  |  |  |  | 68 |  |
| A150. pQCT of the forearm at the ultra-distal radius predicts hip, but not spine, fragility fractures in postmenopausal women. There is lack of sufficient evidence to support this position for men. | Fair | B | W |  |  |  | 1 |  |  |  |  |
| Therapeutic Decisions – |  |  |  |  |  |  |  |  |  |  |  |
| A151. Central DXA measurements at the spine and femur are the preferred method for making therapeutic decisions and should be used if possible. Where QCT and DXA are both available and provide comparable information, DXA is preferred to limit radiation exposure.* | Fair | C | W |  |  |  | 1 |  |  | 68 |  |
| A152. However, if central DXA cannot be done, pharmacologic treatment can be initiated if the fracture probability, as assessed by QCT of the spine or pQCT of the radius using device specific thresholds, and in conjunction with clinical risk factors, is sufficiently high. | Fair | B | W |  |  |  | 1 |  |  |  |  |
| Monitoring – |  |  |  |  |  |  |  |  |  |  |  |
| A153. Trabecular BMD of the lumbar spine measured by QCT can be used to monitor age-, disease- and treatment-related BMD changes. | Fair | B | W |  |  |  | 1 |  |  |  |  |
| A154. Integral and trabecular BMD of the proximal femur measured by QCT can be used to monitor age- and treatment-related BMD changes. | Fair | B | W |  |  |  |  |  |  | 68 |  |
| A155. Trabecular and total BMD of the ultra-distal radius measured by pQCT can be used to monitor age-related BMD changes. | Fair | B | W |  |  |  | 1 |  |  |  |  |
| Finite Element Analysis (FEA) – |  |  |  |  |  |  |  |  |  |  |  |
| A156. Vertebral strength as estimated by QCT-based FEA predicts vertebral fracture in postmenopausal women. | Fair | B | W |  |  |  |  |  |  | 69 |  |
| A157. Vertebral strength as estimated by QCT-based FEA is comparable to spine DXA for prediction of vertebral fractures in older men. | Fair | B | W |  |  |  |  |  |  | 69 |  |
| A158. Femoral strength as estimated by QCT-based FEA is comparable to hip DXA for prediction of hip fractures in postmenopausal women and older men. | Fair | B | W |  |  |  |  |  |  | 69 |  |
| A159. FEA cannot be used to diagnose osteoporosis using the current WHO T-score definition. | Good | A | W |  |  |  |  |  |  | 69 |  |
| A160. Vertebral or femoral strength as estimated by QCT-based FEA can be used to initiate pharmacologic treatment using validated thresholds and in conjunction with clinical risk factors. | Fair | B | W |  |  |  |  |  |  | 69 |  |
| A161. Vertebral or femoral strength as estimated by QCT-based FEA can be used to monitor age- and treatment-related changes. | Fair | B | W |  |  |  |  |  |  | 69 |  |
| Reporting – |  |  |  |  |  |  |  |  |  |  |  |
| A162. For QCT using whole body CT scanners the following additional technical items should be reported:<br>• Tomographic acquisition and reconstruction parameters<br>• kV, mAs<br>• Collimation during acquisition<br>• Table increment per rotation<br>• Table height<br>• Reconstructed slice thickness, reconstruction increment<br>• Reconstruction kernel | Fair | C | W |  |  |  | 1 |  |  |  |  |
| A163. For pQCT using dedicated pQCT scanners, the following additional technical items should be reported:<br>• Tomographic acquisition and reconstruction parameters<br>• Reconstructed slice thickness<br>• Single / multi-slice acquisition mode<br>• Length of scan range in multi-slice acquisition mode | Fair | C | W |  |  |  | 1 |  |  |  |  |

\* The 2007 version of this position did not include, “Where QCT and DXA are both available and provide comparable information, DXA is preferred to limit radiation exposure.”

### 32. QUS

Quantitative Ultrasound (QUS) systems were developed in the 1990s to evaluate fracture risk using the speed of sound (SOS) and/or broadband ultrasound attenuation (BUA). ^82 83^ Many regions on interest have been explored including the heel, proximal tibia, radius, phalanges, and recently the spine and hip. The use of QUS has only been addressed once at the 2007 PDC.

| Position | Quality | Strength | Applicability | 2001 | 2003 | 2005 | 2007 | 2010 | 2013 | 2015 | 2019 |
| --- | --- | --- | --- | --- | --- | --- | --- | --- | --- | --- | --- |
| Acquisition – |  |  |  |  |  |  |  |  |  |  |  |
| A164. The only validated skeletal site for the clinical use of QUS in osteoporosis management is the heel. | Good | A | W |  |  |  | 35 |  |  |  |  |
| Fracture Prediction – |  |  |  |  |  |  |  |  |  |  |  |
| A165. Validated heel QUS devices predict fragility fracture in postmenopausal women (hip, vertebral and global fracture risk) and men over the age of 65 (hip and all non-vertebral fractures) independently of central DXA BMD. | Good | A | W |  |  |  | 35 |  |  |  |  |
| A166. Discordant results between heel QUS and central DXA are not infrequent and are not necessarily an indication of methodological error. | Good | A | W |  |  |  | 35 |  |  |  |  |
| A167. Heel QUS in conjunction with clinical risk factors can be used to identify a population at very low fracture probability in which no further diagnostic evaluation may be necessary. (Examples of device-specific thresholds and case findings strategy are provided in the full text documents printed in the Journal of Clinical Densitometry.) | Good | B | W |  |  |  | 35 |  |  |  |  |
| Therapeutic Decisions – |  |  |  |  |  |  |  |  |  |  |  |
| A168. Central DXA measurements at the spine and femur are preferred for making therapeutic decisions and should be used if possible. However, if central DXA cannot be done, pharmacologic treatment can be initiated if the fracture probability, as assessed by heel QUS, using device specific thresholds and in conjunction with clinical risk factors, is sufficiently high. (Examples of device-specific thresholds are provided in the full text documents printed in the Journal of Clinical Densitometry.) | Fair | C | W |  |  |  | 35 |  |  |  |  |
| Monitoring – |  |  |  |  |  |  |  |  |  |  |  |
| A169. QUS cannot be used to monitor the skeletal effects of treatments for osteoporosis. | Good | A | W |  |  |  | 35 |  |  |  |  |

### 33. pDXA

Peripheral DXA (pDXA) systems are not as widely used in the US as central DXA systems, but still popular for bone evaluations by non-specialty providers (i.e. drug-store assessments) and in other countries. pDXA has only been addressed at the 2007 PDC.

| Position | Quality | Strength | Applicability | 2001 | 2003 | 2005 | 2007 | 2010 | 2013 | 2015 | 2019 |
| --- | --- | --- | --- | --- | --- | --- | --- | --- | --- | --- | --- |
| Fracture Prediction – |  |  |  |  |  |  |  |  |  |  |  |
| A170. Measurement by validated pDXA devices can be used to assess vertebral and global fragility fracture risk in postmenopausal women; however, its vertebral fracture predictive ability is weaker than central DXA and heel QUS. There is lack of sufficient evidence to support this position for men. | Fair | B | W |  |  |  | 36 |  |  |  |  |
| A171. Radius pDXA in conjunction with clinical risk factors can be used to identify a population at very low fracture probability in which no further diagnostic evaluation may be necessary. (Examples of device-specific thresholds and case findings strategy are provided in the full text documents printed in the Journal of Clinical Densitometry.) | Fair | B | W |  |  |  | 36 |  |  |  |  |
| Diagnosis – |  |  |  |  |  |  |  |  |  |  |  |
| A172. The WHO diagnostic classification can only be applied to DXA at the femur neck, total femur, lumbar spine, and the one-third (33%) radius ROI measured by DXA or pDXA devices utilizing a validated young-adult reference database. | Good | A | W |  |  |  | 36 |  |  |  |  |
| Therapeutic Decisions – |  |  |  |  |  |  |  |  |  |  |  |
| A173. Central DXA measurements at the spine and femur are the preferred method for making therapeutic decisions and should be used if possible. However, if central DXA cannot be done, pharmacologic treatment can be initiated if the fracture probability, as assessed by radius pDXA (or DXA) using device specific thresholds and in conjunction with clinical risk factors, is sufficiently high. (Examples of device-specific thresholds are provided in the full text documents printed in the Journal of Clinical Densitometry.) | Fair | B | W |  |  |  | 36 |  |  |  |  |
| Monitoring – |  |  |  |  |  |  |  |  |  |  |  |
| A174. pDXA devices are not clinically useful in monitoring the skeletal effects of presently available medical treatments for osteoporosis. | Good | A | W |  |  |  | 36 |  |  |  |  |

### 34. Body Composition

Body composition by DXA has been available on DXA systems since 1987. However, there were no positions on indications for use, measurement, or reporting before the 2013 PDC. The topic has not been addressed since.

| Position | Quality | Strength | Applicability | 2001 | 2003 | 2005 | 2007 | 2010 | 2013 | 2015 | 2019 |
| --- | --- | --- | --- | --- | --- | --- | --- | --- | --- | --- | --- |
| Indications – |  |  |  |  |  |  |  |  |  |  |  |
| A175. DXA total body composition with regional analysis can be used in the following conditions: |  |  |  |  |  |  |  |  |  |  |  |
| A176. In patients living with HIV to assess fat distribution in those using anti-retroviral agents associated with a risk of lipoatrophy (currently stavudine [d4T] and zidovudine [ZDV, AZT]). | Good | B | W |  |  |  |  |  | 57 |  |  |
| A177. In obese patients undergoing bariatric surgery (or medical, diet, or weight loss regimens with anticipated large weight loss) to assess fat and lean mass changes when weight loss exceeds approximately 10%. The impact on clinical outcomes is uncertain. | Poor | C | W |  |  |  |  |  | 57 |  |  |
| A178. In patients with muscle weakness or poor physical functioning to assess fat and lean mass. The impact on clinical outcomes is uncertain. | Fair | C | W |  |  |  |  |  | 57 |  |  |
| A179. Pregnancy is a contraindication to DXA body composition. Limitations in the use of clinical DXA for total body composition or BMD are weight over the table limit, recent administration of contrast material and/or artifact. Radiopharmaceutical agents may interfere with accuracy of results using systems from some DXA manufacturers. | Fair | B | W |  |  |  |  |  | 57 |  |  |
| Acquisition – |  |  |  |  |  |  |  |  |  |  |  |
| A180. No phantom has been identified to remove systematic differences in body composition when comparing in-vivo results across manufacturers. | Good | B | W |  |  |  |  |  | 59 |  |  |
| A181. An in-vivo cross-calibration study is necessary when comparing in-vivo results across manufacturers. | Fair | B | W |  |  |  |  |  |  |  |  |
| A182. Cross-calibrating systems of the same make and model can be performed with an appropriate whole-body phantom. | Fair | B | W |  |  |  |  |  | 59 |  |  |
| A183. Changes in body composition measures can be evaluated between 2 different systems of the same make and | Fair | B | W |  |  |  |  |  | 59 |  |  |
| model if the systems have been cross-calibrated with an appropriate total body phantom. |  |  |  |  |  |  |  |  |  |  |  |
| A184. When changing hardware, but not the entire system, or when replacing a system with the same technology (make and model), cross-calibration should be performed by having 1 technologist do 10 whole-body phantom scans, with repositioning, before and after hardware change. If a greater than 2% difference in mean percent fat mass, fat mass or lean mass is observed, contact the manufacturer for service/correction. | Fair | B | W |  |  |  |  |  | 59 |  |  |
| A185. No total body phantoms are available at this time that can be used as absolute reference standards for soft-tissue composition or bone mineral mass. | Good | A | W |  |  |  |  |  | 59 |  |  |
| A186. The Quality Control (QC) program at a DXA body composition facility should include adherence to manufacturer guidelines for system maintenance. In addition, if not recommended in the manufacturer protocol, the following QC procedures are advised:<br>• Perform periodic (at least once per week) body composition phantom scans for any DXA system as an independent assessment of system calibration.<br>• Plot and review data from calibration and body composition phantom scans.<br>• Verify the body composition phantom mean percent fat mass and tissue mass after any service performed on the densitometer.<br>• Establish and enforce corrective action thresholds that trigger a call for service.<br>• Maintain service logs.<br>• Comply with radiation surveys and regulatory government inspections, radiation surveys and regulatory requirements. | Fair | B | W |  |  |  |  |  | 59 |  |  |
| A187. Consistent positioning and preparation (eg, fasting state, clothing, time of day, physical activity, empty bladder) of the patient is important for precise measures. | Fair | B | W |  |  |  |  |  | 59 |  |  |
| A188. Positioning of the arms, hands, legs, and feet whenever possible should be according to the NHANES method (palms down isolated from the body, feet neutral, ankles strapped, arms straight or slightly angled, face up with neutral chin). | Fair | B | W |  |  |  |  |  | 59 |  |  |
| A189. "Offset-scanning" should be used in patients who are too wide to fit within the scan boundaries, using a validated procedure for a specific scanner model. | Fair | B | W |  |  |  |  |  | 59 |  |  |
| A190. Every technologist should perform an in-vivo precision assessment for all body composition measures of interest using patients who are representative of the clinic's patient population. | Fair | B | W |  |  |  |  |  | 59 |  |  |
| A191. The minimum acceptable precision for an individual technologist is 3%, 2%, and | Fair | B | W |  |  |  |  |  | 59 |  |  |
| 2% for total fat mass, total lean mass, and percent fat mass, respectively. |  |  |  |  |  |  |  |  |  |  |  |
| A192. Consistently use manufacturer's recommendations for ROI placement. | Fair | B | W |  |  |  |  |  | 59 |  |  |
| A193. Consistently use manufacturer's recommendations for artifact removal. | Fair | B | W |  |  |  |  |  | 59 |  |  |
| Analysis and Reporting – |  |  |  |  |  |  |  |  |  |  |  |
| A194. For adults total body (with head) values of BMI, BMD, BMC, total mass, total lean mass, total fat mass, and percent fat mass should appear on all reports. | Fair | C | W |  |  |  |  |  | 58 |  |  |
| A195. Total body BMC as represented in the NHANES 1999 2004 reference data should be used when using DXA in 4-compartment models. | Fair | B | W |  |  |  |  |  | 58 |  |  |
| A196. DXA measures of adiposity and lean mass include visceral adipose tissue (VAT), appendicular lean mass index (ALMI: appendicular lean mass/ht<sup>2</sup>), android/gynoid percent fat mass ratio, trunk to leg fat mass ratio, lean mass index (LMI: total lean mass/ht<sup>2</sup>), fat mass index (FMI: fat mass/ht<sup>2</sup>) are optional. The clinical utility of these measures is currently uncertain. | Fair | C | W |  |  |  |  |  | 58 |  |  |
| A197. When comparing to the US population, the NHANES 1999 2004 body composition data are most appropriate for different races, both sexes, and for ages from 8 to 85 yr. (Note: Reference to a population does not imply health status.) | Fair | C | L |  |  |  |  |  | 58 |  |  |
| A198. Both Z-scores and percentiles are appropriate to report if derived using methods to adjust for non-normality. | Fair | C | W |  |  |  |  |  | 58 |  |  |
| A199. The use of DXA adiposity measures (percent fat mass or fat mass index) may be useful in risk-stratifying patients for cardio-metabolic outcomes. Specific thresholds to define obesity have not been established. | Fair | C | W |  |  |  |  |  | 58 |  |  |
| A200. "Low lean mass" could be defined using appendicular lean mass divided by height squared (ALM/height<sup>2</sup>) with Z-scores derived from a young adult, race, and sex-matched population. Thresholds for low lean mass from consensus guidelines for sarcopenia await confirmation. | Fair | C | W |  |  |  |  |  | 58 |  |  |

### 35. DXA in Patients with Spinal Cord Injury

Clinical care for patients with spinal cord injuries has been previously limited by the lack of consensus derived guidelines or standards regarding bone density testing and fracture risk prediction. No positions of the ISCD were directly applicable to those with spinal cord injury before the 2019 PDC.

| Position | Quality | Strength | Applicability | 2001 | 2003 | 2005 | 2007 | 2010 | 2013 | 2015 | 2019 |
| --- | --- | --- | --- | --- | --- | --- | --- | --- | --- | --- | --- |
| A201. All adults with SCI resulting in permanent motor or sensory dysfunction should have a DXA scan of the total hip, proximal tibia, and distal femur, as soon as medically stable. | Fair | B | W |  |  |  |  |  |  |  | 78 |
| A202. In adults with SCI, total hip, proximal tibia, and distal femur bone density should be used to diagnose osteoporosis, predict lower extremity fracture risk, and monitor response to therapy when normative data are available. | Fair | B | W |  |  |  |  |  |  |  | 78 |
| A203. Serial DXA assessment of treatment effectiveness among individuals with SCI should include evaluation at the total hip, distal femur, and proximal tibia, following a minimum of 12 months of therapy at 1- to 2-year intervals. Segmental analysis of total hip, distal femur and proximal tibia sub-regions from a whole-body scan should not be used for monitoring treatment. | Fair | B | W |  |  |  |  |  |  |  | 78 |
| A204. There is no established threshold BMD value below which weight-bearing activities are absolutely contraindicated. BMD and clinical risk factors should be used to assess fracture risk prior to engaging in weight-bearing activities. | Poor | C | W |  |  |  |  |  |  |  | 78 |

### 36. DXA in Transgender and Gender Non-conforming individuals

All aspects of bone density testing in transgender and gender nonconforming (TGNC) individuals were poorly defined before the 2019 PDC.

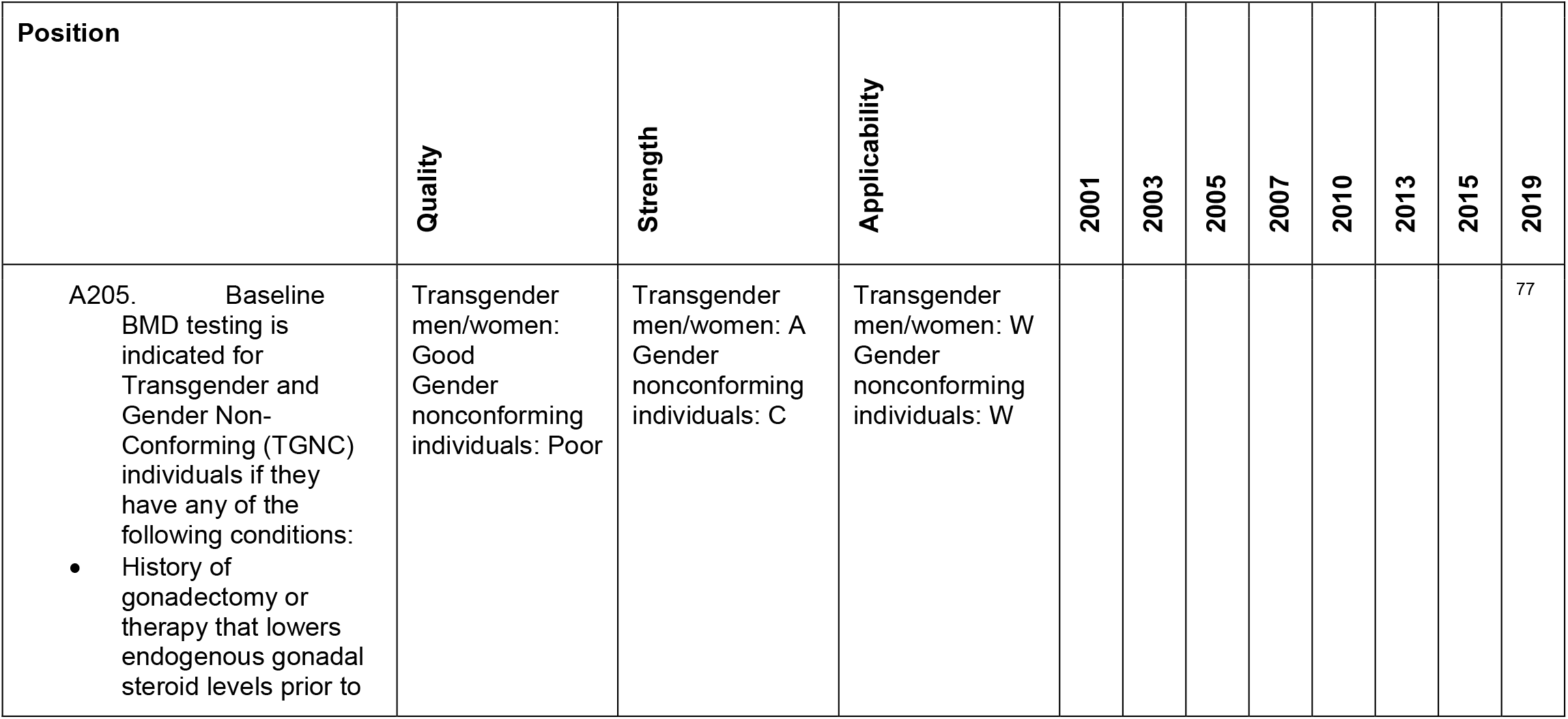
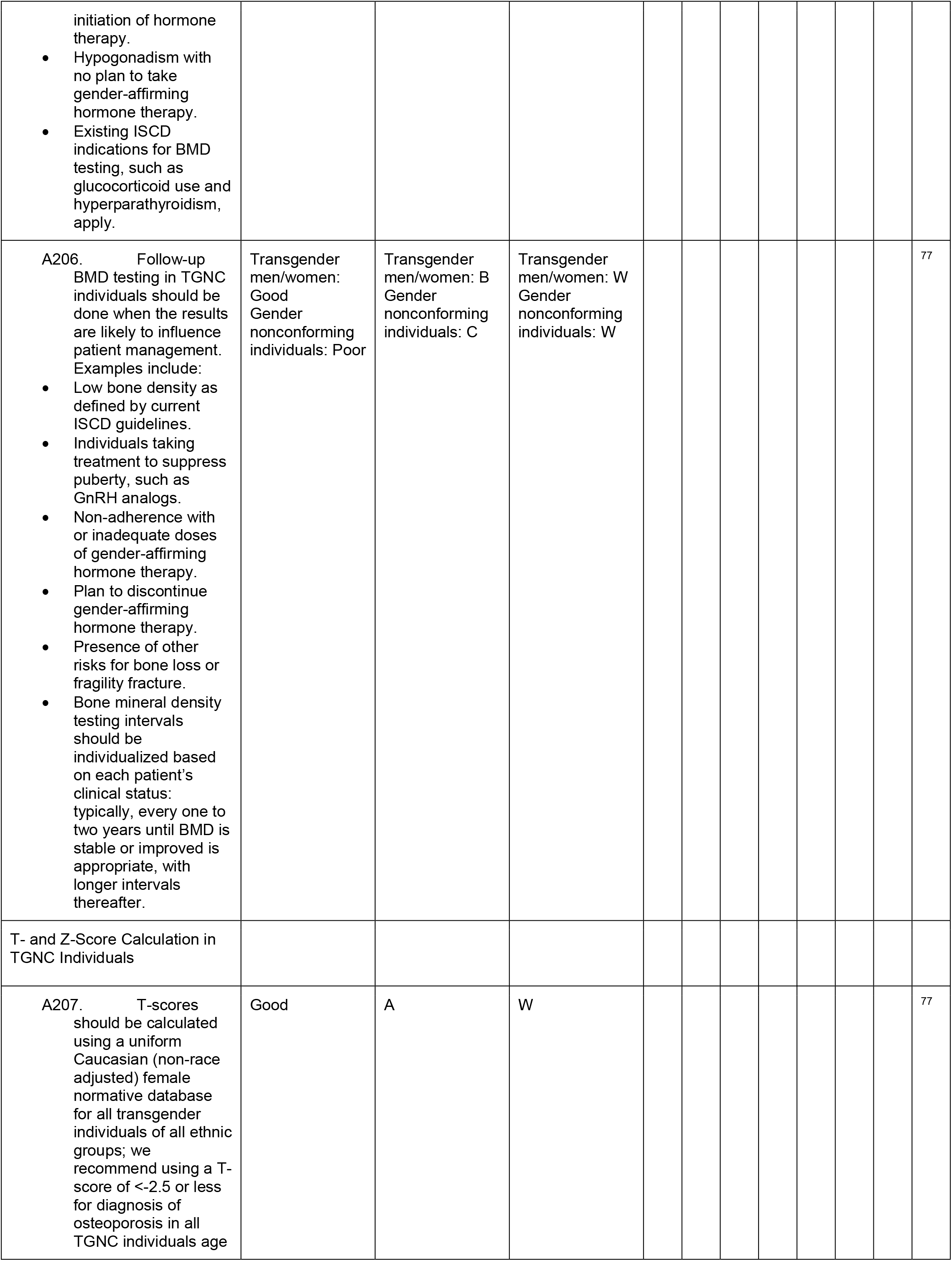
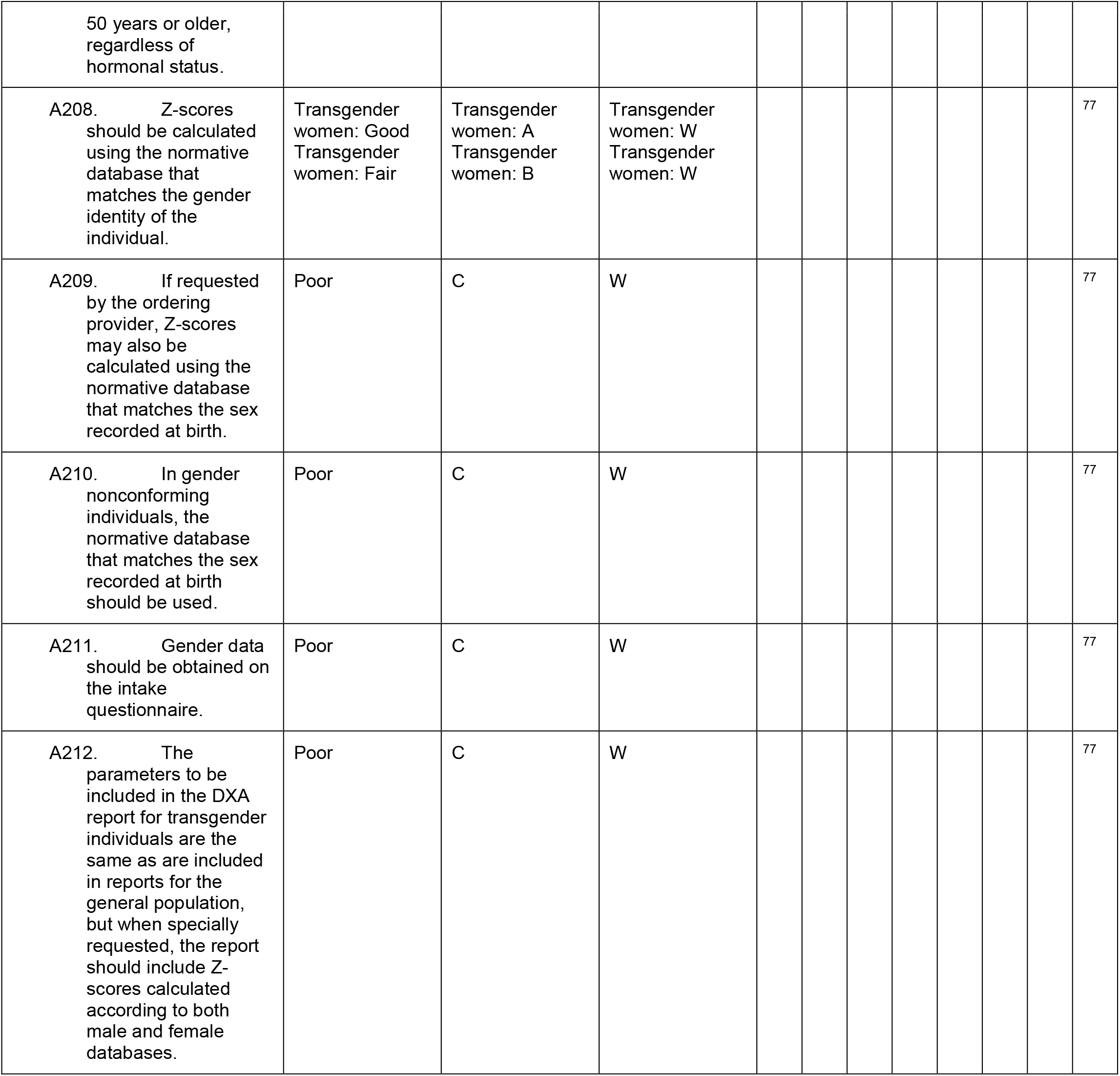

### 37. Peri-prosthetic and orthopedic uses of DXA

Although the ability to measure bone surrounding metal protheses has been available for over 20 years, positions on the utility and reporting of peri-prosthetic bone was not addressed until 2019. The questions considered included, which orthopedic surgery patients should be evaluated for poor bone health prior to surgery, which subsets of patients are at high risk for poor bone health and adverse outcomes, what is the reliability and validity of using bone densitometry techniques and measurement of specific geometries around the hip and knee before and after arthroplasty was determined, how can CT be used to estimate bone quality at common orthopedic surgery sites.

| Position | Quality | Strength | Applicability | 2001 | 2003 | 2005 | 2007 | 2010 | 2013 | 2015 | 2019 |
| --- | --- | --- | --- | --- | --- | --- | --- | --- | --- | --- | --- |
| A213. Bone health assessment should be considered in patients prior to elective orthopedic and spine surgery. BMD should be measured in those meeting ISCD or regional indications for DXA testing.<br>• Routine DXA scans should include PA lumbar spine and hip.<br>• Forearm DXA should be considered in patients having upper limb surgery.<br>• VFA should be considered in patients having spine surgery. | Poor | C | W |  |  |  |  |  |  |  | 76 |
| A214. Elective orthopedic and spine surgery patients with the following conditions are at greater risk for impaired bone health and should have DXA testing:<br>• Diabetes mellitus (long term duration of diabetes (>10yrs) and poor control)<br>◦ Trabecular bone score measurement should be obtained in patients with diabetes, if available.<br>• Inflammatory arthritis<br>• Exposed to chronic corticosteroids ( ≥ 5mg/day for three or more months of treatment)<br>• A low-trauma fracture after 50 years of age<br>• Chronic kidney disease stage 3, 4 and 5<br>• Limited mobility<br>• Smoking | Fair | B | W |  |  |  |  |  |  |  | 76 |
| A215. When poor bone quality is identified during surgery, bone health assessment including DXA testing is indicated. | Poor | C | W |  |  |  |  |  |  |  | 76 |
| A216. When assessing hip and knee arthroplasty, ROI should include periprosthetic metaphyseal and diaphyseal bone around and away from the implant:<br>• After total hip arthroplasty, Gruen zones are recommended at the femur and the DeLee / Charnley or Wilkinson method are recommended at the pelvis.<br>• Modifications of ROI based on patient conditions and implant geometry are acceptable. | Good | B | W |  |  |  |  |  |  |  | 76 |
| A217. Indications for pre-operative DXA testing for patients having hip surgery include:<br>• A Dorr classification of B or C.<br>• A Cortical Index of less than 0.4 measured at 10 cm below the mid lesser trochanter. | Fair | B | W |  |  |  |  |  |  |  | 76 |
| A218. The Cortical Index and/or cortical thickness adjacent to the femoral hip implant can be used to monitor bone ingrowth or resorption, identify periprosthetic loosening, predict subsidence, and assess the effectiveness of medical and surgical | Fair | B | W |  |  |  |  |  |  |  | 76 |
| methods to modulate BMD around the hip prostheses. |  |  |  |  |  |  |  |  |  |  |  |
| A219. Opportunistic CT-based attenuation using Hounsfield Units (HU) can be used to estimate the likelihood of osteoporosis (L1 HU < 100) and normal (L1 HU > 150) bone density to support decisions regarding bone health assessment. | Fair | B | W |  |  |  |  |  |  |  | 76 |

## FRAX POSITIONS BY CATEGORY

The FRAX calculator is a fracture risk assessment tool developed by the World Health Organization (WHO) ^53^. It is widely used to estimate the 10-year probability of a major osteoporotic fracture in an individual based on their age, sex, body mass index, other risk factors, bone mineral density and TBS score (if available). The FRAX calculator is used in clinical practice to identify patients who are at high risk for fragility fractures and who may benefit from preventive interventions. The FRAX statements are organized and numbered in the same fashion as found in the ISCD brochure. FRAX has only been addressed once by the ISCD, at the 2010 PDC. Thus, no edits are presented.

### 1. FRAX Introductory Statement

| Position | Quality | Strength | Applicability | 2001 | 2003 | 2005 | 2007 | 2010 | 2013 | 2015 | 2019 |
| --- | --- | --- | --- | --- | --- | --- | --- | --- | --- | --- | --- |
| F1. FRAX is a computer-based algorithm which uses easily obtained clinical risk factors to estimate an individual's 10-year fracture probability. It may be utilized by clinicians to assist in the identification of patients at high risk for fractures. | NA | NA | NA |  |  |  |  |  |  |  |  |

### 2. FRAX Clinical Statements

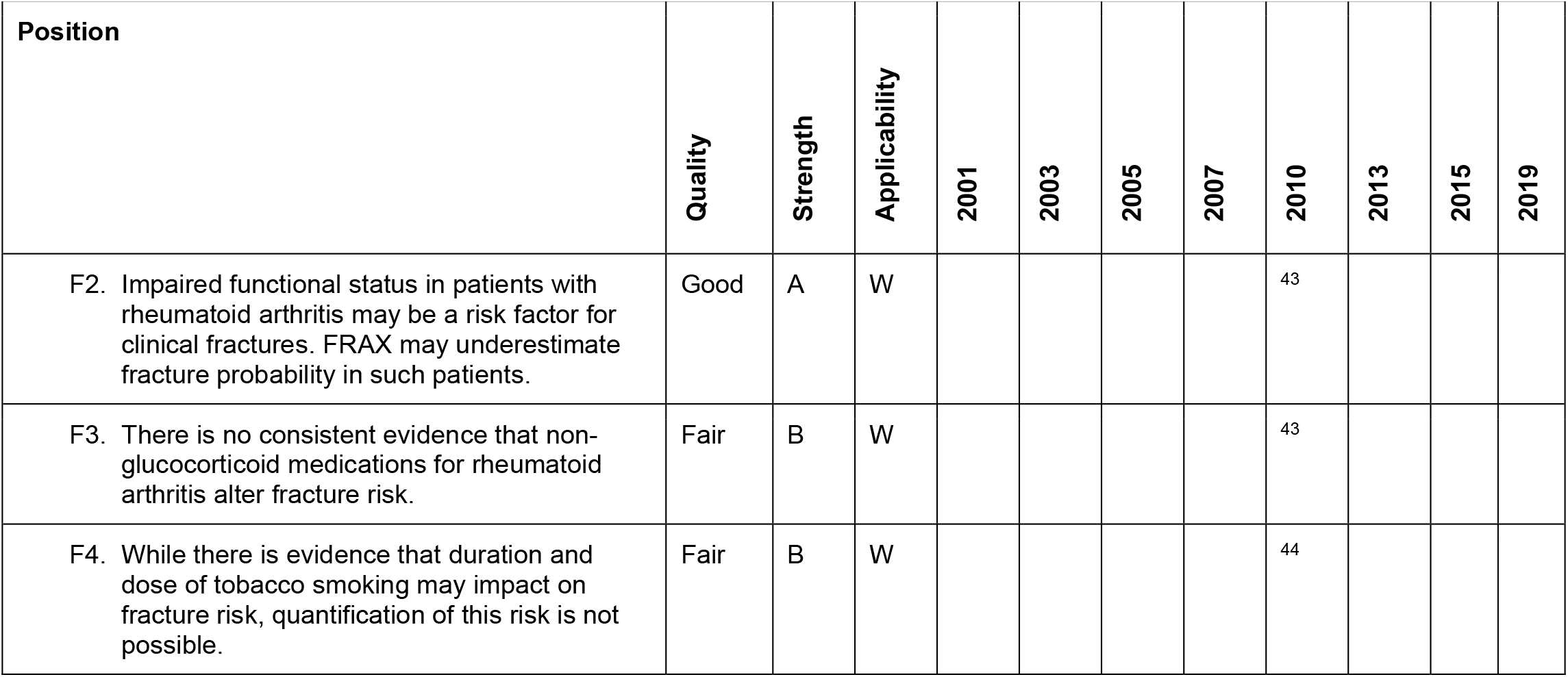
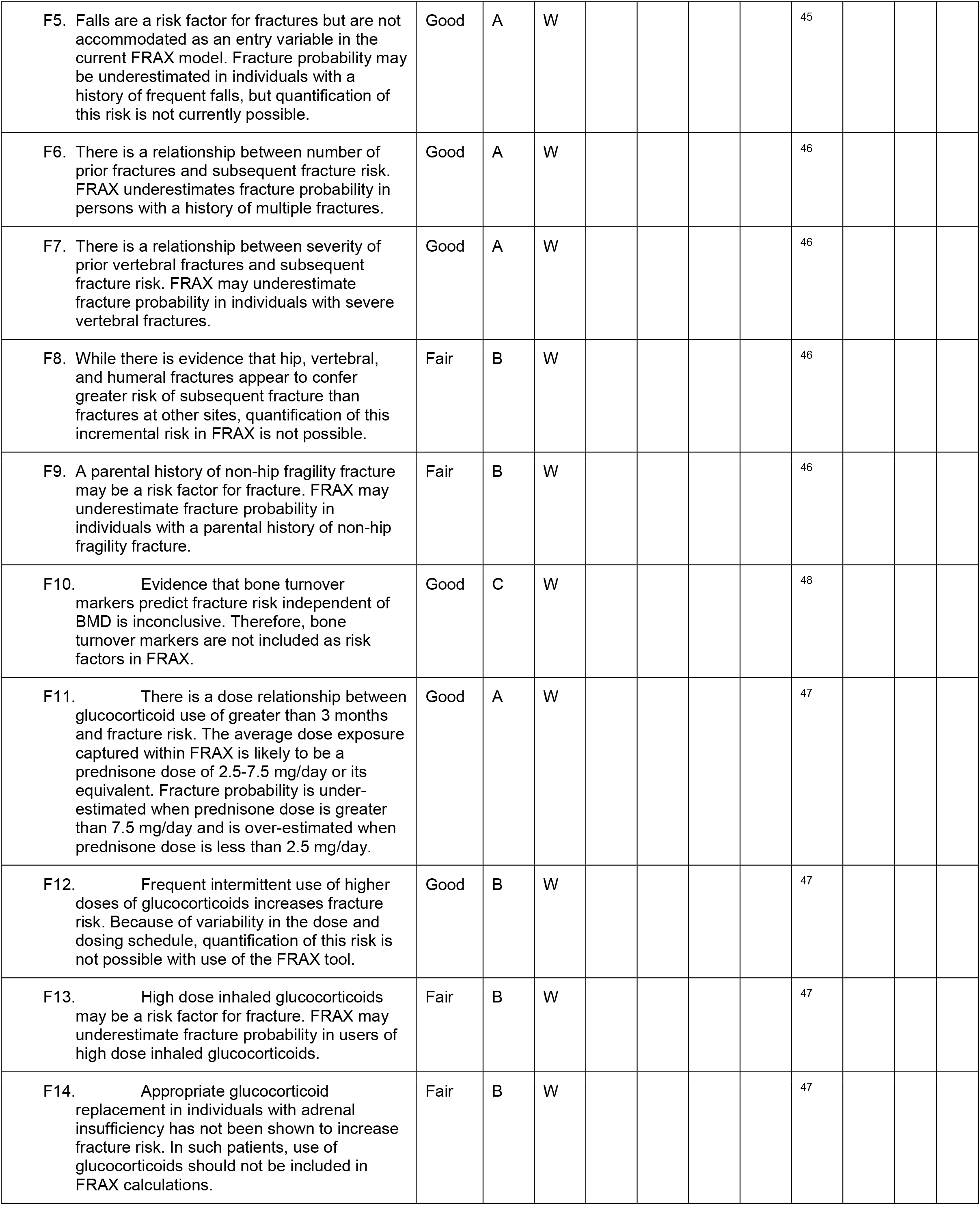

| Position | Quality | Strength | Applicability | 2001 | 2003 | 2005 | 2007 | 2010 | 2013 | 2015 | 2019 |
| --- | --- | --- | --- | --- | --- | --- | --- | --- | --- | --- | --- |
| F2. Impaired functional status in patients with rheumatoid arthritis may be a risk factor for clinical fractures. FRAX may underestimate fracture probability in such patients. | Good | A | W |  |  |  |  | 43 |  |  |  |
| F3. There is no consistent evidence that non-glucocorticoid medications for rheumatoid arthritis alter fracture risk. | Fair | B | W |  |  |  |  | 43 |  |  |  |
| F4. While there is evidence that duration and dose of tobacco smoking may impact on fracture risk, quantification of this risk is not possible. | Fair | B | W |  |  |  |  | 44 |  |  |  |
| F5. Falls are a risk factor for fractures but are not accommodated as an entry variable in the current FRAX model. Fracture probability may be underestimated in individuals with a history of frequent falls, but quantification of this risk is not currently possible. | Good | A | W |  |  |  |  | 45 |  |  |  |
| F6. There is a relationship between number of prior fractures and subsequent fracture risk. FRAX underestimates fracture probability in persons with a history of multiple fractures. | Good | A | W |  |  |  |  | 46 |  |  |  |
| F7. There is a relationship between severity of prior vertebral fractures and subsequent fracture risk. FRAX may underestimate fracture probability in individuals with severe vertebral fractures. | Good | A | W |  |  |  |  | 46 |  |  |  |
| F8. While there is evidence that hip, vertebral, and humeral fractures appear to confer greater risk of subsequent fracture than fractures at other sites, quantification of this incremental risk in FRAX is not possible. | Fair | B | W |  |  |  |  | 46 |  |  |  |
| F9. A parental history of non-hip fragility fracture may be a risk factor for fracture. FRAX may underestimate fracture probability in individuals with a parental history of non-hip fragility fracture. | Fair | B | W |  |  |  |  | 46 |  |  |  |
| F10. Evidence that bone turnover markers predict fracture risk independent of BMD is inconclusive. Therefore, bone turnover markers are not included as risk factors in FRAX. | Good | C | W |  |  |  |  | 48 |  |  |  |
| F11. There is a dose relationship between glucocorticoid use of greater than 3 months and fracture risk. The average dose exposure captured within FRAX is likely to be a prednisone dose of 2.5-7.5 mg/day or its equivalent. Fracture probability is underestimated when prednisone dose is greater than 7.5 mg/day and is over-estimated when prednisone dose is less than 2.5 mg/day. | Good | A | W |  |  |  |  | 47 |  |  |  |
| F12. Frequent intermittent use of higher doses of glucocorticoids increases fracture risk. Because of variability in the dose and dosing schedule, quantification of this risk is not possible with use of the FRAX tool. | Good | B | W |  |  |  |  | 47 |  |  |  |
| F13. High dose inhaled glucocorticoids may be a risk factor for fracture. FRAX may underestimate fracture probability in users of high dose inhaled glucocorticoids. | Fair | B | W |  |  |  |  | 47 |  |  |  |
| F14. Appropriate glucocorticoid replacement in individuals with adrenal insufficiency has not been shown to increase fracture risk. In such patients, use of glucocorticoids should not be included in FRAX calculations. | Fair | B | W |  |  |  |  | 47 |  |  |  |

### 3. FRAX BMD Statements

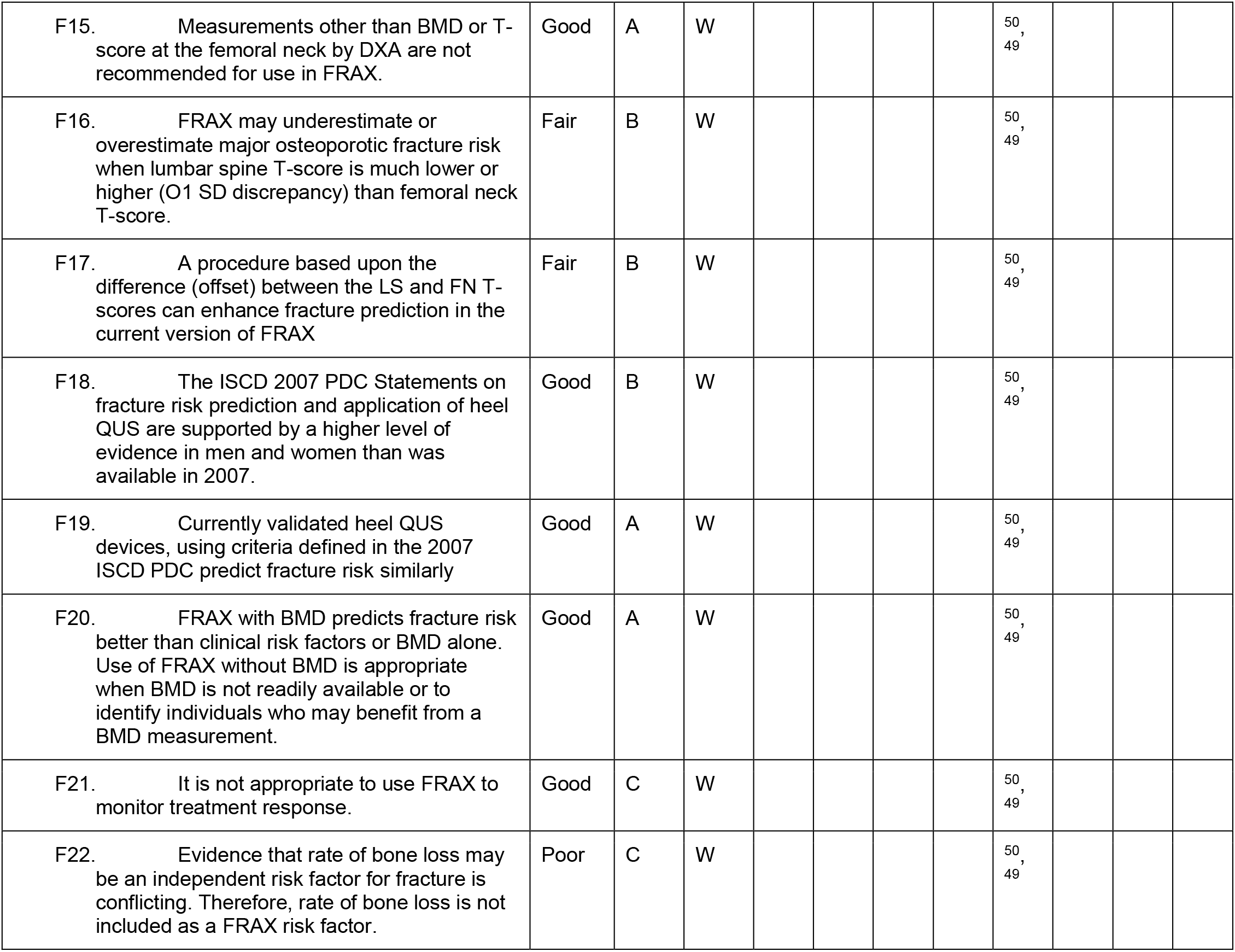

### 4. FRAX International Statements

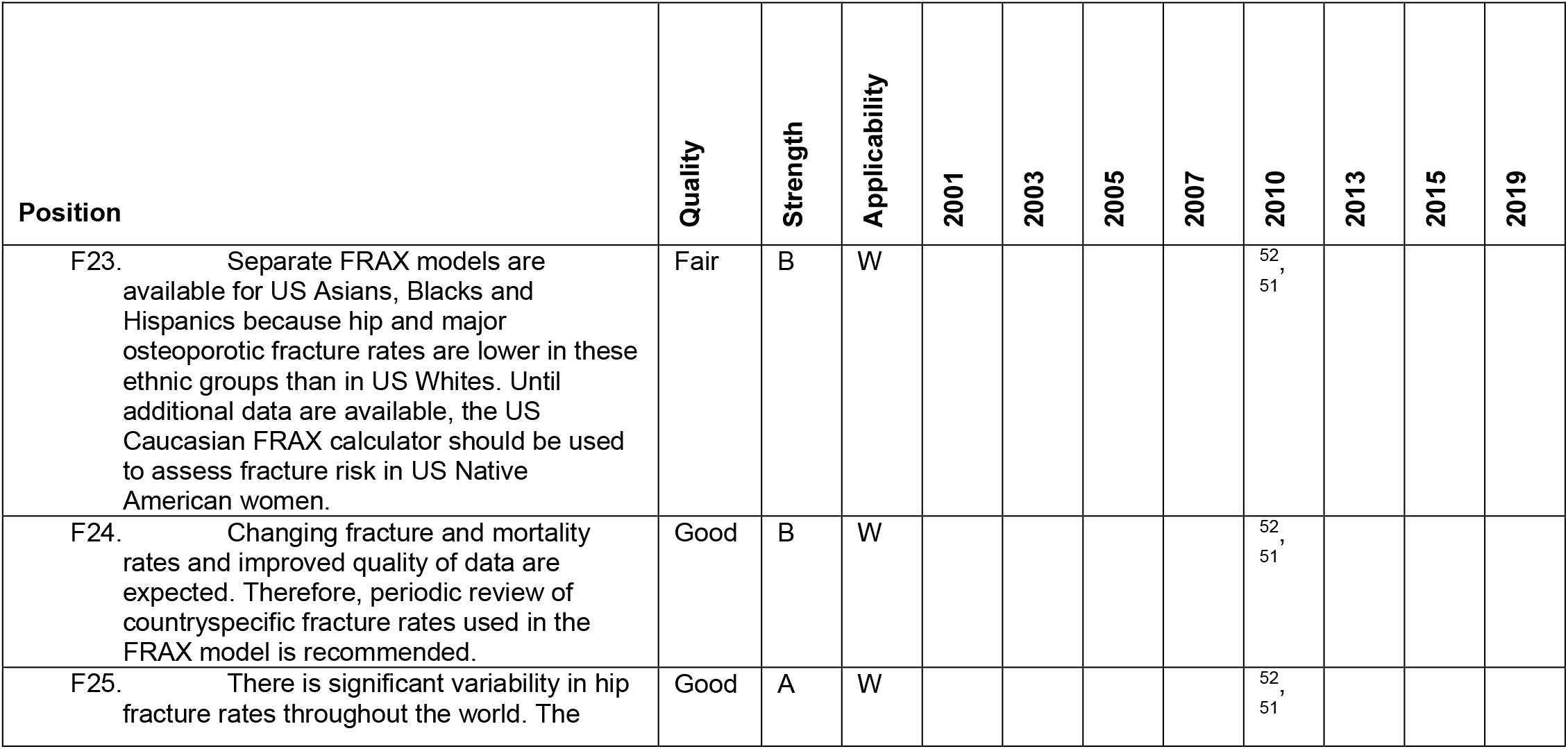
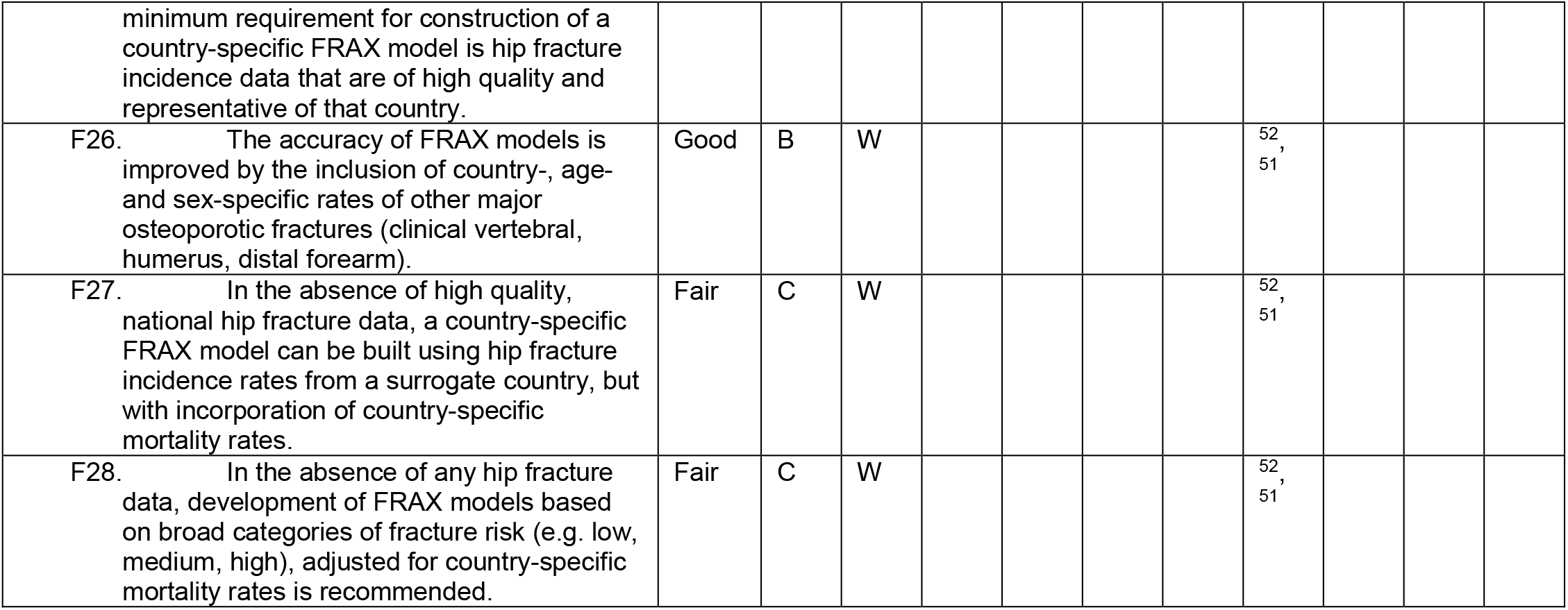

| Position | Quality | Strength | Applicability | 2001 | 2003 | 2005 | 2007 | 2010 | 2013 | 2015 | 2019 |
| --- | --- | --- | --- | --- | --- | --- | --- | --- | --- | --- | --- |
| F23. Separate FRAX models are available for US Asians, Blacks and Hispanics because hip and major osteoporotic fracture rates are lower in these ethnic groups than in US Whites. Until additional data are available, the US Caucasian FRAX calculator should be used to assess fracture risk in US Native American women. | Fair | B | W |  |  |  |  | 52,<br>51 |  |  |  |
| F24. Changing fracture and mortality rates and improved quality of data are expected. Therefore, periodic review of countryspecific fracture rates used in the FRAX model is recommended. | Good | B | W |  |  |  |  | 52,<br>51 |  |  |  |
| F25. There is significant variability in hip fracture rates throughout the world. The | Good | A | W |  |  |  |  | 52,<br>51 |  |  |  |
| minimum requirement for construction of a country-specific FRAX model is hip fracture incidence data that are of high quality and representative of that country. |  |  |  |  |  |  |  |  |  |  |  |
| F26. The accuracy of FRAX models is improved by the inclusion of country-, age- and sex-specific rates of other major osteoporotic fractures (clinical vertebral, humerus, distal forearm). | Good | B | W |  |  |  |  | 52,<br>51 |  |  |  |
| F27. In the absence of high quality, national hip fracture data, a country-specific FRAX model can be built using hip fracture incidence rates from a surrogate country, but with incorporation of country-specific mortality rates. | Fair | C | W |  |  |  |  | 52,<br>51 |  |  |  |
| F28. In the absence of any hip fracture data, development of FRAX models based on broad categories of fracture risk (e.g. low, medium, high), adjusted for country-specific mortality rates is recommended. | Fair | C | W |  |  |  |  | 52,<br>51 |  |  |  |

## PEDIATRIC POSITIONS BY CATEGORY

Special considerations must be made when evaluating bone health in children. The size of the bone is a challenge to DXA systems since they were designed for larger adult bones. These positions were created to be exclusive to children without referral to the adult positions. Thus, some positions may seem similar to their adults counterparts.

### 1. Fracture Prediction and Definition of Osteoporosis

Fracture prediction and the definition of osteoporosis was first addressed at the 2007 Pediatric PDC. These positions were reviewed, and modifications were made to define significant fracture history and draw attention to the degree of trauma in fracture risk prediction. Extensive discussion can be found in Bishop et al. ^63^. The fracture risk statements are largely qualitative due to the lack of quantitative risk models in children.

| Position | Quality | Strength | Applicability | 2001 | 2003 | 2005 | 2007 | 2010 | 2013 | 2015 | 2019 |
| --- | --- | --- | --- | --- | --- | --- | --- | --- | --- | --- | --- |
| P1. Evaluation of bone health should identify children and adolescents who may benefit from interventions to decrease their elevated risk of a clinically significant fracture.* | Fair/Good | B/C | W |  |  |  | 38 |  | 63 |  |  |
| P2. The finding of one or more vertebral compression (crush) fractures is indicative of osteoporosis, in the absence of local disease or high-energy trauma. In such children and adolescents, measuring BMD adds to the overall assessment of bone health. | Fair | B/C | W |  |  |  |  |  | 63 |  |  |
| P3. The diagnosis of osteoporosis in children and adolescents should not be made on the basis of densitometric criteria alone. | Poor | C | W |  |  |  | 38 |  | 63 |  |  |
| P4. In the absence of vertebral compression (crush) fractures, the diagnosis of osteoporosis is indicated by the presence of both a clinically significant fracture history and BMD Z-score $\leq -2.0$. A clinically significant fracture history is one or more of the following: 1) two or more long bone | Poor | C | W | | | | 38 | | 63 | | |
| fractures by age 10 years; 2) three or more long bone fractures at any age up to age 19 years. A BMC/BMD Z-score > -2.0 does not preclude the possibility of skeletal fragility and increased fracture risk.* |  |  |  |  |  |  |  |  |  |  |  |

\* The original position from 2007, "Fracture prediction should primarily identify children at risk of clinically significant fractures, such as fracture of long bones in the lower extremities, vertebral compression fractures, or two or more long-bone fractures of the upper extremities." was split into two positions, P1 and P4.

### 2. DXA Assessment in Children and Adolescents with Disease that May Affect the Skeleton

Like other pediatric position categories, there has been substantial changes in the positions over the years. Three positions from the 2007 Pediatric PDC were removed from the ISCD positions in 2013. The removed positions are the following.

> *In patients with thalassemia major, spine and TBLH BMC and areal BMD should be performed at fracture presentation or age 10 yr, whichever is earlier (Fair-C-W);*
>
> *Therapeutic interventions should not be instituted on the basis of a single DXA measurement (Fair-C-W);*
>
> *When technically feasible, all patients should have spine and TBLH BMC and areal BMD measured before initiation of bone active treatment and to monitor bone-active treatment in conjunction with other clinical data (Poor-C-W)*.

In 2007, they were all based on expert opinion only (grade C) and in 2013 that expert opinion had changed in favor of removing the positions. The full discussion is beyond the scope of this review but can be found in Bianchi et al. ^64^.

| Position | Quality | Strength | Applicability | 2001 | 2003 | 2005 | 2007 | 2010 | 2013 | 2015 | 2019 |
| --- | --- | --- | --- | --- | --- | --- | --- | --- | --- | --- | --- |
| P5. DXA measurement is part of a comprehensive skeletal health assessment in patients with increased risk of fracture. | Good | B | W |  |  |  | 39 |  |  |  |  |
| P6. In patients with primary bone disease, or at risk for secondary bone disease, a DXA should be performed when the patient may benefit from interventions to decrease their elevated risk of a clinically significant fracture, and the DXA results will influence that management.* | Fair | B | W |  |  |  | 39 |  | 64 |  |  |
| P7. DXA should not be performed if safe and appropriate positioning of the child cannot be assured. | Fair | C | W |  |  |  | 39 |  | 64 |  |  |

\* This position was substantially revised to be more conservative. The original position was "In patients with primary bone diseases or potential secondary bone diseases (e.g. due to chronic inflammatory diseases, endocrine disturbances, history of childhood cancer, or prior transplantation (non-renal)), spine and TBLH BMC and areal BMD should be measured at clinical presentation."

### 3. DXA Interpretation and Reporting in Children and Adolescents

Reporting of densitometry for pediatrics has been under extensive revision since the 2007 PDC. The most dramatic changes in interpretation was with the proximal femur. See P11. This site was not originally recommended in 2007 primarily due to expert opinion regarding measures across the ever-changing mineralization around the growth plate of the greater trochanter. However, as evidence amassed, the proximal femur was found to be as precise as other bone regions and pediatric measurements would be useful for following an adolescent into adulthood. Further, many of the positions in this section were downgraded in their quality of evidence and strength of recommendation.

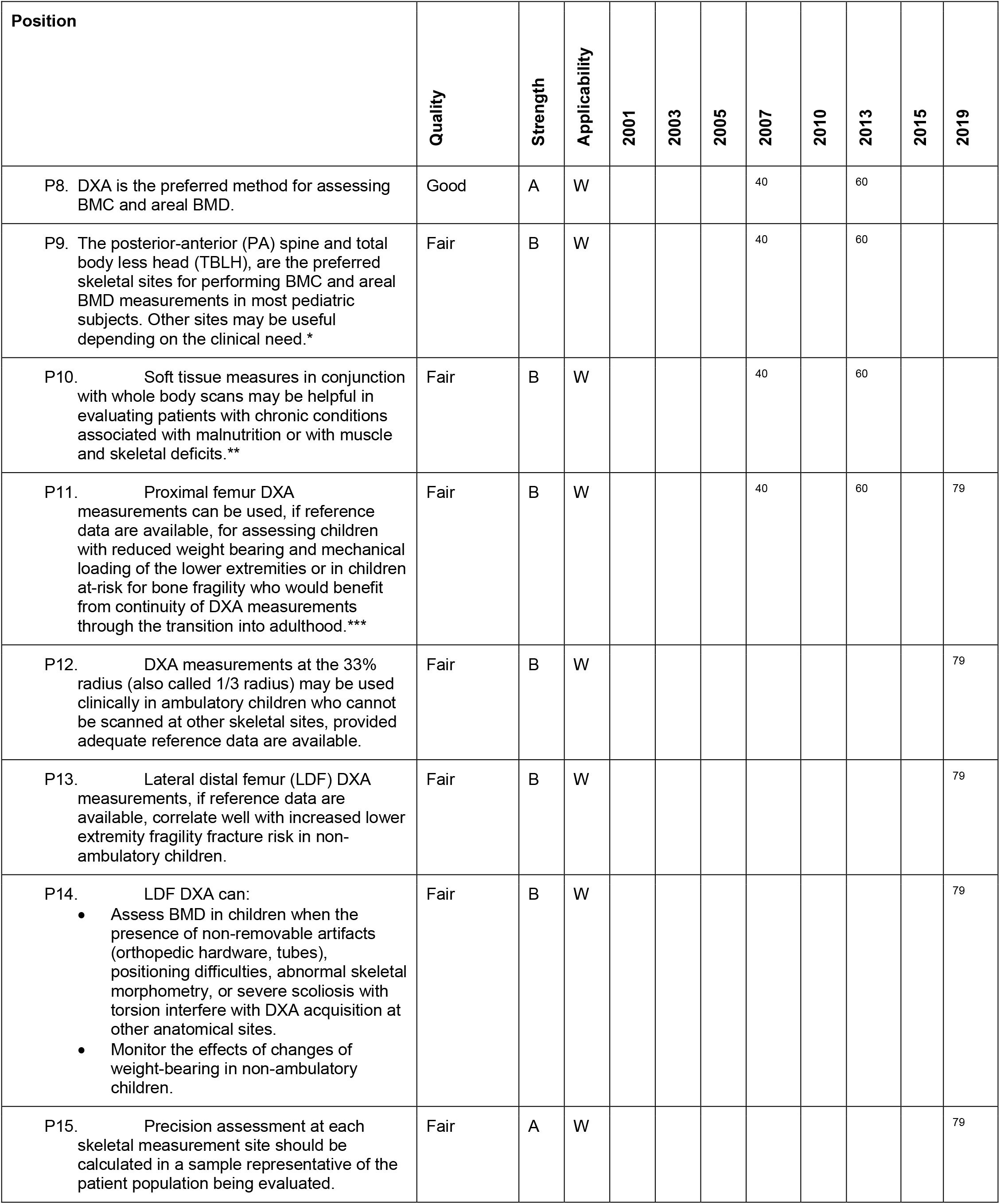
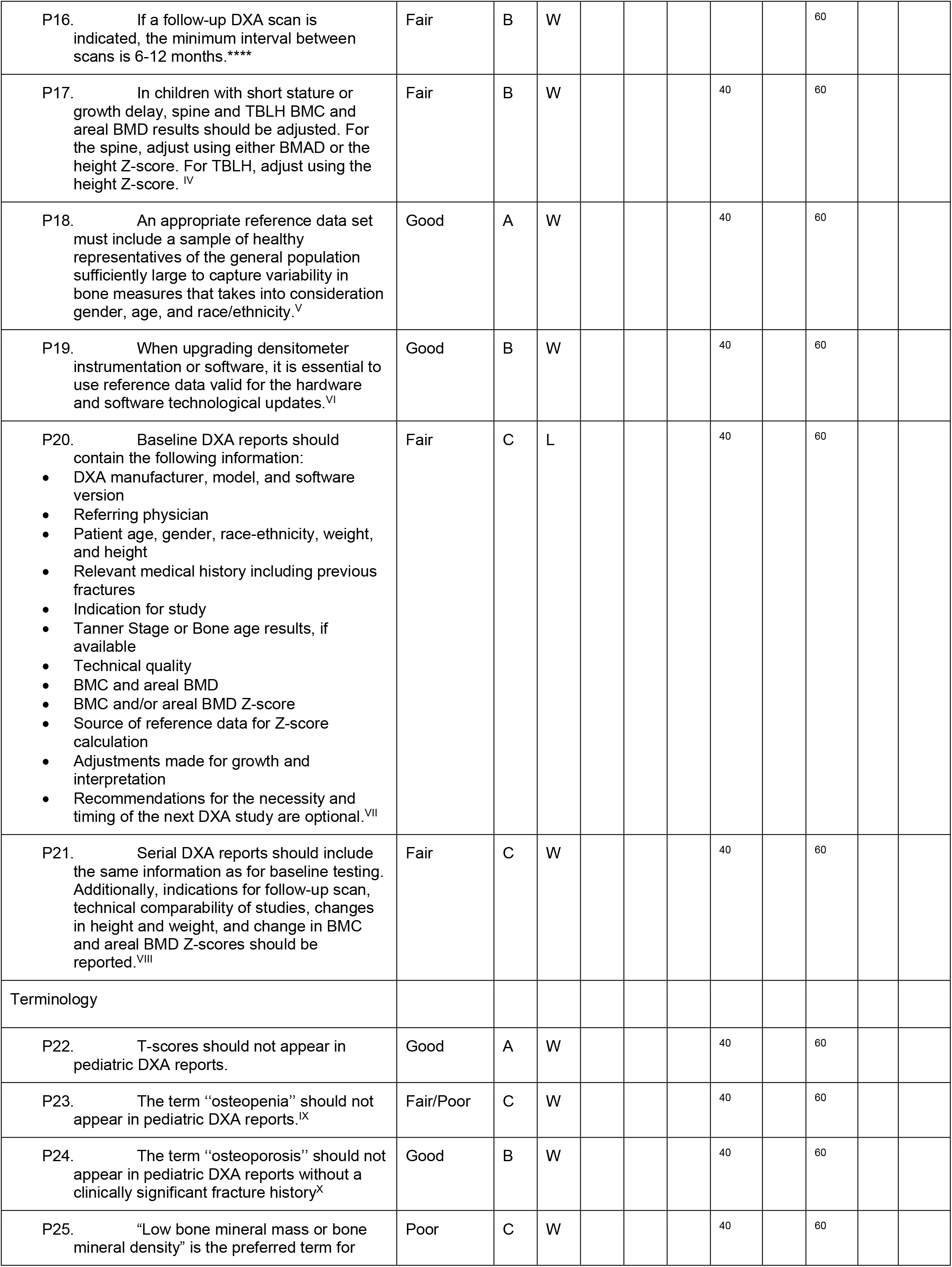
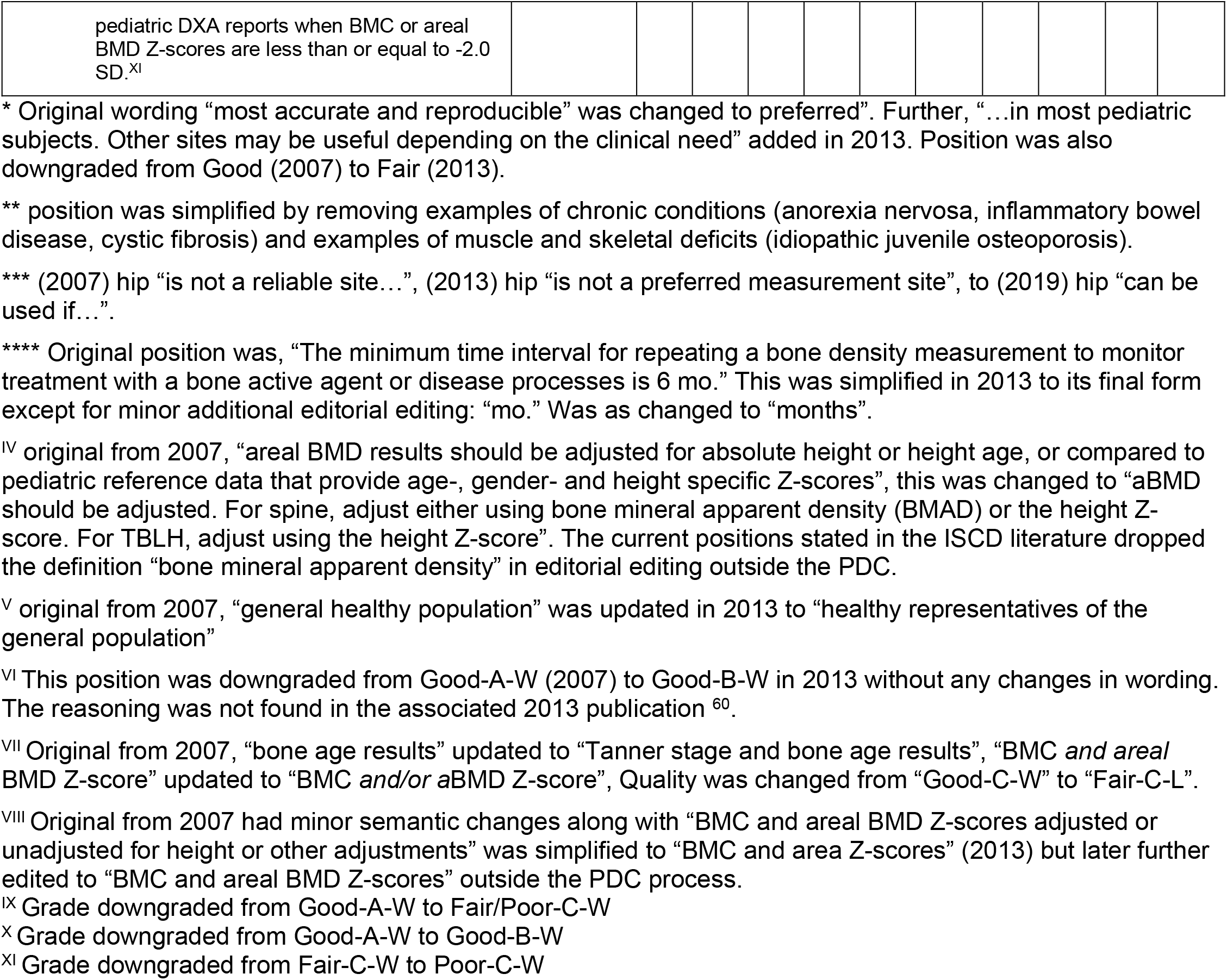

### 4. VFA in Pediatric Patients

The strengths and limitations of vertebral fracture assessment (VFA) by DXA were evaluated for use in children at the 2019 PDC. These positions are in response to the following questions: Should VFA be used as a substitute for spine radiography in the identification of symptomatic / asymptomatic osteoporotic VF in children? When does an abnormal VFA in a child require follow-up spine imaging? What is the VFA method that should be used to detect an osteoporotic VF in children? Are there technical and biological factors that limit the accuracy of DXA-based VFA in children (for example DXA model and software, age, sex, pubertal stage, obesity)? The full discussion of these questions and derivation of the positions can be found in Weber et al. ^79^.

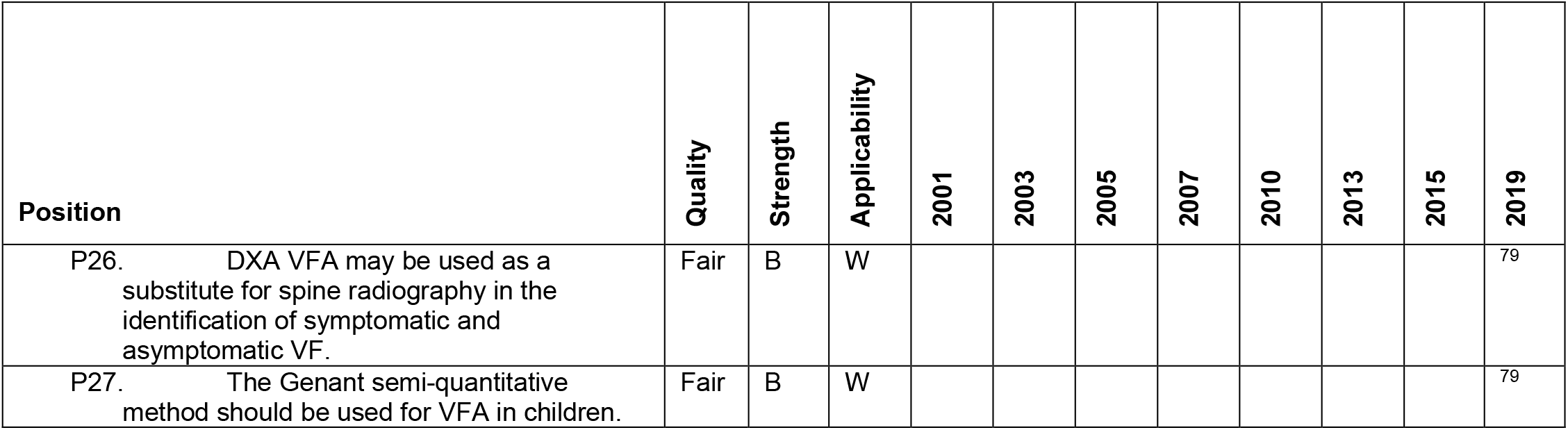
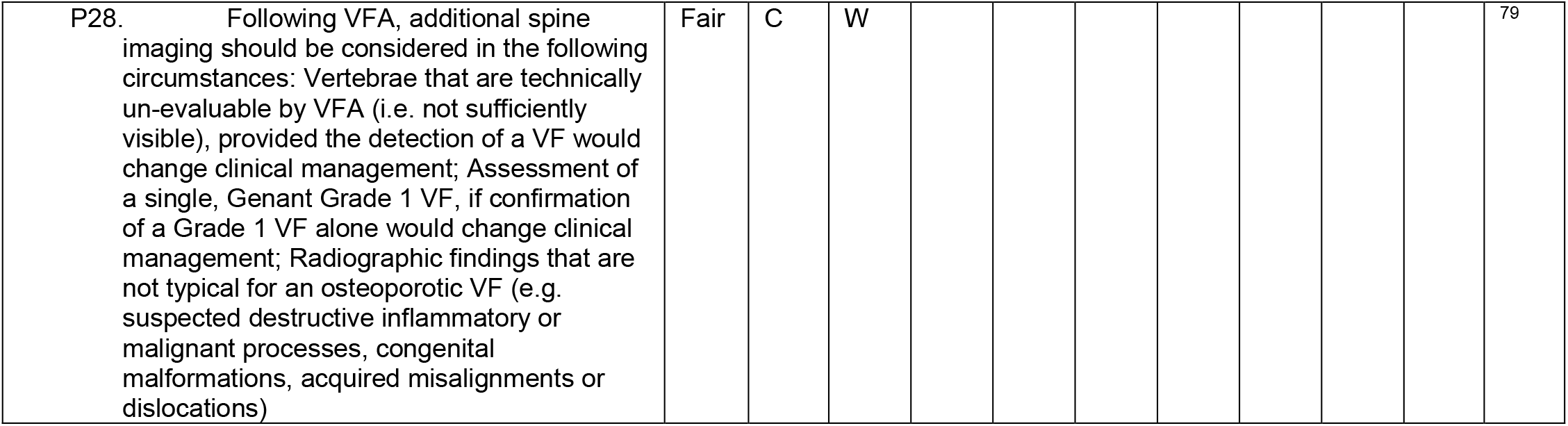

### 5. pQCT in Children and Adolescents

There are currently three positions for pQCT and/or QCT for children listed by the ISCD on their website and all executive summaries since 2013. These three are the result of the 2013 Pediatric PDC. Two of these positions were updated positions from the 2007 Pediatric PDC. The change in wording is noted below the table. There are four additional positions from the 2007 Pediatric PDC that no longer appear in ISCD executive summaries or in their literature. These are listed as positions P34 -P37 and shown in italics. In discusssions with one of the surviving authors (Zemel), this omission is thought to be unintentional editorial deletions. Thus, they are included here and shown in italics. Adams et al. ^62^ provides a thorough presentation of the literature for studies using pQCT and QCT in children and adolescents.

| Position | Quality | Strength | Applicability | 2001 | 2003 | 2005 | 2007 | 2010 | 2013 | 2015 | 2019 |
| --- | --- | --- | --- | --- | --- | --- | --- | --- | --- | --- | --- |
| P29. There is no preferred method for QCT for clinical application in children and adolescents. | Fair | C | W |  |  |  |  |  | 62 |  |  |
| P30. QCT, pQCT, and HR-pQCT are primarily research techniques used to characterize bone deficits in children. They can be used clinically in children where appropriate reference data and expertise are available.* | Fair | B | W |  |  |  | 41 |  | 62 |  |  |
| P31. It is imperative that QCT protocols in children using general CT scanners use appropriate exposure factors, calibration phantoms, and software to optimize results and minimize radiation exposure.** | Good | B | W |  |  |  | 41 |  | 62 |  |  |
| *P32. When the forearm is measured, the non-dominant forearm should be used.* |  |  |  |  |  |  | 41 |  |  |  |  |
| *P33. Measurements sites should include the metaphysis and diaphysis.* |  |  |  |  |  |  | 41 |  |  |  |  |
| *P34. Determination of the precision error, LSC, and monitoring time interval should be performed as described for DXA.* |  |  |  |  |  |  | 41 |  |  |  |  |
| *P35. pQCT reports should include*<br>*Manufacturer, model and software version*<br>*Referring physician Patient age, gender, race/ethnicity, weight and height Relevant medical history including previous fractures*<br>*Indication for measurement Bone age results, if available Measurement site Limb length Scan acquisition and analysis parameters Scan technical quality Reference data source for Z-score calculation*<br>*Metaphyseal total and trabecular vBMD and Z-scores Diaphyseal BMC, cortical vBMD,* |  |  |  |  |  |  | 41 |  |  |  |  |
| cortical thickness, cross-sectional moment of inertia, and SSI results and Z-scores.<br>Adjustments made for growth and maturation Interpretation |  |  |  |  |  |  |  |  |  |  |  |

\* Original wording, "Reference data are not sufficient for the clinical use of pQCT for fracture prediction or diagnosis of low bone mass." \*\* Original wording, "Quality control procedures should be performed as described for central DXA."

### 6. Densitometry in Infants and Young Children

These are the only positions that directly address bone densitometry in Infants and children less than 5 years old. They were not included in the 2007 pediatric PDC. The primary paper, Kalkwarf et al. ^61^ offers not only justification for each position, but a comprehensive literature review for densitometry in young children. They found that there was insufficient information regarding methodology, reproducibility, and reference data to recommended forearm and femur measurements, and regarding to methods for accounting for growth delay as is done in older children Even though the positions document stated the positions were likely to be revisited as more data becomes available, they have not in the past 8 years most likely due to a continued lack of additional data.

| Position | Quality | Strength | Applicability | 2001 | 2003 | 2005 | 2007 | 2010 | 2013 | 2015 | 2019 |
| --- | --- | --- | --- | --- | --- | --- | --- | --- | --- | --- | --- |
| P36. DXA is an appropriate method for clinical densitometry of infants and young children. | Fair | B | W |  |  |  |  |  | 61 |  |  |
| P37. DXA lumbar spine measurements are feasible and can provide reproducible measures of BMC and aBMD for infants and young children 0-5 years of age. | Fair | B | W |  |  |  |  |  | 61 |  |  |
| P38. DXA whole body measurements are feasible and can provide reproducible measures of BMC and aBMD for children $\geq 3$ years of age. | Fair | B | W | | | | | | 61 | | |
| P39. DXA whole body BMC measurements for children < 3 years of age are of limited clinical utility due to feasibility and lack of normative data. Areal BMD should not be utilized routinely due to difficulty inappropriate positioning. | Fair | C | W |  |  |  |  |  | 61 |  |  |
| P40. Forearm and femur measurements are technically feasible in infants and young children, but there is insufficient information regarding methodology, reproducibility, and reference data for these measurements sites to be clinically useful at this time. | Fair | C | W |  |  |  |  |  | 61 |  |  |
| P41. In infants and children below 5 years of age, the impact of growth delay on the interpretation of the DXA results should be considered, but it is not quantifiable presently. | Fair | C | W |  |  |  |  |  | 61 |  |  |

## NOMENCLATURE POSITIONS THAT APPLY TO ADULTS AND PEDIATRICS

### 1. DXA Nomenclature

These positions have only been discussed at the 2003 PDC and summarized in its executive summary (3). The positions were in response to a lack of consistency in reporting key DXA measures. These recommendations have been inconsistently followed in the literature especially in terms of labeling DXA as DEXA. For example, a Google Scholar search performed on 08/25/2021 for papers published in 2020 with either DXA or DEXA in the title found that 22 percent (38/169) of the titles used DEXA. The positions have not been modified since their inception and are broadly used. None of the positions were rated using the RAND criteria.

| Position | Quality | Strength | Applicability | 2001 | 2003 | 2005 | 2007 | 2010 | 2013 | 2015 | 2019 |
| --- | --- | --- | --- | --- | --- | --- | --- | --- | --- | --- | --- |
| N1. DXA – not DEXA. | NA | NA | NA |  | 26 |  |  |  |  |  |  |
| N2. T-score – not T score, t-score, or t score | NA | NA | NA |  | 26 |  |  |  |  |  |  |
| N3. Z-score – not Z score, z-score, or z score | NA | NA | NA |  | 26 |  |  |  |  |  |  |

### 2. DXA Decimal Digits

Like DXA nomenclature, these positions have only been discussed at the 2003 PDC and summarized in its executive summary (3). The positions were in response to a lack of consistency in reporting quantitative measures. The positions have not been modified since their inception and are broadly used. None of the positions were rated using the RAND criteria.

| Position | Quality | Strength | Applicability | 2001 | 2003 | 2005 | 2007 | 2010 | 2013 | 2015 | 2019 |
| --- | --- | --- | --- | --- | --- | --- | --- | --- | --- | --- | --- |
| Preferred number of decimal digits for DXA reporting are as follows: | NA | NA | NA |  | 26 |  |  |  |  |  |  |
| N4. BMD: three digits (e.g., 0.927 g/cm<sup>2</sup>). | NA | NA | NA |  | 26 |  |  |  |  |  |  |
| N5. T-score: one digit (e.g., –2.3). | NA | NA | NA |  | 26 |  |  |  |  |  |  |
| N6. Z-score: one digit (e.g., 1.7). | NA | NA | NA |  | 26 |  |  |  |  |  |  |
| N7. BMC: two digits (e.g., 31.76 g). | NA | NA | NA |  | 26 |  |  |  |  |  |  |
| N8. Area: two digits (e.g., 43.25 cm<sup>2</sup>). | NA | NA | NA |  | 26 |  |  |  |  |  |  |
| N9. % reference database: Integer (e.g., 82%). | NA | NA | NA |  | 26 |  |  |  |  |  |  |

## DISCUSSION

### Summary of evidence

This review has presented the context of the current 290 ISCD positions and provided a means to follow the etiology and pedigree of each position. The positions have been well utilized as evidenced by the over 6,700 cumulative citations to date. Their relevance is increasing as evidenced by the steady increase in annual citations. With over 230 authors participating in the writing and rationale for these positions, they are the most vetted and complete set of guidelines on the appropriate use of clinical densitometry. However, many positions have gone through extensive revisions while others have not been modified in over 20 years.

Before this scoping review, there was no single guide to the origins and rationale for the ISCD Positions. It could be argued that without this context, a clinician is blindly following an established paradigm without understanding of when, how and why it is appropriate.

### Limitations

This scoping review, like all reviews, will be out of date as soon as it is published. But this is the nature of reviews. It is a snapshot of the field and its position at the time of this writing. Future researchers can hopefully use this as a starting point for updates after future PDCs. The review was also limited to the positions of the ISCD exclusively. There are positions for the use of DXA and guidelines for diagnosing osteoporosis from other organizations including the World Health Organization, National Osteoporosis Foundation and the International Osteoporosis Foundation. These were considered out of scope.

### Conclusions

With 290 official positions, the ISCD has created one of the most complete and well documented set of recommendations for the use of DXA, QCT, pQCT and QUS to evaluate bone health in adults and children. This review links all source documents and wording changes to the current positions in the hope that it will be of use to those considering updates to the positions, and as an educational tool for those that seek a deeper understanding of how osteoporosis is managed and diagnosed. Updates to this review are warranted after each PDC.

## Data Availability

All data generated produced for this review is contained in the manuscript

## ACKNOWLEDGEMENTS

We appreciate the contributions from the following individuals: Geraldine Ragsac for her extensive research and copywriting.

## REFERENCES

1. Engelke K, Adams JE, Armbrecht G, Augat P, Bogado CE, Bouxsein ML, Felsenberg D, Ito M, Prevrhal S, Hans DB, Lewiecki EM. Clinical use of quantitative computed tomography and peripheral quantitative computed tomography in the management of osteoporosis in adults: the 2007 ISCD Official Positions. J Clin Densitom. 2008;11(1):123–62. Epub 2008/04/30. doi: 10.1016/j.jocd.2007.12.010. PubMed PMID: 18442757.

2. Peters MD, Marnie C, Tricco AC, Pollock D, Munn Z, Alexander L, McInerney P, Godfrey CM, Khalil H. Updated methodological guidance for the conduct of scoping reviews. JBI Evidence Synthesis. 2020;18(10):2119–26.

3. Harzing A-W, Alakangas S. Google Scholar, Scopus and the Web of Science: a longitudinal and cross-disciplinary comparison. Scientometrics. 2016;106(2):787–804.

4. Densitometry ISfC. Official Positions: ISCD; 2023 [cited 2023 01/28/2023]Adult, Pediatric, Frax Positions]. Available from: https://iscd.org/learn/official-positions/.

5. Lenchik L, Leib ES, Hamdy RC, Binkley NC, Miller PD, Watts NB, International Society for Clinical Densitometry Position Development P, Scientific Advisory C. Executive summary International Society for Clinical Densitometry position development conference Denver, Colorado July 20-22, 2001. J Clin Densitom. 2002;5 Suppl(3, Supplement):S1–3. doi: 10.1385/jcd:5:3s:s01. PubMed PMID: 12464705.

6. Writing Group for the IPDC. Position statement: executive summary. The Writing Group for the International Society for Clinical Densitometry (ISCD) Position Development Conference. J Clin Densitom. 2004;7(1):7–12. doi: 10.1385/jcd:7:1:7. PubMed PMID: 14742882.

7. Binkley N, Bilezikian JP, Kendler DL, Leib ES, Lewiecki EM, Petak SM, International Society for Clinical D. Official positions of the International Society for Clinical Densitometry and Executive Summary of the 2005 Position Development Conference. J Clin Densitom. 2006;9(1):4–14. Epub 20060512. doi: 10.1016/j.jocd.2006.05.002. PubMed PMID: 16731426.

8. Baim S, Binkley N, Bilezikian JP, Kendler DL, Hans DB, Lewiecki EM, Silverman S. Official Positions of the International Society for Clinical Densitometry and executive summary of the 2007 ISCD Position Development Conference. J Clin Densitom. 2008;11(1):75–91. Epub 2008/04/30. doi: 10.1016/j.jocd.2007.12.007. PubMed PMID: 18442754.

9. Baim S, Leonard MB, Bianchi ML, Hans DB, Kalkwarf HJ, Langman CB, Rauch F. Official Positions of the International Society for Clinical Densitometry and executive summary of the 2007 ISCD Pediatric Position Development Conference. J Clin Densitom. 2008;11(1):6–21. Epub 2008/04/30. doi: 10.1016/j.jocd.2007.12.002. PubMed PMID: 18442749.

10. Hans DB, Kanis JA, Baim S, Bilezikian JP, Binkley N, Cauley JA, Compston JE, Cooper C, Dawson-Hughes B, El-Hajj Fuleihan G, Leslie WD, Lewiecki EM, Luckey MM, McCloskey EV, Papapoulos SE, Poiana C, Rizzoli R, Members FPDC. Joint Official Positions of the International Society for Clinical Densitometry and International Osteoporosis Foundation on FRAX((R)). Executive Summary of the 2010 Position Development Conference on Interpretation and use of FRAX(R) in clinical practice. J Clin Densitom. 2011;14(3):171–80. Epub 2011/08/04. doi: 10.1016/j.jocd.2011.05.007. PubMed PMID: 21810521.

11. Schousboe JT, Shepherd JA, Bilezikian JP, Baim S. Executive summary of the 2013 International Society for Clinical Densitometry Position Development Conference on bone densitometry. J Clin Densitom. 2013;16(4):455–66. Epub 2013/11/05. doi: 10.1016/j.jocd.2013.08.004. PubMed PMID: 24183638.

12. Shepherd JA, Baim S, Bilezikian JP, Schousboe JT. Executive summary of the 2013 International Society for Clinical Densitometry Position Development Conference on Body Composition. J Clin Densitom. 2013;16(4):489–95. Epub 2013/11/05. doi: 10.1016/j.jocd.2013.08.005. PubMed PMID: 24183639.

13. Gordon CM, Leonard MB, Zemel BS, International Society for Clinical D. 2013 Pediatric Position Development Conference: executive summary and reflections. J Clin Densitom. 2014;17(2):219–24. Epub 20140320. doi: 10.1016/j.jocd.2014.01.007. PubMed PMID: 24657108.

14. Shepherd JA, Schousboe JT, Broy SB, Engelke K, Leslie WD. Executive Summary of the 2015 ISCD Position Development Conference on Advanced Measures From DXA and QCT: Fracture Prediction Beyond BMD. J Clin Densitom. 2015;18(3):274–86. Epub 2015/08/19. doi: 10.1016/j.jocd.2015.06.013. PubMed PMID: 26277847.

15. Shuhart CR, Yeap SS, Anderson PA, Jankowski LG, Lewiecki EM, Morse LR, Rosen HN, Weber DR, Zemel BS, Shepherd JA. Executive Summary of the 2019 ISCD Position Development Conference on Monitoring Treatment, DXA Cross-calibration and Least Significant Change, Spinal Cord Injury, Peri-prosthetic and Orthopedic Bone Health, Transgender Medicine, and Pediatrics. J Clin Densitom. 2019;22(4):453–71. Epub 20190705. doi: 10.1016/j.jocd.2019.07.001. PubMed PMID: 31400968.

16. Leib ES, Lenchik L, Bilezikian JP, Maricic MJ, Watts NB. Position statements of the International Society for Clinical Densitometry: methodology. J Clin Densitom. 2002;5 Suppl(3, Supplement):S5–10. doi: 10.1385/jcd:5:3s:s05. PubMed PMID: 12464706.

17. Hamdy RC, Petak SM, Lenchik L, International Society for Clinical Densitometry Position Development P, Scientific Advisory C. Which central dual X-ray absorptiometry skeletal sites and regions of interest should be used to determine the diagnosis of osteoporosis? J Clin Densitom. 2002;5 Suppl(3, Supplement):S11–8. doi: 10.1385/jcd:5:3s:s11. PubMed PMID: 12464707.

18. Binkley NC, Schmeer P, Wasnich RD, Lenchik L, International Society for Clinical Densitometry Position Development P, Scientific Advisory C. What are the criteria by which a densitometric diagnosis of osteoporosis can be made in males and non-Caucasians? J Clin Densitom. 2002;5 Suppl(3, Supplement):S19–27. doi: 10.1385/jcd:5:3s:s19. PubMed PMID: 12464708.

19. Lenchik L, Kiebzak GM, Blunt BA, International Society for Clinical Densitometry Position Development P, Scientific Advisory C. What is the role of serial bone mineral density measurements in patient management? J Clin Densitom. 2002;5 Suppl(3, Supplement):S29–38. doi: 10.1385/jcd:5:3s:s29. PubMed PMID: 12464709.

20. Miller PD, Njeh CF, Jankowski LG, Lenchik L, International Society for Clinical Densitometry Position Development P, Scientific Advisory C. What are the standards by which bone mass measurement at peripheral skeletal sites should be used in the diagnosis of osteoporosis? J Clin Densitom. 2002;5 Suppl(3, Supplement):S39–45. doi: 10.1385/jcd:5:3s:s39. PubMed PMID: 12464710.

21. Leib ES, Lewiecki EM, Binkley N, Hamdy RC, International Society for Clinical D. Official positions of the International Society for Clinical Densitometry. J Clin Densitom. 2004;7(1):1–6. doi: 10.1385/jcd:7:1:1. PubMed PMID: 14742881.

22. Writing Group for the IPDC. Position statement: introduction, methods, and participants. The Writing Group for the International Society for Clinical Densitometry (ISCD) Position Development Conference. J Clin Densitom. 2004;7(1):13–6. doi: 10.1385/jcd:7:1:13. PubMed PMID: 14742883.

23. Writing Group for the IPDC. Diagnosis of osteoporosis in men, premenopausal women, and children. J Clin Densitom. 2004;7(1):17–26. doi: 10.1385/jcd:7:1:17. PubMed PMID: 14742884.

24. Writing Group for the IPDC. Technical standardization for dual-energy x-ray absorptiometry. J Clin Densitom. 2004;7(1):27–36. doi: 10.1385/jcd:7:1:27. PubMed PMID: 14742885.

25. Writing Group for the IPDC. Indications and reporting for dual-energy x-ray absorptiometry. J Clin Densitom. 2004;7(1):37–44. doi: 10.1385/jcd:7:1:37. PubMed PMID: 14742886.

26. Writing Group for the IPDC. Nomenclature and decimal places in bone densitometry. J Clin Densitom. 2004;7(1):45–50. doi: 10.1385/jcd:7:1:45. PubMed PMID: 14742887.

27. Khan AA, Bachrach L, Brown JP, Hanley DA, Josse RG, Kendler DL, Leib ES, Lentle BC, Leslie WD, Lewiecki EM. Standards and guidelines for performing central dual-energy x-ray absorptiometry in premenopausal women, men, and children: a report from the Canadian Panel of the International Society of Clinical Densitometry. Journal of Clinical Densitometry. 2004;7(1):51–63.

28. Khan AA, Brown JP, Kendler DL, Leslie WD, Lentle BC, Lewiecki EM, Miller PD, Nicholson RL, Olszynski WP, Watts NB. The 2002 Canadian bone densitometry recommendations: take-home messages. CMAJ. 2002;167(10):1141–5.

29. Hans D, Downs RW, Jr., Duboeuf F, Greenspan S, Jankowski LG, Kiebzak GM, Petak SM, International Society for Clinical D. Skeletal sites for osteoporosis diagnosis: the 2005 ISCD Official Positions. J Clin Densitom. 2006;9(1):15–21. Epub 20060512. doi: 10.1016/j.jocd.2006.05.003. PubMed PMID: 16731427.

30. Leslie WD, Adler RA, El-Hajj Fuleihan G, Hodsman AB, Kendler DL, McClung M, Miller PD, Watts NB, International Society for Clinical D. Application of the 1994 WHO classification to populations other than postmenopausal Caucasian women: the 2005 ISCD Official Positions. J Clin Densitom. 2006;9(1):22–30. Epub 20060512. doi: 10.1016/j.jocd.2006.05.004. PubMed PMID: 16731428.

31. Shepherd JA, Lu Y, Wilson K, Fuerst T, Genant H, Hangartner TN, Wilson C, Hans D, Leib ES, International Society for Clinical Densitometry Committee on Standards of Bone M. Cross-calibration and minimum precision standards for dual-energy X-ray absorptiometry: the 2005 ISCD Official Positions. J Clin Densitom. 2006;9(1):31–6. Epub 20060512. doi: 10.1016/j.jocd.2006.05.005. PubMed PMID: 16731429.

32. Vokes T, Bachman D, Baim S, Binkley N, Broy S, Ferrar L, Lewiecki EM, Richmond B, Schousboe J, International Society for Clinical D. Vertebral fracture assessment: the 2005 ISCD Official Positions. J Clin Densitom. 2006;9(1):37–46. Epub 20060512. doi: 10.1016/j.jocd.2006.05.006. PubMed PMID: 16731430.

33. Schousboe JT, Vokes T, Broy SB, Ferrar L, McKiernan F, Roux C, Binkley N. Vertebral Fracture Assessment: the 2007 ISCD Official Positions. J Clin Densitom. 2008;11(1):92–108. Epub 2008/04/30. doi: 10.1016/j.jocd.2007.12.008. PubMed PMID: 18442755.

34. Simonelli C, Adler RA, Blake GM, Caudill JP, Khan A, Leib E, Maricic M, Prior JC, Eis SR, Rosen C, Kendler DL. Dual-Energy X-Ray Absorptiometry Technical issues: the 2007 ISCD Official Positions. J Clin Densitom. 2008;11(1):109–22. Epub 2008/04/30. doi: 10.1016/j.jocd.2007.12.009. PubMed PMID: 18442756.

35. Krieg MA, Barkmann R, Gonnelli S, Stewart A, Bauer DC, Del Rio Barquero L, Kaufman JJ, Lorenc R, Miller PD, Olszynski WP, Poiana C, Schott AM, Lewiecki EM, Hans D. Quantitative ultrasound in the management of osteoporosis: the 2007 ISCD Official Positions. J Clin Densitom. 2008;11(1):163–87. Epub 2008/04/30. doi: 10.1016/j.jocd.2007.12.011. PubMed PMID: 18442758.

36. Hans DB, Shepherd JA, Schwartz EN, Reid DM, Blake GM, Fordham JN, Fuerst T, Hadji P, Itabashi A, Krieg MA, Lewiecki EM. Peripheral dual-energy X-ray absorptiometry in the management of osteoporosis: the 2007 ISCD Official Positions. J Clin Densitom. 2008;11(1):188–206. Epub 2008/04/30. doi: 10.1016/j.jocd.2007.12.012. PubMed PMID: 18442759.

37. Fitch K, Bernstein S, Aguilar M. The RAND/UCLA Appropriateness Method User’s Manual. 2001.

38. Rauch F, Plotkin H, DiMeglio L, Engelbert RH, Henderson RC, Munns C, Wenkert D, Zeitler P. Fracture prediction and the definition of osteoporosis in children and adolescents: the ISCD 2007 Pediatric Official Positions. J Clin Densitom. 2008;11(1):22–8. Epub 2008/04/30. doi: 10.1016/j.jocd.2007.12.003. PubMed PMID: 18442750.

39. Bishop N, Braillon P, Burnham J, Cimaz R, Davies J, Fewtrell M, Hogler W, Kennedy K, Makitie O, Mughal Z, Shaw N, Vogiatzi M, Ward K, Bianchi ML. Dual-energy X-ray aborptiometry assessment in children and adolescents with diseases that may affect the skeleton: the 2007 ISCD Pediatric Official Positions. J Clin Densitom. 2008;11(1):29–42. Epub 2008/04/30. doi: 10.1016/j.jocd.2007.12.004. PubMed PMID: 18442751.

40. Gordon CM, Bachrach LK, Carpenter TO, Crabtree N, El-Hajj Fuleihan G, Kutilek S, Lorenc RS, Tosi LL, Ward KA, Ward LM, Kalkwarf HJ. Dual energy X-ray absorptiometry interpretation and reporting in children and adolescents: the 2007 ISCD Pediatric Official Positions. J Clin Densitom. 2008;11(1):43–58. Epub 2008/04/30. doi: 10.1016/j.jocd.2007.12.005. PubMed PMID: 18442752.

41. Zemel B, Bass S, Binkley T, Ducher G, Macdonald H, McKay H, Moyer-Mileur L, Shepherd J, Specker B, Ward K, Hans D. Peripheral quantitative computed tomography in children and adolescents: the 2007 ISCD Pediatric Official Positions. J Clin Densitom. 2008;11(1):59–74. Epub 2008/04/30. doi: 10.1016/j.jocd.2007.12.006. PubMed PMID: 18442753.

42. McCloskey EV, Binkley N, Members FPDC. FRAX((R)) Clinical Task Force of the 2010 Joint International Society for Clinical Densitometry & International Osteoporosis Foundation Position Development Conference. J Clin Densitom. 2011;14(3):181–3. Epub 2011/08/04. doi: 10.1016/j.jocd.2011.05.013. PubMed PMID: 21810522.

43. Broy SB, Tanner SB, Members FRDC. Official Positions for FRAX(R) clinical regarding rheumatoid arthritis from Joint Official Positions Development Conference of the International Society for Clinical Densitometry and International Osteoporosis Foundation on FRAX(R). J Clin Densitom. 2011;14(3):184–9. Epub 2011/08/04. doi: 10.1016/j.jocd.2011.05.012. PubMed PMID: 21810523.

44. Dimai HP, Chandran M, Members FRDC. Official Positions for FRAX(R) clinical regarding smoking from Joint Official Positions Development Conference of the International Society for Clinical Densitometry and International Osteoporosis Foundation on FRAX(R). J Clin Densitom. 2011;14(3):190–3. Epub 2011/08/04. doi: 10.1016/j.jocd.2011.05.011. PubMed PMID: 21810524.

45. Masud T, Binkley N, Boonen S, Hannan MT, Members FPDC. Official Positions for FRAX(R) clinical regarding falls and frailty: can falls and frailty be used in FRAX(R)? From Joint Official Positions Development Conference of the International Society for Clinical Densitometry and International Osteoporosis Foundation on FRAX(R). J Clin Densitom. 2011;14(3):194–204. Epub 2011/08/04. doi: 10.1016/j.jocd.2011.05.010. PubMed PMID: 21810525.

46. Blank RD, Members FPDC. Official Positions for FRAX(R) clinical regarding prior fractures from Joint Official Positions Development Conference of the International Society for Clinical Densitometry and International Osteoporosis Foundation on FRAX(R). J Clin Densitom. 2011;14(3):205–11. Epub 2011/08/04. doi: 10.1016/j.jocd.2011.05.009. PubMed PMID: 21810526; PMCID: PMC6819950.

47. Leib ES, Saag KG, Adachi JD, Geusens PP, Binkley N, McCloskey EV, Hans DB, Members FPDC. Official Positions for FRAX((R)) clinical regarding glucocorticoids: the impact of the use of glucocorticoids on the estimate by FRAX((R)) of the 10 year risk of fracture from Joint Official Positions Development Conference of the International Society for Clinical Densitometry and International Osteoporosis Foundation on FRAX((R)). J Clin Densitom. 2011;14(3):212–9. Epub 2011/08/04. doi: 10.1016/j.jocd.2011.05.014. PubMed PMID: 21810527.

48. McCloskey EV, Vasikaran S, Cooper C, Members FPDC. Official Positions for FRAX(R) clinical regarding biochemical markers from Joint Official Positions Development Conference of the International Society for Clinical Densitometry and International Osteoporosis Foundation on FRAX(R). J Clin Densitom. 2011;14(3):220–2. Epub 2011/08/04. doi: 10.1016/j.jocd.2011.05.008. PubMed PMID: 21810528.

49. Lewiecki EM, Compston JE, Miller PD, Adachi JD, Adams JE, Leslie WD, Kanis JA, Members FPDC. FRAX((R)) Bone Mineral Density Task Force of the 2010 Joint International Society for Clinical Densitometry & International Osteoporosis Foundation Position Development Conference. J Clin Densitom. 2011;14(3):223–5. Epub 2011/08/04. doi: 10.1016/j.jocd.2011.05.018. PubMed PMID: 21810529.

50. Lewiecki EM, Compston JE, Miller PD, Adachi JD, Adams JE, Leslie WD, Kanis JA, Moayyeri A, Adler RA, Hans DB, Kendler DL, Diez-Perez A, Krieg MA, Masri BK, Lorenc RR, Bauer DC, Blake GM, Josse RG, Clark P, Khan AA, Members FPDC. Official Positions for FRAX(R) Bone Mineral Density and FRAX(R) simplification from Joint Official Positions Development Conference of the International Society for Clinical Densitometry and International Osteoporosis Foundation on FRAX(R). J Clin Densitom. 2011;14(3):226–36. Epub 2011/08/04. doi: 10.1016/j.jocd.2011.05.017. PubMed PMID: 21810530.

51. Cauley JA, El-Hajj Fuleihan G, Luckey MM, Members FPDC. FRAX(R) International Task Force of the 2010 Joint International Society for Clinical Densitometry & International Osteoporosis Foundation Position Development Conference. J Clin Densitom. 2011;14(3):237–9. Epub 2011/08/04. doi: 10.1016/j.jocd.2011.05.016. PubMed PMID: 21810531.

52. Cauley JA, El-Hajj Fuleihan G, Arabi A, Fujiwara S, Ragi-Eis S, Calderon A, Chionh SB, Chen Z, Curtis JR, Danielson ME, Hanley DA, Kroger H, Kung AW, Lesnyak O, Nieves J, Pluskiewicz W, El Rassi R, Silverman S, Schott AM, Rizzoli R, Luckey M, Members FPC. Official Positions for FRAX(R) clinical regarding international differences from Joint Official Positions Development Conference of the International Society for Clinical Densitometry and International Osteoporosis Foundation on FRAX(R). J Clin Densitom. 2011;14(3):240–62. Epub 2011/08/04. doi: 10.1016/j.jocd.2011.05.015. PubMed PMID: 21810532.

53. Kanis J, Johnell O, Odén A, Johansson H, McCloskey E. FRAX™ and the assessment of fracture probability in men and women from the UK. Osteoporosis international. 2008;19(4):385–97.

54. Malabanan AO, Rosen HN, Vokes TJ, Deal CL, Alele JD, Olenginski TP, Schousboe JT. Indications of DXA in women younger than 65 yr and men younger than 70 yr: the 2013 Official Positions. J Clin Densitom. 2013;16(4):467–71. Epub 20130920. doi: 10.1016/j.jocd.2013.08.002. PubMed PMID: 24055260.

55. Watts NB, Leslie WD, Foldes AJ, Miller PD. 2013 International Society for Clinical Densitometry Position Development Conference: Task Force on Normative Databases. J Clin Densitom. 2013;16(4):472–81. Epub 20130926. doi: 10.1016/j.jocd.2013.08.001. PubMed PMID: 24076161.

56. Rosen HN, Vokes TJ, Malabanan AO, Deal CL, Alele JD, Olenginski TP, Schousboe JT. The Official Positions of the International Society for Clinical Densitometry: vertebral fracture assessment. J Clin Densitom. 2013;16(4):482–8. Epub 20130922. doi: 10.1016/j.jocd.2013.08.003. PubMed PMID: 24063846.

57. Kendler DL, Borges JL, Fielding RA, Itabashi A, Krueger D, Mulligan K, Camargos BM, Sabowitz B, Wu CH, Yu EW, Shepherd J. The Official Positions of the International Society for Clinical Densitometry: Indications of Use and Reporting of DXA for Body Composition. J Clin Densitom. 2013;16(4):496–507. Epub 20131016. doi: 10.1016/j.jocd.2013.08.020. PubMed PMID: 24090645.

58. Petak S, Barbu CG, Yu EW, Fielding R, Mulligan K, Sabowitz B, Wu CH, Shepherd JA. The Official Positions of the International Society for Clinical Densitometry: body composition analysis reporting. J Clin Densitom. 2013;16(4):508–19. Epub 2013/11/05. doi: 10.1016/j.jocd.2013.08.018. PubMed PMID: 24183640.

59. Hangartner TN, Warner S, Braillon P, Jankowski L, Shepherd J. The Official Positions of the International Society for Clinical Densitometry: acquisition of dual-energy X-ray absorptiometry body composition and considerations regarding analysis and repeatability of measures. J Clin Densitom. 2013;16(4):520–36. Epub 2013/11/05. doi: 10.1016/j.jocd.2013.08.007. PubMed PMID: 24183641.

60. Crabtree NJ, Arabi A, Bachrach LK, Fewtrell M, El-Hajj Fuleihan G, Kecskemethy HH, Jaworski M, Gordon CM, International Society for Clinical D. Dual-energy X-ray absorptiometry interpretation and reporting in children and adolescents: the revised 2013 ISCD Pediatric Official Positions. J Clin Densitom. 2014;17(2):225–42. Epub 20140329. doi: 10.1016/j.jocd.2014.01.003. PubMed PMID: 24690232.

61. Kalkwarf HJ, Abrams SA, DiMeglio LA, Koo WW, Specker BL, Weiler H, International Society for Clinial D. Bone densitometry in infants and young children: the 2013 ISCD Pediatric Official Positions. J Clin Densitom. 2014;17(2):243–57. Epub 20140324. doi: 10.1016/j.jocd.2014.01.002. PubMed PMID: 24674638.

62. Adams JE, Engelke K, Zemel BS, Ward KA, International Society of Clinical D. Quantitative computer tomography in children and adolescents: the 2013 ISCD Pediatric Official Positions. J Clin Densitom. 2014;17(2):258–74. Epub 2014/05/06. doi: 10.1016/j.jocd.2014.01.006. PubMed PMID: 24792821.

63. Bishop N, Arundel P, Clark E, Dimitri P, Farr J, Jones G, Makitie O, Munns CF, Shaw N, International Society of Clinical D. Fracture prediction and the definition of osteoporosis in children and adolescents: the ISCD 2013 Pediatric Official Positions. J Clin Densitom. 2014;17(2):275–80. Epub 20140314. doi: 10.1016/j.jocd.2014.01.004. PubMed PMID: 24631254.

64. Bianchi ML, Leonard MB, Bechtold S, Hogler W, Mughal MZ, Schonau E, Sylvester FA, Vogiatzi M,van den Heuvel-Eibrink MM, Ward L, International Society for Clinical D. Bone health in children and adolescents with chronic diseases that may affect the skeleton: the 2013 ISCD Pediatric Official Positions. J Clin Densitom. 2014;17(2):281–94. Epub 20140319. doi: 10.1016/j.jocd.2014.01.005. PubMed PMID: 24656723.

65. Broy SB, Cauley JA, Lewiecki ME, Schousboe JT, Shepherd JA, Leslie WD. Fracture Risk Prediction by Non-BMD DXA Measures: the 2015 ISCD Official Positions Part 1: Hip Geometry. J Clin Densitom. 2015;18(3):287–308. Epub 2015/08/19. doi: 10.1016/j.jocd.2015.06.005. PubMed PMID: 26277848.

66. Silva BC, Broy SB, Boutroy S, Schousboe JT, Shepherd JA, Leslie WD. Fracture Risk Prediction by Non-BMD DXA Measures: the 2015 ISCD Official Positions Part 2: Trabecular Bone Score. J Clin Densitom. 2015;18(3):309–30. Epub 2015/08/19. doi: 10.1016/j.jocd.2015.06.008. PubMed PMID: 26277849.

67. Beck TJ, Broy SB. Measurement of Hip Geometry-Technical Background. J Clin Densitom. 2015;18(3):331–7. Epub 2015/08/19. doi: 10.1016/j.jocd.2015.06.006. PubMed PMID: 26277850.

68. Engelke K, Lang T, Khosla S, Qin L, Zysset P, Leslie WD, Shepherd JA, Schousboe JT. Clinical Use of Quantitative Computed Tomography (QCT) of the Hip in the Management of Osteoporosis in Adults: the 2015 ISCD Official Positions-Part I. J Clin Densitom. 2015;18(3):338–58. Epub 2015/08/19. doi: 10.1016/j.jocd.2015.06.012. PubMed PMID: 26277851.

69. Zysset P, Qin L, Lang T, Khosla S, Leslie WD, Shepherd JA, Schousboe JT, Engelke K. Clinical Use of Quantitative Computed Tomography-Based Finite Element Analysis of the Hip and Spine in the Management of Osteoporosis in Adults: the 2015 ISCD Official Positions-Part II. J Clin Densitom. 2015;18(3):359–92. Epub 2015/08/19. doi: 10.1016/j.jocd.2015.06.011. PubMed PMID: 26277852.

70. Engelke K, Lang T, Khosla S, Qin L, Zysset P, Leslie WD, Shepherd JA, Shousboe JT. Clinical Use of Quantitative Computed Tomography-Based Advanced Techniques in the Management of Osteoporosis in Adults: the 2015 ISCD Official Positions-Part III. J Clin Densitom. 2015;18(3):393–407. Epub 2015/08/19. doi: 10.1016/j.jocd.2015.06.010. PubMed PMID: 26277853.

71. Jankowski LG, Warner S, Gaither K, Lenchik L, Fan B, Lu Y, Shepherd J. Cross-calibration, Least Significant Change and Quality Assurance in Multiple Dual-Energy X-ray Absorptiometry Scanner Environments: 2019 ISCD Official Position. J Clin Densitom. 2019;22(4):472–83. Epub 20190907. doi: 10.1016/j.jocd.2019.09.001. PubMed PMID: 31558404.

72. Borges JLC, Sousa da Silva M, Ward RJ, Diemer KM, Yeap SS, Lewiecki EM. Repeating Vertebral Fracture Assessment: 2019 ISCD Official Position. J Clin Densitom. 2019;22(4):484–8. Epub 20190710. doi: 10.1016/j.jocd.2019.07.005. PubMed PMID: 31375350.

73. Kendler DL, Compston J, Carey JJ, Wu CH, Ibrahim A, Lewiecki EM. Repeating Measurement of Bone Mineral Density when Monitoring with Dual-energy X-ray Absorptiometry: 2019 ISCD Official Position. J Clin Densitom. 2019;22(4):489–500. Epub 20190717. doi: 10.1016/j.jocd.2019.07.010. PubMed PMID: 31378452.

74. Krohn K, Schwartz EN, Chung YS, Lewiecki EM. Dual-energy X-ray Absorptiometry Monitoring with Trabecular Bone Score: 2019 ISCD Official Position. J Clin Densitom. 2019;22(4):501–5. Epub 20190709. doi: 10.1016/j.jocd.2019.07.006. PubMed PMID: 31383412.

75. Cheung AM, McKenna MJ, van de Laarschot DM, Zillikens MC, Peck V, Srighanthan J, Lewiecki EM. Detection of Atypical Femur Fractures. J Clin Densitom. 2019;22(4):506–16. Epub 20190710. doi: 10.1016/j.jocd.2019.07.003. PubMed PMID: 31377055.

76. Anderson PA, Morgan SL, Krueger D, Zapalowski C, Tanner B, Jeray KJ, Krohn KD, Lane JP, Yeap SS, Shuhart CR, Shepherd J. Use of Bone Health Evaluation in Orthopedic Surgery: 2019 ISCD Official Position. J Clin Densitom. 2019;22(4):517–43. Epub 20190816. doi: 10.1016/j.jocd.2019.07.013. PubMed PMID: 31519473.

77. Rosen HN, Hamnvik OR, Jaisamrarn U, Malabanan AO, Safer JD, Tangpricha V, Wattanachanya L, Yeap SS. Bone Densitometry in Transgender and Gender Non-Conforming (TGNC) Individuals: 2019 ISCD Official Position. J Clin Densitom. 2019;22(4):544–53. Epub 20190710. doi: 10.1016/j.jocd.2019.07.004. PubMed PMID: 31327665.

78. Morse LR, Biering-Soerensen F, Carbone LD, Cervinka T, Cirnigliaro CM, Johnston TE, Liu N, Troy KL, Weaver FM, Shuhart C, Craven BC. Bone Mineral Density Testing in Spinal Cord Injury: 2019 ISCD Official Position. J Clin Densitom. 2019;22(4):554–66. Epub 20190803. doi: 10.1016/j.jocd.2019.07.012. PubMed PMID: 31501005.

79. Weber DR, Boyce A, Gordon C, Hogler W, Kecskemethy HH, Misra M, Swolin-Eide D, Tebben P, Ward LM, Wasserman H, Shuhart C, Zemel BS. The Utility of DXA Assessment at the Forearm, Proximal Femur, and Lateral Distal Femur, and Vertebral Fracture Assessment in the Pediatric Population: 2019 ISCD Official Position. J Clin Densitom. 2019;22(4):567–89. Epub 20190710. doi: 10.1016/j.jocd.2019.07.002. PubMed PMID: 31421951; PMCID: PMC7010480.

80. Lewiecki EM, Laster AJ, Miller PD, Bilezikian JP. More bone density testing is needed, not less. Journal of Bone and Mineral Research. 2012;27(4):739–42.

81. Black DM, Abrahamsen B, Bouxsein ML, Einhorn T, Napoli N. Atypical femur fractures: review of epidemiology, relationship to bisphosphonates, prevention, and clinical management. Endocrine reviews. 2019;40(2):333–68.

82. Njeh C, Boivin C, Langton C. The role of ultrasound in the assessment of osteoporosis: a review. Osteoporosis International. 1997;7:7–22.

83. Hans D, Métrailler A, Gonzalez Rodriguez E, Lamy O, Shevroja E. Quantitative ultrasound (QUS) in the management of osteoporosis and assessment of fracture risk: an update. Bone Quantitative Ultrasound: New Horizons: Springer; 2022. p. 7–34.

